# Leveraging epigenetic biomarker proxies for precision medicine

**DOI:** 10.1101/2024.12.06.24318612

**Authors:** Natàlia Carreras-Gallo, Qingwen Chen, Laura Balagué-Dobón, Andrea Aparicio, Ilinca M. Giosan, Rita Dargham, Daniel Phelps, Tao Guo, Max Melnikas, Kevin M. Mendez, Yulu Chen, Athena Carangan, Srikar Vempaty, Sayf Hassouneh, Michael McGeachie, Yichen Huang, Dawn L. DeMeo, Scott T. Weiss, Tavis Mendez, Florence Comite, Karsten Suhre, Ryan Smith, Varun B. Dwaraka, Jessica A. Lasky-Su

## Abstract

**Background:** Clinical and molecular biomarkers are central to disease risk stratification, but their direct measurement remains fragmented across cohorts and analytical platforms, limiting scalability and reuse in population-scale studies.

**Results:** We developed DNAm-based surrogates, termed epigenetic biomarker proxies (EBPs), in 4,418 participants from the Massachusetts General Brigham Aging Biobank Cohort (MGB-ABC). Using mutual-information feature selection followed by elastic net regression, with gradient boosting for poorly fitted targets, we retained 1,694 EBPs with a Spearman correlation > 0.2 with their target in held-out data: 42 clinical laboratory traits, 689 metabolites and 963 proteins. We validated EBPs in seven independent cohorts totalling 57,454 participants, spanning paediatric to older-adult populations, differing disease burdens and distinct biospecimen and molecular profiling platforms; biomarker correlations, disease associations and clinical-range detection were broadly replicated.

**Conclusions:** EBPs can be constructed at scale from a single DNAm assay and capture biologically and clinically relevant biological variation across clinical, proteomic and metabolomic domains. Their combination of modest cross-sectional concordance with robust disease association suggests that EBPs may capture biological information that is related to, but not identical to, contemporaneous biomarker concentrations. EBPs provide complementary harmonised proxies for molecular phenotyping and population-scale research, rather than as replacements for direct biomarker measurement.

## Background

The advent of multiomics has brought substantial potential to transform patient treatment into a tailored “personalized” framework. High-dimensional molecular profiling offers the promise of refined risk stratification and disease monitoring. Multi-omic profiles provide important insight into the physiologic processes contributing to health. However, despite rapid technological advances, the clinical impact of multi-omics has fallen short of their transformative potential [1]. A major drawback is that omic profiling does not fit into the context of the current clinical system. It is impractical on a large scale due to prohibitive costs, repeated sample draws, and complex data interpretation. These are among the critical barriers that must be resolved in order for broad-scale adoption among practitioners and the full realization of its potential. Addressing these barriers is essential for translating molecular insights into routine clinical care.

Among the layers of multi-omics, epigenetic changes stand out for regulating gene expression while being influenced by environmental and lifestyle factors including diet, exercise, smoking, and alcohol consumption [2,3]. DNA methylation (DNAm) patterns have been shown to provide dynamic insight into physiological responses to environmental stressors across the lifespan and are considered promising predictive biomarkers across a broad spectrum of health outcomes [3–6]. The DNAm-based research has enabled the development of robust phenotypic algorithms, including biological age clocks and methylation risk scores (MRSs), for predicting aging-related outcomes and disease risk [7,8]. MRSs have demonstrated high accuracy in diagnosing and predicting disease, often outperforming polygenic risk scores (PRSs) [7,8].

In recent years, DNAm data has been further used to create surrogate measures (a.k.a. DNAm scores, DNAm surrogates, EpiScores) of proteins and metabolites, enabling access to otherwise costly or inaccessible clinical metrics. DNAm algorithms have been developed to estimate selected blood proteins and metabolites, capturing shifts in inflammation, metabolic function, and environmental exposures [9–13]. Recent work has demonstrated the ability of DNAm surrogates to reflect relevant biology and serve as markers of disease, augmenting existing clinical biomarkers for risk stratification [12,14,15]. Further, DNAm surrogates have been integrated into the development of biological aging clocks, showing improved predictive accuracy for longevity and strong associations with age-related chronic disease outcomes [4,14,15], and their clinical relevance has led to the development of diagnostic tests for Mendelian genetic disorders [16]. However, existing studies have focused on limited biomarker sets and have not systematically evaluated the validity, longitudinal performance, or generalizability of DNAm surrogates across diverse molecular traits and clinical outcomes. Large-scale validation across independent cohorts also remains necessary to establish their clinical utility.

Here, we address this gap by building DNAm surrogates, termed epigenetic biomarker proxies (EBPs) of approximately 1,600 clinical, metabolomic, and proteomic measures from over 4,000 participants in the Massachusetts General Brigham’s Aging Biobank Cohort (MGB-ABC) [15]. We evaluate correlations between EBPs and the corresponding observed measurements and compare the effect estimates of associations with health outcomes, observing consistent patterns. Using multiple analytic frameworks, we identified sets of EBPs that have important clinical relevance and can inform underlying pathobiology. Importantly, we validate these findings across seven independent cohorts, ranging from healthy volunteers to multimorbid patients, to confirm their robustness and applicability in diverse clinical settings. Together, these findings establish EBPs as a scalable framework for integrating molecular information with longitudinal clinical data, with potential to monitor disease and inform future clinical risk.

## Results

### Study design and multi-cohort validation framework

We developed DNAm surrogates, termed epigenetic biomarker proxies (EBPs), to approximate clinical measures, metabolites, and proteins using 4,418 participants from the Mass General Brigham Aging Biobank Cohort (MGB-ABC) as a development cohort (Figure 1A). EBPs were trained using elastic net and gradient-boosted models and evaluated in held-out internal testing data. We then validated EBP algorithms across seven independent external cohorts (total n = 57,454), including six cohorts (n = 26,442) spanning diverse age distributions, disease burdens, and biospecimen platforms, and an additional external cohort (TruDiagnostic biobank, n = 31,012) used to evaluate biomarker concordance, disease associations, classification of clinically defined abnormal laboratory values, and real-world and longitudinal behavior of EBPs. (Figure 1B; Additional file 2: Table S1).

**Figure 1.**
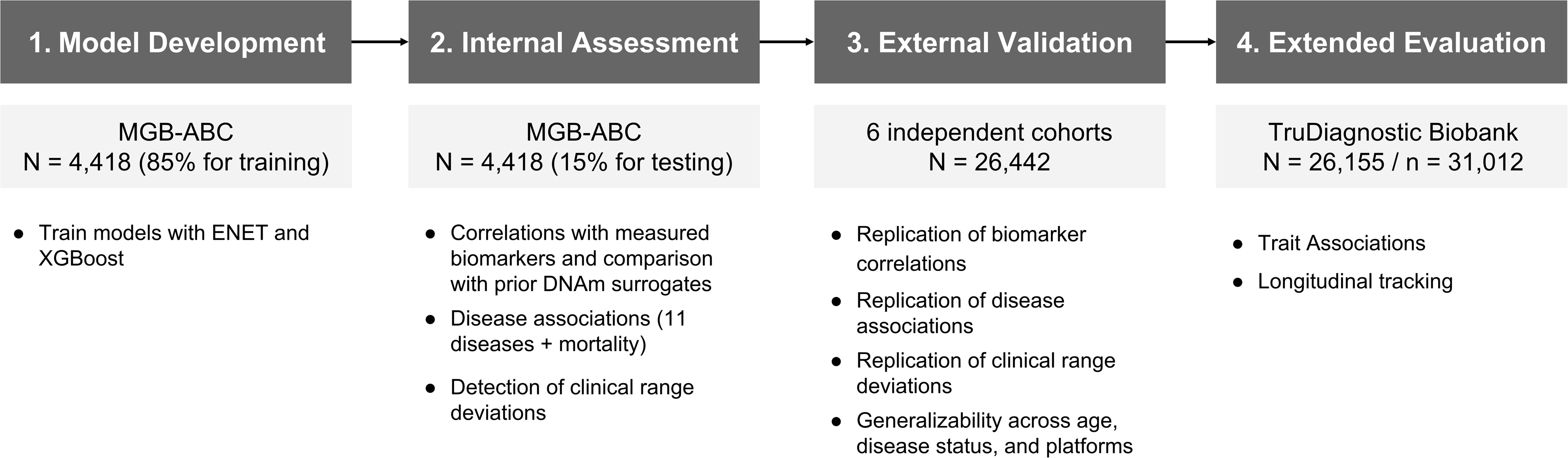
Study design and multi-cohort validation framework. (A) DNA methylation– based epigenetic biomarker proxies (EBPs) were developed and internally tested in 4,418 participants from the Massachusetts General Brigham Aging Biobank Cohort (MGB-ABC). Trained models were then applied, without retraining, to six independent external cohorts (total n = 26,442) spanning diverse age ranges, disease burdens, and biospecimen platforms. External analyses using the test set from MGB-ABC and an additional external cohort (TruDiagnostic biobank, n = 31,012) evaluated biomarker concordance, disease associations, classification of clinically defined abnormal laboratory values, and real-world and longitudinal behavior of EBPs. (B) Summary of the cohorts used in the study, classified by sample size, age range, whether they are enriched in any disease, data types used for the different analyses, and the phase where they were used. (C) Flowchart showing the development of the EBPs. At the top, the total number of biomarkers available in the MGB-ABC. At the bottom, the total number of EBPs selected.

Validation analyses comprised four complementary evaluations: (i) concordance between EBPs and observed biomarkers, (ii) replication of incident and prevalent biomarker-disease associations, (iii) detection of out-of-clinical-range values, and (iv) assessment of real-world and longitudinal EBP behavior across independent datasets. Together, this framework tests the generalizability and clinical relevance of EBPs across populations and study designs.

### Development of epigenetic biomarker proxies in the MGB-ABC cohort

We developed a machine learning pipeline to train EBPs for 1,150 metabolites (Metabolon), 1,979 proteins (Seer SP100), and 44 clinical measures derived from electronic health records (EHRs). The pipeline integrates mutual information-based CpG feature selection with regularized regression or gradient-boosted modeling. Top informative CpG sites were selected to reduce dimensionality, and data were split into training and testing sets to evaluate model performance. Elastic-net regression models were trained across a range of regularization settings, with optimized models selected based on minimized prediction error. For biomarkers underfit by elastic-net model, XGBoost was applied to capture non-linear relationships, retaining models that achieved a correlation of at least 0.2 with predicted targets. This framework yielded 1,694 EBPs for downstream analyses, comprising 689 metabolite, 963 protein, and 42 clinical EBPs (Figure 1C).

### Comparison of EBP concordance with observed biomarkers and prior DNAm surrogates

We assessed concordance between EBPs and observed biomarkers in the internal testing set using Spearman correlation (Additional file 1: Fig. S1A). Across molecular classes, EBPs showed moderate-to-strong agreement with observed values. Clinical EBPs demonstrated the highest concordance on average (mean: 0.41; range: 0.23– 0.67) while mean correlations were 0.29 for both metabolite (range: 0.20–0.59) and protein (range: 0.20–0.54) EBPs (Additional file 2: Table S2, Table S3, Table S4). Because EBPs were retained conditional on achieving a Spearman correlation ≥ 0.2 in this same held-out test set, these internal estimates may be subject to bias and are best interpreted alongside the fully external validation results reported below. A total of 21 clinical, 71 metabolite, and 153 protein EBPs achieved correlations greater than 0.4. Pathway-level analyses revealed heterogeneity in concordance, with nucleotide-associated metabolites exhibiting higher average correlation and energy-related metabolites showing lower correlation (Additional file 1: Fig. S1B). Detailed distributions and representative EBPs with high and low concordance are provided in Additional file 1.

When compared with previously published DNAm-based surrogates, including protein EpiScores[12], Nightingale metabolite surrogates [17], and GrimAge-derived protein components [4,18] EBPs demonstrated comparable or higher correlations with observed biomarkers for most overlapping targets (Additional file 1: Fig. S2).

### Comparison of EBP- and observed biomarker–disease associations in the development cohort (MGB-ABC)

To evaluate whether EBPs reproduce clinically meaningful biomarker–disease association patterns, we next compared EBP- and observed biomarker–disease association estimates. Across 12 incident and prevalent outcomes, EBP-derived and observed-biomarker effect estimates were directionally concordant for multiple associations (Figure 2, Additional file 2: Table S5). Notably, EBPs for glycemic traits and renal function markers recapitulated expected associations with type 2 diabetes and chronic kidney disease, respectively, while inflammatory and cardiometabolic proxies showed coherent patterns across cardiovascular and related outcomes, indicating concordance of association patterns.

**Figure 2.**
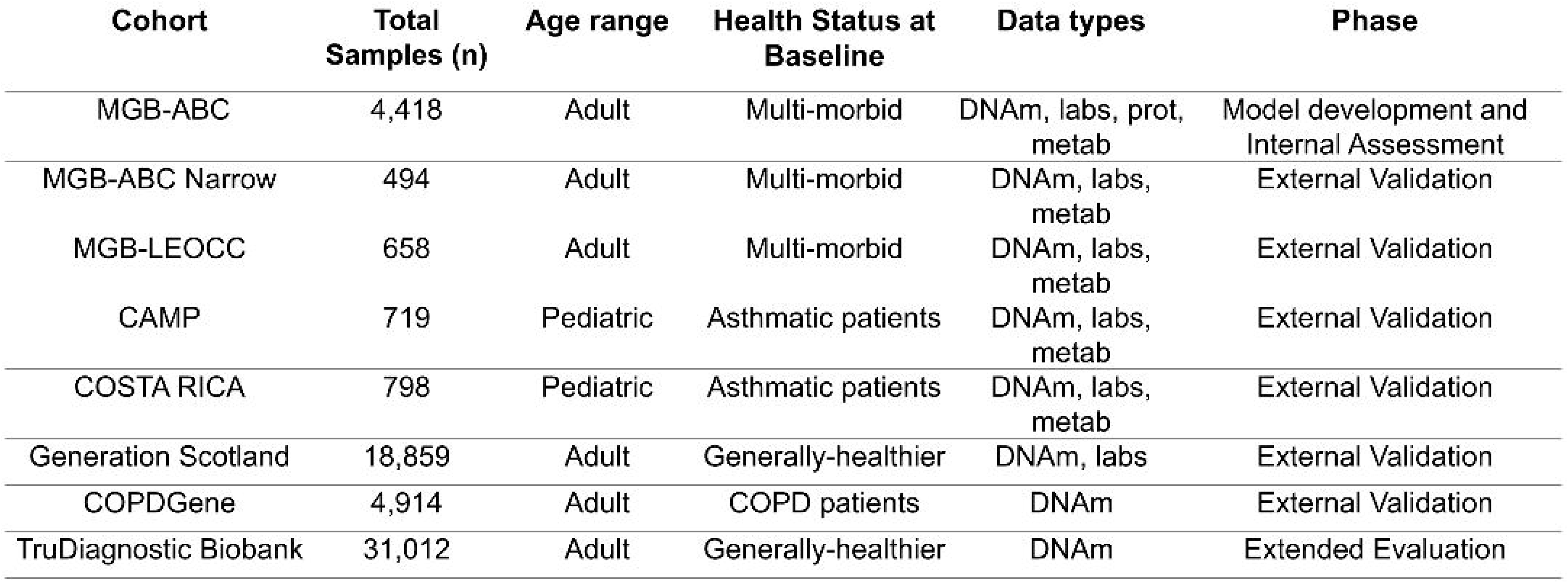
Alignment of observed and EBP-derived effect sizes across outcomes and biomarker types. Scatterplots compare observed hazard ratios (left) and odds ratios (right) with corresponding epigenetic biomarker (EBP) effect sizes for clinical lab measures, metabolites, and proteins. Each point represents a trait, colored by associated disease or outcome. The diagonal line indicates perfect concordance; selected traits are labeled.

We assessed associations with all-cause mortality and 11 chronic diseases in the MGB-ABC testing cohort using both time-to-event and cross-sectional models to derive hazard ratios and odds ratios, respectively (Additional file 2: Table S5). Table 1 summarizes the number of FDR-significant associations identified for EBPs and observed biomarkers. In incident analyses, 1,652 EBP–disease associations and 1,200 observed biomarker–disease associations were significant, with 532 shared between approaches. In prevalent analyses, 4,115 significant EBP associations and 4,380 observed associations were identified, of which 2,201 were shared.

**Table 1.**
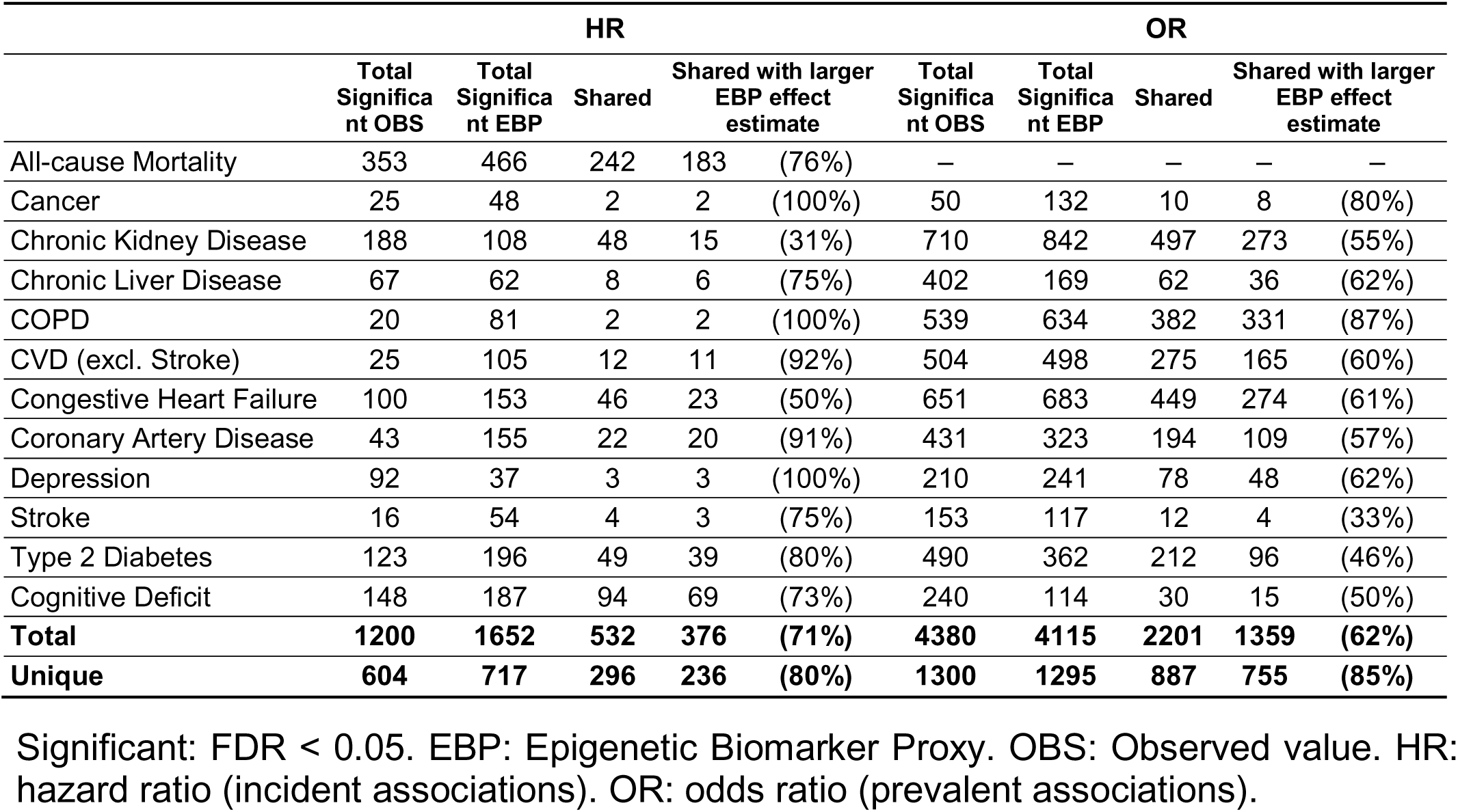
Summary of FDR-significant associations for EBPs and corresponding observed biomarkers across all-cause mortality and 11 chronic diseases. For associations significant in both analyses, the table also reports the number for which the EBP-derived effect estimate was larger in magnitude.

For associations significant in both approaches, the effect sizes from EBP were generally consistent with the observed biomarkers (Figure 2). Moreover, several of the EBP-derived effect estimates were larger in magnitude than those from observed biomarkers including 68% of clinical, 75% of metabolite, and 60% of protein associations in time-to-event models, and 70%, 51%, and 74%, respectively, in cross-sectional models (Additional file 2: Table S6). Importantly, the magnitude of these associations is descriptive in nature.

To further characterize disease-specific association patterns, we generated volcano plots comparing EBPs and observed biomarkers for each outcome (Additional file 1: Figs. S3–S14) and forest plots summarizing the strongest positive and negative associations per disease alongside corresponding observed biomarkers (Additional file 1: Figs. S15–S26).

Additionally, to assess whether EBPs follow the expected tendency of biomarkers to be associated with multiple diseases, we examined cross-disease association patterns. We found that multiple EBPs were associated with more than one chronic disease or with mortality at FDR < 0.05. For example, EBPs for clinical traits such as red cell distribution width, body mass index, glucose, and hematocrit—established markers of systemic health—were associated with multiple disease outcomes, including cardiovascular disease, type 2 diabetes, and cancer.

In time-to-event analyses (Figure 3A), 9 clinical, 22 metabolite, and 1 protein EBPs were significantly associated with seven or more health outcomes (FDR < 0.05), with all-cause mortality and cognitive deficit showing the highest degree of overlap. In cross-sectional analyses (Figure 3B), 6 clinical, 8 metabolite, and 12 protein EBPs were associated with nine or more health outcomes, with chronic kidney disease and congestive heart failure sharing the greatest number of multi-morbid EBPs. In contrast to incident associations, a larger number of protein EBPs were associated with multiple diseases in the prevalent setting, whereas cancer-related EBPs showed lower cross-disease overlap in both analyses, suggesting greater disease specificity. Together, these findings demonstrate both outcome-specific and cross-outcome EBP association patterns.

**Figure 3.**
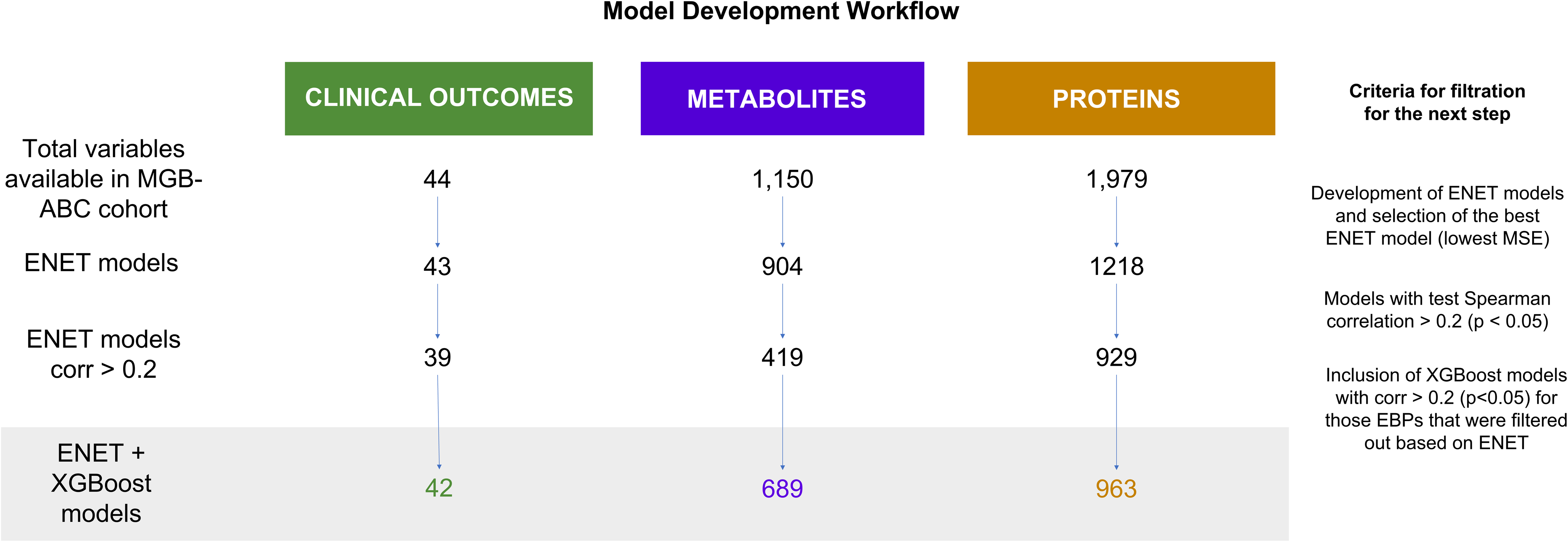
Heatmap showing significant EBPs for each disease and multi-disease EBPs. (A) Incident associations based on time-to-event analyses; EBPs significantly associated (FDR < 0.05) with seven or more diseases are shown. (B) Prevalent associations based on cross-sectional analyses; EBPs associated with nine or more diseases are shown. Horizontal bars indicate the number of significant EBPs per disease. EBPs are color-coded by class: clinical labs (green), metabolites (blue), and proteins (orange). In the heatmaps, square shading reflects the false discovery rate (FDR), with darker shades indicating stronger significance (lower FDR) and white indicating non-significance (FDR ≥ 0.05).

### Detection of clinical range deviations using EBPs

To evaluate the potential clinical utility of EBPs, we assessed their capacity to classify deviations from reference ranges (“in-range” vs. “out-of-range”) for clinical measures in the MGB-ABC test set. We focused on 35 clinical biomarkers with at least one out-of-range (flagged) value, and less than 100% flagged values (Figure 4, Additional file 2: Table S7). Across these markers, EBPs achieved a mean classification accuracy of 77.83% (median: 77.48%; range: 52.11%–95.08%), mean sensitivity of 62.03% (median: 62.46%; range: 35%–94.71%), and mean specificity of 78.2% (median: 83.64%; range: 14.29%–97.75%). Sensitivity increased with the prevalence of flagged values, while accuracy and specificity declined, consistent with trade-offs between false positives and false negatives (correlations: accuracy = –0.53; sensitivity = 0.77; specificity = –0.94). Notably, the EBPs for globulin, creatinine, lymphocyte counts, and hemoglobin all showed accuracies above 75 percent.

**Figure 4.**
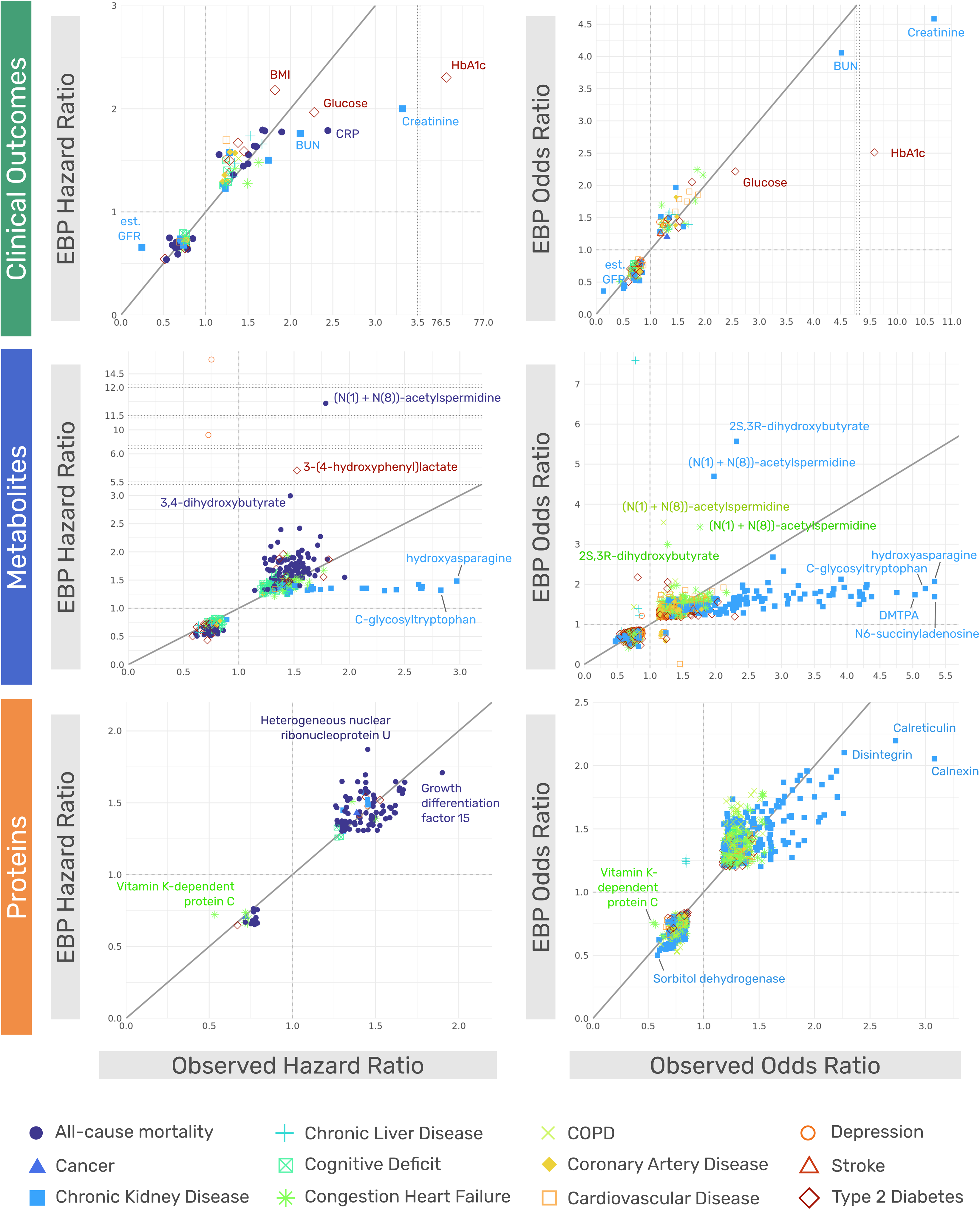
Performance of EBPs in identifying clinically abnormal lab values in the MGB-ABC test set. Accuracy (orange circles), sensitivity (green crosses), and specificity (blue Xs) of clinical EBPs in detecting out-of-range lab values based on standard reference intervals. Biomarkers are sorted by accuracy, and shading reflects the proportion of flagged (abnormal) observed values for each biomarker. The red dashed line indicates performance of 75%.

### External validation across independent cohorts

To evaluate the generalizability of EBP models across populations, we deployed the trained models to seven independent external cohorts spanning different age groups, disease distributions, and biospecimen sources (Additional file 2: Table S1). EBPs for clinical values, metabolites, and proteins were computed using DNAm data from each cohort. Validation analyses were performed according to the molecular and phenotypic data available in each cohort, enabling assessment of biomarker concordance, disease associations, and clinical range classification across diverse study designs.

### Replication of biomarker correlations

We evaluated EBP–biomarker correlations across five external cohorts with available clinical or molecular measurements, including MGB-ABC Narrow, MGB-LEOCC, CAMP, Costa Rica (CRA), and Generation Scotland (GS). The direction and relative pattern of correlations observed in the MGB-ABC testing set were generally preserved across validation cohorts, although the magnitude varied across cohorts (Spearman r = 0.31–0.96 (Spearman r = 0.31–0.96), with correlations above 0.50 in all cohorts except MGB-LEOCC (r = 0.31; Additional file 1: Fig. S27A).In several cohorts, including MGB-ABC Narrow and MGB-LEOCC, a majority of EBP–observation correlations were comparable to or higher than those observed in the MGB-ABC testing set, with similar trends also observed in CAMP, CRA, and GS (Additional file 1: Fig. S27B). Notably, robust replication was observed in the CAMP and CRA cohorts, which consist of pediatric participants and therefore differ substantially in demographic composition from the adult-based MGB development cohort.

Metabolite-level correlation patterns were also broadly replicated, although correlations were generally modest in magnitude. Overall concordance patterns remained consistent with those observed in the development cohort, with absolute correlations for some metabolites modestly attenuated in MGB-ABC Narrow and MGB-LEOCC (Additional file 1: Fig. S28). Pathway-level analyses also showed consistent replication across cohorts, with amino acid and lipid metabolites showing consistently strong concordance, and androgenic steroid EBPs exhibiting the highest agreement with observed values (Additional file 1: Figs. S29–S30).

### Replication of disease associations

We validated the incident and prevalent associations between the obtained EBPs and multiple diseases that were identified in our test set, using two external validation cohorts: Generation Scotland, COPDGene (Figure 5, Additional file 2: Table S5, Additional file 2: Table S8). Across all cohorts, we observed similar trends in effect sizes, particularly for the EBPs of clinical markers such as glucose, BMI, HbA1c, and creatinine, which remained aligned with their known disease associations. Notably, key metabolite EBPs—including N-acetylmethionine, HPMA, and (N¹,N)-acetylspermidine—also demonstrated consistent effect patterns across diseases, such as COPD and cardiovascular disease. Protein EBPs such as plasminogen activator inhibitor 1 and cold-inducible RNA-binding protein showed strong replication in Generation Scotland, while markers like Lamin A/C and SDF3 remained predictive in COPDGene.

**Figure 5.**
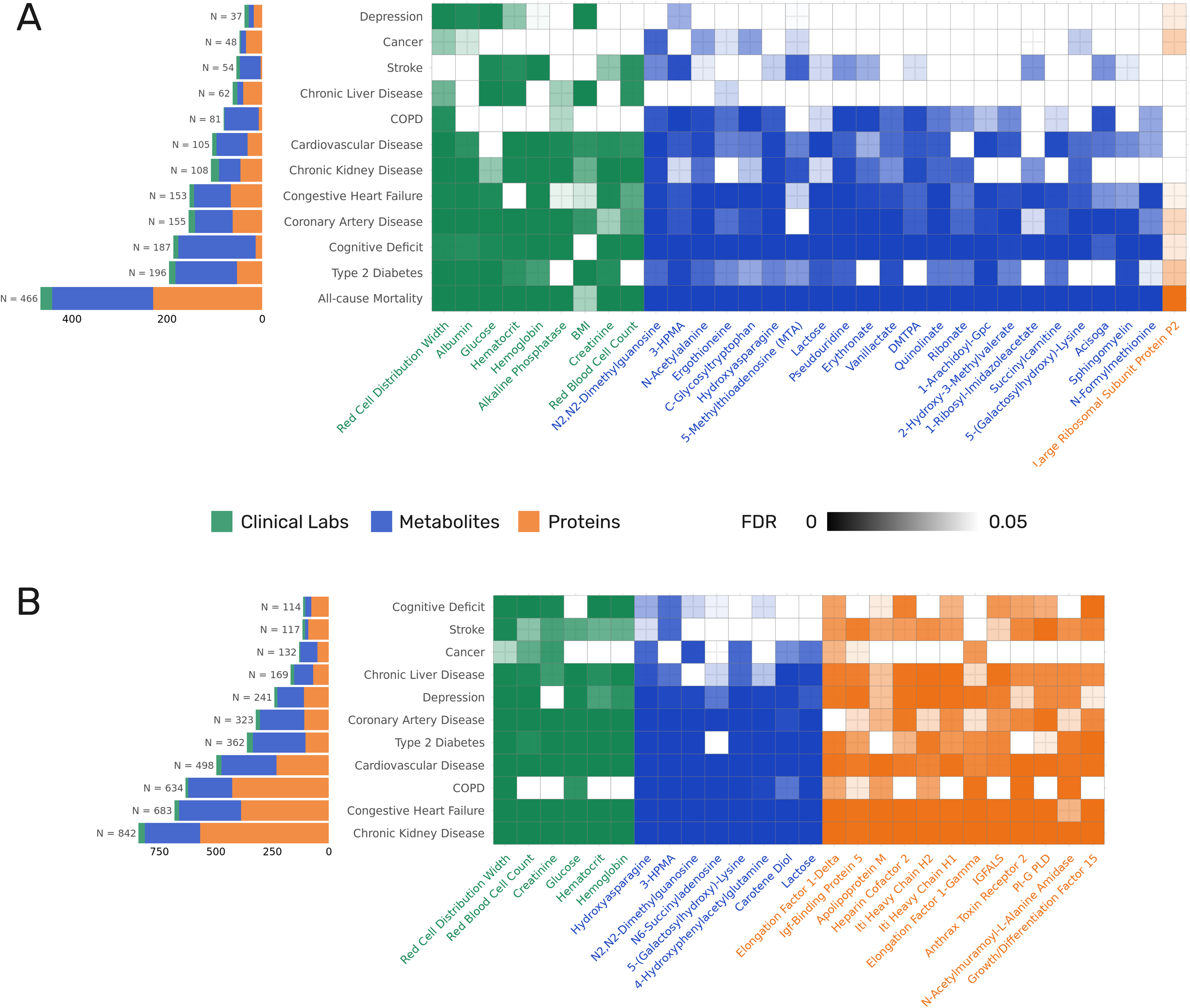
Validation of EBP-derived effect sizes in external cohorts. Scatterplots show hazard ratios (Generation Scotland, left) and odds ratios (COPDGene, right) for their corresponding epigenetic biomarker (EBP) predictions across clinical labs, metabolites, and proteins compared to the Hazard Ratios and Odd Ratio in the MGB-ABC test set. Each point represents a trait, colored by the associated disease. The diagonal line indicates perfect concordance; selected traits are labeled.

### Replication of clinical range deviations detection

We evaluated the ability of EBPs to classify deviations from reference ranges in validation cohorts with available clinical measurements (MGB-ABC Narrow and MGB-LEOCC), using the same methodology applied in the MGB-ABC test set. In the MGB-ABC Narrow cohort, EBPs achieved a mean classification accuracy for detecting out-of-range measurements of 80.3%, with a mean sensitivity of 65.5% and mean specificity of 79.5%; corresponding medians were 82.8%, 67.0%, and 87.5%, respectively (Additional file 2: Table S9). In the MGB-LEOCC cohort, the mean classification accuracy was 78.8%, with a mean sensitivity of 57.8% and mean specificity of 78.6%; median values were 81.9%, 58.6%, and 86.2%, respectively (Additional file 2: Table S10). These results were consistent with those observed in the MGB-ABC test set (mean and median classification accuracy of 77.8 and 77.5% respectively), characterized by consistently high specificity and more variable sensitivity across clinical markers (Additional file 1: Fig. S31). Together, these findings support the generalizability of MGB-trained EBP models for classifying out-of-range clinical measurements across diverse populations and clinical settings.

### External application and longitudinal behavior of EBPs

Having established that EBPs replicate biomarker correlations and disease associations across multiple independent cohorts with diverse demographic and clinical characteristics, we next evaluated their potential utility beyond cross-sectional validation.

We performed two analyses in an independent cohort. First, to assess whether DNAm-based EBPs reflect known biological and clinical patterns, we tested a pre-specified, clinician-curated set of EBP–trait associations in 31,012 samples from the TruDiagnostic cohort using self-reported lifestyle, disease, and supplement data. Rather than an agnostic screen, this hypothesis-driven design targeted associations with strong prior biological plausibility (10 lifestyle factors, 37 diseases, and 21 supplement categories), yielding 227 EBP–trait contrasts in the pooled, sex-adjusted analysis (Additional file 2: Table S11) and 836 contrasts in the sex- and array-stratified analysis (Additional file 2: Table S12). Associations were modeled with linear regression adjusted for biological sex, and p-values were corrected within each table using the Benjamini–Hochberg FDR. Consistent with the a priori design, a large fraction of contrasts remained significant after correction (124/227 [54.6%] in Additional file 2: Table S11 and 289/836 [34.6%] in Additional file 2: Table S12 at FDR < 0.05), as expected when candidate associations are pre-selected for plausibility rather than tested indiscriminately. The single strongest signal, the association between the urate EBP and biological sex, reflects the well-established sexual dimorphism of urate (higher in males) rather than uncontrolled confounding; accordingly, all remaining factor associations were adjusted for, or stratified by, sex.

Among the FDR-significant associations, several aligned with known biology. Gout was linked to elevated DNAm levels of uric acid pathway metabolites, including xanthine, urate, and allantoin, particularly in males (Figure 6A–D). Individuals with high blood pressure had higher DNAm phenylacetylglutamine and vanillactate levels (Figure 6E– H), consistent with cardiovascular risk profiles. NAD+ and Omega-3 supplementation also showed expected epigenetic shifts: NAD+ users had elevated DNAm 1-methylnicotinamide and N1-methyl-2-pyridone-5-carboxamide (Figure 6I–L, Additional file 1: Fig. S32A–F), while Omega-3 users showed increases in DNAm DHA, DPA, and EPA (Additional file 1: Fig. S32G–I).

**Figure 6.**
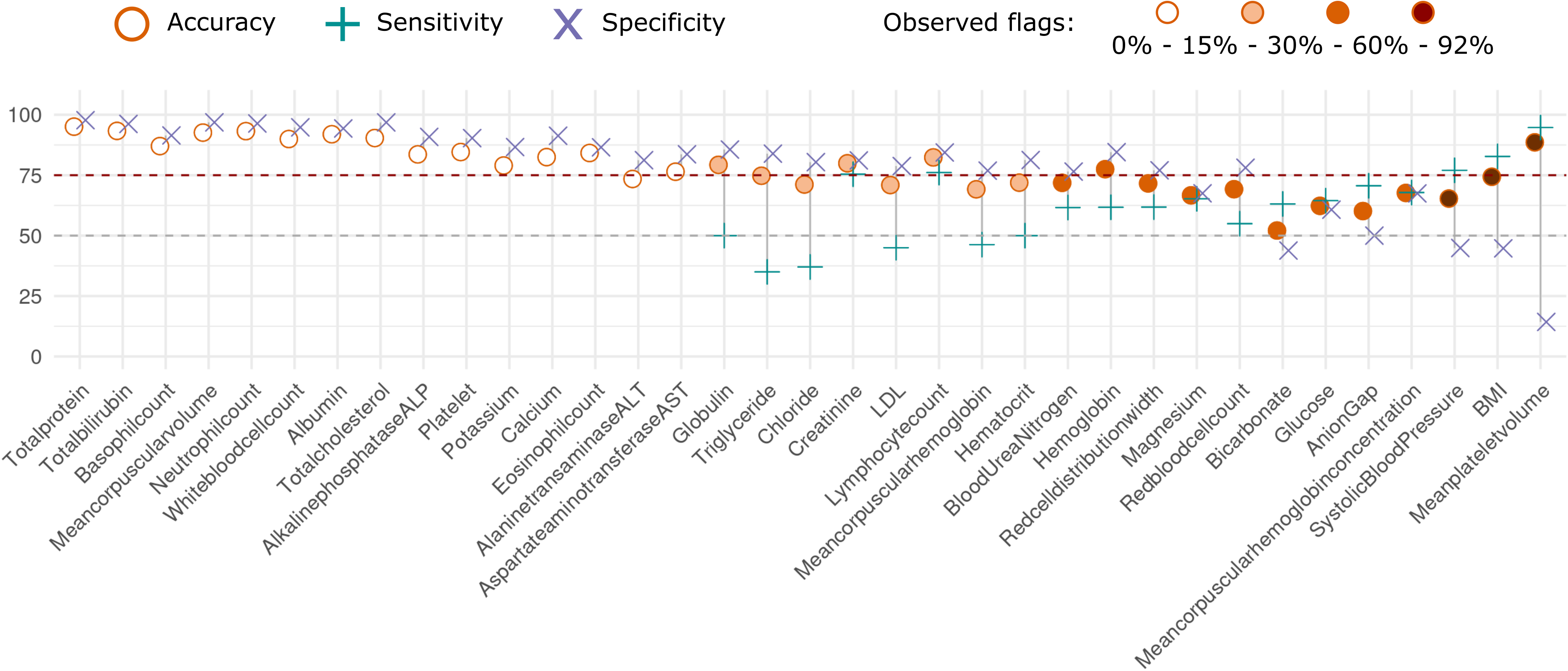
Validation analyses evaluating the changes in the EBP levels in different scenarios. (A-C) Association of Xanthine, Urate, and Allantoin EBPs with the presence or absence of gout. (D) Uric acid pathway. (E) Association of Phenylacetylglutamine EBP with high blood pressure. (F) Implication of Phenylacetylglutamine in the prevalence of high blood pressure. (G) Association of Vanillactate EBP with high blood pressure. (H) Implication of Vanillactate in the high blood pressure prevalence. (I-K) Association of Nicotinamide, 1-Methylnicotinamide, and N1-methyl-2-pyridone-5-carboxamide EBPs with NAD Supplementation. (L) NAD supplementation mode of action.

Finally, we evaluated the longitudinal stability of EBPs by comparing their trajectories to matched clinical measurements across multiple timepoints in a subset of ten individuals from the TruDiagnostic cohort. Each individual had at least four paired DNAm and clinical measurements, covering a panel of 33 biomarkers. After standardizing (mean-centering and normalizing) the observed and EBP-predicted values, we calculated their absolute difference at each timepoint. Then, we obtained the within-individual variance of these differences across timepoints. Finally, we standardized these variances relative to the pooled distribution across all individuals and biomarkers to obtain an instability z-score for each individual-biomarker. This provided a descriptive assessment of concordance and temporal instability between observed and EBP-predicted values.

Visual inspection of white blood cell counts (Additional file 1: Fig. S33), consistent with quantitative summaries, showed qualitatively similar EBP and observed trajectories for several biomarkers across time, with relatively low deviation across the five representative individuals shown. Among these, basophil, lymphocyte, and neutrophil EBPs showed particularly stable alignment with their clinical counterparts, while monocyte EBPs showed greater variability in select individuals. We also observed overall low trajectory variance z-scores in the difference between EBP and observed values across timepoints for most biomarkers (Additional file 1: Fig. S33).

Together, these results demonstrate that DNA methylation–based EBPs can be constructed at scale and highlight the reproducibility of the selected correlation, association, and classification results across diverse populations, disease contexts, and data platforms.

## Discussion

In this study, we developed EBPs for 1,694 circulating proteins, metabolites, and clinical measures and evaluated their clinical utility in relation to chronic diseases and mortality over up to 15 years. Our findings demonstrate that EBPs capture many of the significant incident and prevalent disease associations that were observed in the actual biomarkers. We found that when associations for both EBPs and their observed counterparts were statistically significant, EBPs often exhibited larger effect estimates than the corresponding observed biomarkers, which may reflect temporally integrated biological signals rather than superiority over direct measurements. In addition, EBPs of clinical biochemistries detect deviations from standard clinical ranges with moderate-to-high accuracy, findings validated in seven external cohorts covering 57,454 samples. In an exploratory analysis of ten individuals, EBPs also showed qualitatively consistent longitudinal tracking relative to observed clinical values. Together, these results highlight the potential utility of EBPs as scalable molecular proxies for population-level risk stratification and longitudinal phenotyping. Beyond serving as an initial screen for standard clinical biochemistries, EBPs extend molecular coverage to metabolic and inflammatory domains, providing complementary insight into underlying physiology. While further work is needed to refine and expand these models, EBPs represent a scalable framework for population-level disease risk stratification and longitudinal biological characterization.

A central conceptual insight emerging from this work is that epigenetic biomarker proxies do not merely approximate contemporaneous biomarker concentrations, but instead appear to encode temporally integrated biological information reflecting cumulative exposures, physiological regulation, and disease-related processes. This property may explain why EBPs often show modest cross-sectional correlations with observed biomarkers yet exhibit robust associations with long-term disease outcomes. This suggests that EBPs may function less as point-in-time molecular measurements and more as molecular summaries of sustained biological states, complementing traditional clinical assays that capture acute physiology.

We extended upon prior work by substantially expanding the scope of DNAm surrogates, increasing to >1,600 EBPs that capture clinical, proteomic, and metabolomic domains. We show that when both the observed biomarkers and EBPs were significantly associated with disease, in some cases, EBPs had higher disease effect size estimates than their measured counterparts. Previous work, ranging from studies of a singular protein up to 109 protein EpiScores, also observed stronger disease associations compared with the actual measured proteins and captured relevant inflammatory biology [11,12,17,19–21]. While such findings may initially appear counterintuitive, they are consistent with the hypothesis that EBPs capture biological signals integrated over longer timescales than single laboratory measurements. Another study creating 64 metabolite surrogates found that these improved the prediction of mortality, but did not evaluate their association across disease outcomes [17].

By extending these observations across clinical, proteomic, and metabolomic domains, our results demonstrate that the same principles apply across a broader range of physiological processes and disease outcomes. Together, these results suggest that EBPs may capture biologically relevant disease-associated signals across molecular domains. Importantly, these results were observed across seven independent cohorts representing a range of age distributions, disease burdens, and biospecimen sources. Across cohorts, many EBP–disease associations observed in the development dataset were replicated, including consistent patterns for chronic kidney disease, cardiovascular outcomes, and mortality, supporting the generalizability of the EBP framework beyond the original training population.

We next evaluated the potential utility of EBPs in clinically relevant contexts by assessing their ability to detect deviations from established reference ranges and to track longitudinal changes in clinical biomarkers. EBPs demonstrated moderate-to-high classification performance for identifying out-of-clinical-range laboratory values, characterized by generally good specificity and more variable sensitivity across biomarkers. Notably, EBPs for creatinine and lymphocyte count performed particularly well, highlighting the potential for EBPs to inform population-level screening and prioritization for immune system status and kidney-related risk in future studies [22]. Consistent with classification theory, we observed trade-offs between sensitivity and specificity that were influenced by the prevalence of out-of-range values, underscoring the need for further validation across cohorts with differing disease prevalence. In exploratory longitudinal analyses of ten individuals, many EBPs tracked observed biomarker trajectories with qualitatively good concordance, suggesting their potential utility for longitudinal measurements. Beyond laboratory-based validation, EBPs also reflected biologically coherent patterns associated with specific diseases and sustained exposures, including uric acid pathway metabolites in gout, NAD - related metabolites following supplementation, and omega-3 fatty acid–related EBPs in supplement users. Together, these observations suggest that EBPs can capture biologically meaningful variation associated with disease states and long-term exposures, extending their relevance beyond cross-sectional laboratory concordance.

It is important to highlight that current clinical biomarkers provide direct measures of physiology with specific clinical implications that EBPs may not have. Thus, EBPs should be considered complementary tools rather than replacements for traditional biomarkers. Nonetheless, EBPs have an enormous potential as inexpensive screening tools to simultaneously assess multiple health outcomes and identify individuals who may benefit from further diagnostic testing. They can also serve as risk stratification and/or diagnostic/prognostic biomarkers, particularly in situations where current biomarkers are limited or invasive. Further refinement and expansion of EBPs may capture molecular patient profiles that may offer guidance for clinicians as a precision medicine tool - where none currently exists - as an approach to guide overall patient care. EBPs may also be used as a complement to other risk scores. While methylation risk scores can provide important information about overall disease risk, EBPs can often supplement this information to provide actionable recommendations for heterogeneous diseases. For example, a cardiovascular methylation risk score identifies an individual at risk, but the incorporation of lipid, inflammatory, HBA1C EBPs provides more information that may help guide clinical treatment [23].

Despite growing interest in precision medicine, widespread clinical implementation of multi-omic profiling remains constrained by cost, infrastructure requirements, and challenges in integrating high-dimensional molecular data into existing clinical workflows. By leveraging DNAm as a single, scalable molecular readout, EBPs provide a framework for capturing information across multiple biological pathways from a single biosample. In this context, EBPs may support population-level risk stratification and prioritization for targeted diagnostic evaluation, particularly where repeated or comprehensive molecular testing is impractical.

Beyond clinical laboratories, we identified numerous protein and metabolite EBPs that exhibited strong correlations with observed values and were associated with multiple disease outcomes, suggesting relevance to overall health status. For example, glucuronate and pseudouridine EBPs were associated with cardiovascular disease, congestive heart failure, cognitive impairment, chronic kidney disease, and all-cause mortality, consistent with prior reports linking these metabolites to aging, longevity, and healthspan [24–26]. Similarly, protein EBPs such as large ribosomal subunit proteins and growth differentiation factor 15 demonstrated multimorbid associations aligned with previous studies identifying these proteins as systemic biomarkers [27–29]. With further validation, such EBPs may complement existing laboratory data to support monitoring of systemic health and physiological resilience.

Despite these strengths, several limitations warrant consideration. While this study represents the largest systematic effort to date to develop and evaluate DNAm–based biomarker proxies across clinical, proteomic, and metabolomic domains, some EBPs— particularly those derived from global proteomic and metabolomic platforms—lack absolute quantitative calibration, precluding direct definition of clinical reference ranges. Future studies incorporating targeted assays may address this limitation. In addition, model complexity relative to sample size likely contributed to some degree of overfitting in elastic net and XGBoost models. Nevertheless, validation against observed biomarkers and disease outcomes demonstrated that EBPs recapitulate expected biological associations, supporting their robustness as molecular surrogates. Expansion to larger biobank datasets and additional populations will further refine model stability and generalizability. Future work may also extend EBPs to biomarkers that are currently difficult to measure or exhibit lower concordance with DNAm.

## Conclusions

In summary, this study demonstrates that EBPs can be constructed at scale to capture clinically relevant biological variation across diverse molecular domains and disease contexts. By integrating information from clinical laboratory measures, proteins, and metabolites into a single molecular framework, EBPs provide a systematic approach for linking epigenetic variation with longitudinal health outcomes. While further validation and refinement are required, particularly for individual-level clinical use, these findings establish EBPs as a generalizable platform for population-level risk stratification, biological characterization, and future translational research.

## Methods

### Discovery and Validation Cohorts

#### Development Cohort: Massachusetts General Brigham Aging Biobank Cohort (MGB-ABC)

All model development, feature selection, and hyperparameter tuning were performed exclusively in the MGB-ABC cohort. External cohorts were used solely for independent validation and downstream analyses. The Massachusetts General Brigham (MGB) Biobank is a large institutional resource that includes research data and biospecimens—plasma, serum, and DNA—from over 100,000 consented patients recruited through routine clinical care at Massachusetts General Hospital (MGH) and Brigham and Women’s Hospital (BWH). All participants are linked to longitudinal electronic medical record (EMR) data, including demographics, diagnoses, procedures, prescriptions, laboratory values, and clinical notes, along with health surveys capturing lifestyle, environmental exposures, and family history. The MGB-Aging Biobank Cohort (MGB-ABC) consists of 4,418 participants randomly selected from the larger Biobank to create a proportionately representative aging cohort based on age, sex, and BMI. Blood samples were collected either during clinical visits or dedicated research draws and processed for genomic, epigenetic, proteomic, and metabolomic profiling. The development cohort includes 1,148 individuals from the ABC cohort with a metabolomic profile, and 1,228 with a proteomics profile. Each sample is linked to corresponding clinical data, and a subset of participants also completed structured questionnaires. Metabolomic data were available for select individuals and were accessed through the MGB Biobank Portal for this study.

The Phenotype Discovery Center (PDC) of MGB integrates various data sources, including Research Patient Data Registry (RPDR), health information surveys, and genotype results, into the Biobank Portal. This portal combines specimen data with EMR data, creating a comprehensive SQL Server database with a user-friendly web-based application[30]. Researchers can conduct queries, visualize longitudinal data with timestamps, employ established algorithms to define phenotypes, utilize automated natural language processing (NLP) tools for analyzing EMR data using the Informatics for Integrating Biology and the Bedside (i2b2) toolkit [31].They can also request samples from cases and controls. Data in the Biobank Portal database includes narrative data from clinic notes, text reports (cardiology, pathology, radiology, operative, discharge summaries), codified data (e.g., demographics, diagnoses, procedures, labs and medications), and patient-reported data from the health information survey on exposures and family history. Validated phenotypes are available in the Biobank Portal user interface for genotyped Biobank participants, and other relevant measures, such as lung function, were extracted using a self-developed algorithm incorporating NLP.

There were a few clinical biomarkers that were available only for a subset of users. For those specific ones, we supplemented the dataset with 76 individuals recruited through a precision medicine and wellness program. These samples were integrated into the development dataset before train-test split and only baseline data was used to develop the clinical EBPs.

#### MGB-ABC Narrow Cohort

The MGB-ABC Narrow cohort includes 494 individuals with a metabolomics profile and a selection of 42 clinical tests from the MGB Biobank, selected to optimize clinical data with the same criteria as the MGB-ABC Cohort, without overlap with our development set. The cohort is 53% male, predominantly white (85.22%), and the mean age is 61.9 (range 20.8 to 96.3).

#### MGB-LEOCC Cohort

The MGB-Longitudinal Electronic Omics of COVID-19 Cohort (MGB-LEOCC) was created from the MGB Biobank and comprises all individuals with available plasma samples and a positive COVID-19 diagnosis in the EMR from the start of the COVID-19 pandemic (01/01/2020) through 10/27/2020 [32]. A total of 658 participants with banked plasma samples contributed prior to COVID-19 infection were included in our study. The cohort is 41% male, predominantly white (60.18%), and the mean age is 51.5 (range 7.9 to 94.3).

#### Childhood Asthma Management Program (CAMP) Cohort

The Childhood Asthma Management Program (CAMP) has been extensively described elsewhere [33]. In brief, CAMP was a longitudinal cohort that recruited children with asthma and then followed them up for an average of 16 years. In total, 719 subjects with DNAm and clinical laboratory tests data were included in this analysis [34]. DNA methylation was measured at the baseline timepoint. The mean age at baseline was 8.8 years. The original study population was predominantly male (63–64% across the three time-points) and predominantly non-Hispanic white (70–72%).

#### Genetic Epidemiology of Asthma in Costa Rica Study (CRA)

The Genetic Epidemiology of Asthma in Costa Rica Study (CRA) is a family-based study designed to investigate the genetic, environmental, and immunologic determinants of asthma in children. Recruitment was conducted in the Central Valley of Costa Rica, a region with high asthma prevalence, and included children aged 6–14 years with physician-diagnosed asthma along with their parents. Participants completed detailed respiratory and environmental questionnaires, underwent pulmonary function testing, and provided blood samples for DNA extraction and biomarker analyses. Compared to other cohorts in this study, the Costa Rica cohort represents a younger population with enriched familial aggregation of asthma and allergen sensitization. All study procedures were reviewed and approved by the Institutional Review Boards of the Hospital Nacional de Niños in San José, Costa Rica, and collaborating U.S. institutions, and informed consent was obtained from all participants or their guardians. Samples from 798 individuals were used for this analysis with metabolomic and DNAm profiling [34]. 59% were male and the mean age at baseline was 9.22 (ranging from 5.44 to 15.16).

#### Generation Scotland Biobank Cohort

The Generation Scotland (GenS) cohort is a large, family-based population study designed to investigate genetic, epigenetic, and environmental determinants of health and disease across Scotland. Between 2006 and 2011, approximately 24,000 adult participants were recruited through general medical practices across the country, with many individuals enrolled as part of extended family groups to facilitate heritability and familial analyses. Participants completed detailed health and lifestyle questionnaires and attended in-person clinic visits where anthropometric, cognitive, and physiological measurements were obtained. Blood samples were collected for biochemical assays, DNA extraction, and biobanking. DNAm data were generated from peripheral blood samples for a subset of participants and used in the present analyses. Compared to participants from the MGB and TruDiagnostic cohorts, GS participants represent a population-based sample with broad age distribution and health status reflective of the general Scottish population. All participants provided written informed consent, and study procedures were approved by the relevant institutional and National Health Service (NHS) research ethics committees. For this study, we used 18,859 participants with an average age of 47.1. All participants were white and 41.2% were male.

#### COPDGene

The COPDGene cohort is part of a large, multicenter, prospective study designed to identify genetic and environmental determinants of chronic obstructive pulmonary disease (COPD). Between 2007 and 2011, the study enrolled over 10,000 current and former smokers (⅔ non-Hispanic white, ⅓ African American) aged 45–80 years with a minimum of 10 pack-years of smoking history across 21 clinical centers in the United States. Participants completed detailed questionnaires on medical history, smoking behavior, and lifestyle, and underwent standardized spirometry. Blood samples were collected for DNA extraction. DNA methylation data were generated for a subset of Phase 1 (baseline) participants. The study was approved by institutional review boards at all 21 participating centers, and all participants provided written informed consent prior to enrollment. Data from Phase 1 included a total of 4,914 individuals used for our validation, comprising 54% of males and aged between 51.7 and 66.2.

#### TruDiagnostic Biobank Cohort

The TruDiagnostic Biobank cohort includes 31,012 samples from 26,155 participants, recruited primarily from the United States between October 2020 and September 2024, who completed the commercial TruDiagnostic TruAge test and had their DNAm data generated. Compared to participants from the MGB cohort, these participants were in better health. The majority of the samples were collected under a healthcare provider’s recommendation or guidance while less than 5% were in a direct-to-consumer setting, possibly introducing a self-selection bias as most participants were likely to seek preventative medicine and have fewer comorbidities than normal patient populations. Participants were also asked to complete a survey about personal information, medical history, social history, lifestyle, and family history. The study involving human participants was reviewed and approved by the IRCM Institutional Review Board (IRB) and all participants provided written informed consent to take part in the study. Among the participants from this cohort, 58.5% were male and ages ranged from 4.4 to 100 years old.

### Profiling

#### Methylation profiling

The DNA methylation data for this analysis was previously described [15]. Briefly, whole blood samples were processed using the Illumina Infinium® MethylationEPIC 850K BeadChip, which offers extensive genome-wide coverage across over 850,000 methylation sites, encompassing key regulatory and functional genomic regions. In the MGB cohorts, sample preparation and bisulfite conversion were conducted at TruDiagnostic Inc. under standardized conditions. Quality control and preprocessing followed established pipelines (minfi and ENmix), with stringent criteria applied to retain 4,418 high-quality samples and 863,430 reliable probes. Data normalization employed Funnorm and RCP methods to minimize inter-sample variability [35].

In the TruDiagnostic cohort, the same process was followed for processing the samples, but half of them were analyzed using the Infinium® MethylationEPIC v2 BeadChip instead of the Infinium® MethylationEPIC 850K BeadChip. To enable application of EBPs to samples profiled on the Infinium® MethylationEPIC v2 array, CpG sites absent from EPIC v1 were imputed using age-stratified mean methylation levels computed in 5-year windows. This approach preserves age-dependent methylation structure while avoiding model retraining and has minimal impact on downstream association analyses, as validation analyses focus on relative concordance rather than absolute prediction error.

#### Metabolomic Profiling

Untargeted global plasma metabolomics data were previously generated as previously described [15]. Briefly, profiling was conducted by Metabolon Inc. using established quality control procedures, with coefficients of variation assessed through blinded QC samples to control batch effects. Metabolite extraction and profiling followed protocols as previously described, utilizing four distinct LC-MS methods tailored to detect complementary metabolite classes: (1) amines and polar metabolites in positive ion mode, (2) central and polar metabolites in negative ion mode, (3) polar and non-polar lipids, and (4) free fatty acids, bile acids, and intermediate polarity metabolites [36,37].

Metabolite identification was facilitated through an automated comparison of ion features against a reference library of ∼8,000 standards, supported by manual curation for quality assurance. Quantification involved area-under-the-curve measurements normalized per run-day to address instrument tuning variability, setting median values to 1.0. Metabolite identification adhered to stringent matching criteria, with new spectral entries created for unnamed compounds, identified through consistent recurrence patterns.

Data quality control and processing applied stringent thresholds, excluding features with signal-to-noise ratios <10 or >10% missing values, with remaining gaps imputed at half the minimum peak intensity. Features with a CV >25% in pooled samples were excluded for technical consistency. The final dataset comprised 1,150 metabolites from 1,148 samples, log-transformed for normalization, and pareto scaled to harmonize measurement scales across the metabolome.

#### Proteomic Profiling

The proteomic data used in this study were largely the same as described previously [15]. In summary, relative protein levels were quantified from 2,000 samples (including 1,600 from the MGB-ABC cohort and 400 process controls) using the Proteograph Product Suite (Seer, Inc.) in conjunction with LC-MS. We included 400 process-control runs (∼20% of total Proteograph runs), consistent with Seer’s guidance for maintaining robust QC coverage across large, multi-batch proteomics studies. Controls were distributed across the full study to provide continuous monitoring of assay performance and instrument stability. Proteins were captured using five proprietary nanoparticles on the Seer SP100 Proteograph, enabling selective binding based on physicochemical properties. Captured proteins were subsequently digested with trypsin, and relative quantification was performed using the data-independent acquisition (DIA) method integrated within the Proteograph Analysis Software (PAS). This produced estimations of a total of 28,490 peptides across blood samples (average 15,239), and 10,265 (average 4,281) across the controls.

The proteomic normalization procedures employed in this study were consistent with those previously detailed in the Tarkin study [38]. Proteomics processing involved the Seer Proteograph workflow for plasma sample preparation and peptide generation, followed by analysis on a Bruker timsTOF Pro 2 mass spectrometer using the dia-PASEF method. Peptide and protein intensities were derived through DIA-NN (v1.8.1) with a library-free search using UniProt (UP000005640_9606) and processed with the match between runs (MBR) option. Libraries were constructed to exclude or include common protein-altering variant (PAV) peptides, detailed in prior studies. DIA-NN’s normalized intensities (PG.Normalised) served as the protein readouts. From the total 28,317 protein groups quantified across 5 nanoparticles, 6,063 were present in at least 20% of samples. For proteins detected across multiple nanoparticle runs, the run with the highest total intensity was retained.

In order to adjust by genetic compounds, we subset the individuals with genetic and proteomic data. Imputed genotype data were filtered for biallelic variants using bcftools (v1.16) and converted to plink format, with subsequent filtering parameters set as –geno 0.1, –mac 10, –maf 0.05, and –hwe 1E-15. A final set of 1,260 samples was retained with proteomic data matched to 5,461,287 genetic variants. Principal components were computed using plink2 with the –pca option.

Protein data underwent log10 transformation, residualization by age, sex, and the first ten genotype principal components from the associated genotype data from the same sample, followed by inverse-normal scaling. This normalization approach supports robust proteomic analyses, building on established high-dimensional omics workflows. The final processed dataset represented 1,979 proteins.

### EBP development

#### Feature selection

To develop DNA methylation-based predictors for specific protein levels, metabolite concentrations, and clinical lab test values, we used the normalized DNAm dataset, transposing it so that CpG sites were considered features. In addition, sex was encoded as a binary feature (males and females), and was included as a penalized covariate, along with chronological age. Prior to training, we first filtered CpG site features based on mutual information to retain those most informative for each target outcome. The feature selection process began with discretizing features in the complete dataset by dividing each continuous methylation variable into 15 bins, thus converting it into discrete intervals. Mutual information scores between each CpG feature and the target outcome (e.g., specific metabolite levels) were then computed using the mutinformation function in the R package infotheo. To maintain specificity, mutual information was calculated between each feature and the target while excluding the target variable itself from the calculations. To focus on highly informative features, we applied a 90th percentile threshold, selecting only those CpG sites with mutual information scores above this cutoff. This percentile-based filtering ensured the retention of only the most relevant features, and filtered scores were saved separately for each target variable. The resulting dataset, containing the reduced CpG features, was combined with the target outcome to create the dataset. From this dataset, samples were split at random into an 85-15 train test ratio; the same train and test datasets were used through the whole training process.

#### Elastic Net Training

For training, we selected samples with quantified target outcomes and available chronological age and sex data. Models were generated using the glmnet package in R (version 4.1-8). Gaussian penalized regression models were trained on CpG site data and metabolite, protein, or clinical outcome levels using four alpha values (0.01, 0.1, 0.5, and 1) to assess the impact of varying the L1-L2 penalty balance. Hyperparameters were tuned through 10-fold cross-validation with cv.glmnet to select the optimal lambda for each alpha, minimizing mean squared error (MSE). All CpG sites and covariates were penalized, and features with at least five non-zero coefficients were retained in the final predictor models. Performance metrics—including MSE, mean absolute error (MAE), and Spearman correlation—were computed on test datasets to assess model fidelity. If more than one elastic net model was retained for a unique biomarker, we selected the one with the lowest MSE. EBPs failing to achieve a Spearman correlation ≥ 0.2 (p < 0.05) in the held-out test set were considered insufficiently predictive and evaluated using an alternative nonlinear modeling approach.

#### XGBoost Training

For targets not adequately captured by elastic net models, XGBoost was applied as a secondary modeling strategy. XGBoost models were trained and evaluated in a high-throughput R pipeline (version 4.1.1) using the data.table, xgboost, and caret packages, with the goal of maximizing predictive accuracy through optimized hyperparameters. Pre-filtered mutual information (MI)-based datasets were generated. Hyperparameter tuning was conducted using a grid search on a parameter grid with nrounds set to 10, eta values of 0.05 and 0.2, alpha values of 0.1, 0.5, and 1, and lambda set to 1. The caret package’s train function facilitated 3-fold cross-validation with parallel processing for efficiency. The optimal parameters identified from this grid search were applied to train the final XGBoost model, increasing nrounds to 1000 and incorporating an early_stopping_rounds parameter of 50 to prevent overfitting. This feature selection process improved model performance by reducing noise and enhancing signal quality, enabling the models to focus on CpG sites with the highest predictive potential for each target outcome. Model performance on the test data was assessed via mean squared error (MSE), mean absolute error (MAE), and Spearman correlation, with SHAP values calculated to capture CpG site contributions. We only selected those EBPs with Spearman test correlation higher than 0.2 (p<0.05) and without elastic net model retained.

### Statistical analyses

#### Evaluation of EBP performance

To evaluate the performance of the EBPs, we calculated the Spearman correlation between the EBPs and the observed clinical, metabolite, and protein levels. We also compared the EBPs with previously developed DNAm surrogates, including 109 Protein EpiScores [12], 64 Nightingale metabolite surrogates[17], and 12 principal-component (PC) based surrogate proteins [4,18]. Of these, there was an overlap between EBPs and 18 Episcores, 6 Nightingale metabolites, and 5 common PC-based protein surrogates. Using the testing dataset, we calculated the correlations between all DNAm surrogates and the observed values.

#### Association of EBPs to disease outcomes

To evaluate the clinical utility of EBPs, we analyzed their associations with diseases and compared these associations to those of the observed targeted measures. Diagnosis information, sourced from EMR data, allowed us to curate major chronic diseases prevalent in the MGB-ABC cohort using ICD-9/10-CM codes. For each disease, we identified both prevalent and incident cases by extracting the date of the first diagnosis. Patients diagnosed with a specific disease before plasma collection were classified as prevalent cases and included in a cross-sectional analysis. In this analysis, we estimated the odds ratio (OR) of each EBP or observed measure using logistic regression. Conversely, patients diagnosed after plasma collection were classified as incident cases and included in a prospective analysis, where we estimated the hazard ratio (HR) using a Cox proportional hazards model. All models were adjusted for age and sex to account for potential confounding factors. Additionally, we calculated the false discovery rate (FDR) to correct for inflated type I errors due to multiple comparisons. We applied the Benjamini-Hochberg procedure separately within each combination of molecular class (clinical, metabolite, protein) and effect-size type (HR, OR), pooling across all EBPs and all 12 outcomes within that stratum (six FDR families in total). Effect estimates were interpreted as measures of association rather than diagnostic performance, and differences between EBPs and observed biomarkers were not assumed to reflect superiority of one measurement modality over another.

#### Accuracy assessment of clinical EBP to detect deviations from recommended clinical ranges

The observed measurements, and EBP values of each biomarker were mean centered and normalized by dividing the individual values by the cohort’s standard deviation. To transform the reference ranges accordingly, we subtracted from each one the mean of the corresponding observed value, and then divided the result by the observed standard deviation. To assess the accuracy of EBPs in detecting clinical deviations, we first compared the observed measurements of the clinical biomarkers with more than 5 samples to a general clinical reference (Additional file 2: Table S13), and classified them as either within range or outside of range. For those biomarkers with more than 0% and less than 100% flags, we also compared the EBPs corresponding to each of the clinical values to the same reference. The cases when both the EBP and the observed value are within the reference ranges (true positive), or outside of the reference ranges (true negative) are considered correct detections. EBPs outside of the reference range that correspond to within range observed values are called false positives, while EBPs within the reference range corresponding to observed values outside of range are considered false negatives. The accuracy is calculated as the percentage of correct detections with respect to the total number of measurements. The specificity is calculated as the number of true negatives divided by the total number of in range observed measurements, and the sensitivity is calculated as the number of true positives divided by the total number of out of range observed measurements. To ensure statistical reliability of the specificity estimate, it was calculated on biomarkers with more than 15% flags only.

#### Metadata analysis

The TruDiagnostic Cohort represents individuals who have performed TruDiagnostic testing commercially in the past and consented to participate in research. Due to the high cost of this testing and the reporting offering (Biological Aging), participants within this cohort are generally self-selected to be more affluent and healthier than the general population. When completing the test, users will fill out basic demographic, lifestyle, medication, and disease surveys. To analyze the association of lifestyle, and demographic features with the EBPs, we used a pre-specified, hypothesis-driven design rather than an agnostic proxy-wide screen: a team of clinicians curated a focused list of EBP–trait pairs selected a priori for their strong prior biological plausibility (10 lifestyle factors, 37 diseases, and 21 supplement categories). Each association was evaluated with linear regression. In the pooled analysis (Additional file 2: Table S11), models were of the form EBP ∼ trait + biological sex (i.e., adjusted for biological sex; the sex term was omitted only when the trait itself was sex or when a single sex was represented). In addition, we repeated each association stratified by biological sex and DNAm array (EBP ∼ trait within each stratum; Additional file 2: Table S12). For each table, raw p-values were corrected for multiple comparisons across all factor–EBP contrasts in that table using the Benjamini–Hochberg false discovery rate procedure; both nominal and FDR-adjusted p-values are reported in Additional file 2: Tables S11 and S12. Because the tested associations were selected a priori for biological plausibility, a comparatively high proportion of significant results is expected and does not reflect uncontrolled false-positive inflation.

#### Longitudinal analysis

To explore longitudinal behavior of EBPs, we performed an exploratory analysis in 10 individuals with DNAm data and lab-based clinical biomarkers in at least four timepoints from the TruDiagnostic cohort. The observed measurements, and EBP values of each biomarker were mean centered and normalized by dividing the result by the cohort’s standard deviation. For each individual–biomarker pair, we calculated the variance of these absolute differences across the available timepoints. Finally, the variance estimates were pooled across all biomarkers and individuals in order to obtain an instability z-score for each individual–biomarker pair (subtracting the mean of the pooled variance estimates and dividing by their standard deviation).

## Declarations

### Ethics approval and consent to participate

This study was conducted in accordance with all applicable ethical guidelines and regulations. Ethical approval was obtained from the Institutional Review Board (IRB)/Ethics Committee at BWH (2014P001109), and informed consent was obtained from all participants prior to their inclusion in the study. Data were anonymized to protect participant privacy, and confidentiality was strictly maintained throughout the study. All human subject research adhered to the principles outlined in the Declaration of Helsinki, ensuring respect for individuals, beneficence, and justice.

### Consent for publication

Not applicable.

### Competing interests

NCG, LBD, IMG, RD, DP, AC, SV, SH, TM, RS, and VBD are employees of TruDiagnostic. JLS is a scientific advisor to TruDiagnostic, Precision Inc and Ahara Inc. This work was completed under a sponsored research agreement between Brigham Women’s Hospital and TruDiagnostic. This work was funded in part by TruDiagnostic. JLS, QC, VBD, RS have filed patents on work from this publication. DLD has received support from Bayer through the Pulmonary Drug Discovery Lab and from Astra Zeneca as a member of Medical Advisory Board. DLD has received grant support from the Alpha-1 Foundation.

## Funding

Efforts for QC, YC, TG, and JLS are supported by R01HL123915 and R01HL155742. Efforts for AA, JLS, and KM are supported by OT2HL161841 from the NIH/NHLBI. Efforts for YC and JLS are supported by R01HL141826 from the NIH/NHLBI. Efforts for JLS are supported by R01HL169300 from the NIH/NHLBI. Efforts for JLS are supported by U19AI168643 from the NIH/NIAID. KS is supported by the Biomedical Research Program at Weill Cornell Medicine in Qatar, a program funded by the Qatar Foundation, and also supported by Qatar National Research Fund (QNRF) grant ARG01-0420-230007.

DNA methylation profiling of the Generation Scotland samples was carried out by the Genetics Core Laboratory at the Edinburgh Clinical Research Facility, University of Edinburgh and was funded by the Medical Research Council UK and Wellcome Trust [104036/Z/14/Z and 220857/Z/20/Z]. Core support was received from the Chief Scientist Office of the Scottish Government Health Directorates [CZD/16/6] and the Scottish Funding Council [HR03006].

DNA methylation profiling for the Childhood Asthma Management Program (CAMP), the Genetic Epidemiology of Asthma in Costa Rica Study (CRA) and the Genetic Epidemiology of COPD (COPDGene) was performed through the Trans-Omics for Precision Medicine Program (TOPMed). All TOPMed data is person-sensitive, however it can be requested for access and can be made available through the TOPMed consortium after careful review and approval by the TOPMed Data Access Committee (https://topmed.nhlbi.nih.gov/). Participant consent and Data Use limitations differ within and across TOPMed studies and should be requested individually. Additional documentation, such as local IRB approval and/or letters of collaboration with the primary study PI(s) may be required. The CAMP data is available at the database of Genotypes and Phenotypes (dbGaP) repository (phs001726. v2. p1); the CRA data are available at dbGaP repository (phs000988. v5. p1); the COPDGene data are available at dbGAP repository (phs000951.v6.p5).

## Authors’ contributions

Data integrity: QC and VBD Conceived and designed the study - JLS, RS, VBD Performed analysis of physical samples for TruD cohort - TM Data processing and normalization - QC, VBD, KS, DLD, YH Algorithm development - QC, VBD, DP, SV, SH Statistical analysis and validation - QC, VBD, AA, NCG Analyses and visualizations - NCG, LBD, AA, RD, AC, TG, DLD, YH Drafted and edited manuscript - all authors

All authors read and approved the final manuscript.

## Data Availability

Requests for raw data, analyzed data and materials used to generate the results in this study will be reviewed by the contact PIs (Jessica Lasky-Su or Varun Dwaraka) to determine if the request is subject to intellectual property or confidentiality obligations. Datasets from electronic medical health records are not publicly available due to reasons of sensitivity. Restrictions apply to the use of these data due to privacy. Data that can be shared for research purposes upon request via a Data Use Agreement with the corresponding authors (Jessica Lasky-Su or Varun Dwaraka) by email. The code used in this study is publicly available on GitHub at https://github.com/LaskySuLab/EBPs

## Acknowledgements

We are grateful to all the people who have taken part in Generation Scotland, the general practitioners and the Scottish School of Primary Care for their help in recruiting them, the scientific steering committee and the whole GS team, which includes interviewers, computer and laboratory technicians, clerical workers, research scientists, volunteers, managers, receptionists, healthcare assistants and nurses.

COPDGene work was supported by NHLBI grants U01 HL089897 and U01 HL089856 and by NIH contract 75N92023D00011. The COPDGene study (NCT00608764) has also been supported by the COPD Foundation through contributions made to an Industry Advisory Committee that has included AstraZeneca, Bayer Pharmaceuticals, Boehringer-Ingelheim, Genentech, GlaxoSmithKline, Novartis, Pfizer, and Sunovion.

Molecular data for the Trans-Omics in Precision Medicine (TOPMed) program was supported by the National Heart, Lung and Blood Institute (NHLBI). Methylomics for “NHLBI TOPMed: COPDGene” (phs000951) was performed at Northwest Genomics Center (HHSN268201600032I). Methylomics for “NHLBI TOPMed: CRA_CAMP” (phs000951) was performed at Keck Molecular Genomics Core Facility (HHSN268201600038I). Core support including centralized genomic read mapping and genotype calling, along with variant quality metrics and filtering were provided by the TOPMed Informatics Research Center (3R01HL-117626-02S1; contract HHSN268201800002I). Core support including phenotype harmonization, data management, sample-identity QC, and general program coordination were provided by the TOPMed Data Coordinating Center (R01HL-120393; U01HL-120393; contract HHSN268201800001I). We gratefully acknowledge the studies and participants who provided biological samples and data for TOPMed.

## Additional files

**Additional file 1.** Supplementary Figures S1–S33.

**Additional file 2.** Tables S1–S13.xlsx. Supplementary Tables S1–S13. Excel workbook containing one worksheet per supplementary table, plus an index worksheet listing the title and description of each table. Full legends are repeated in the first row of every worksheet.

---

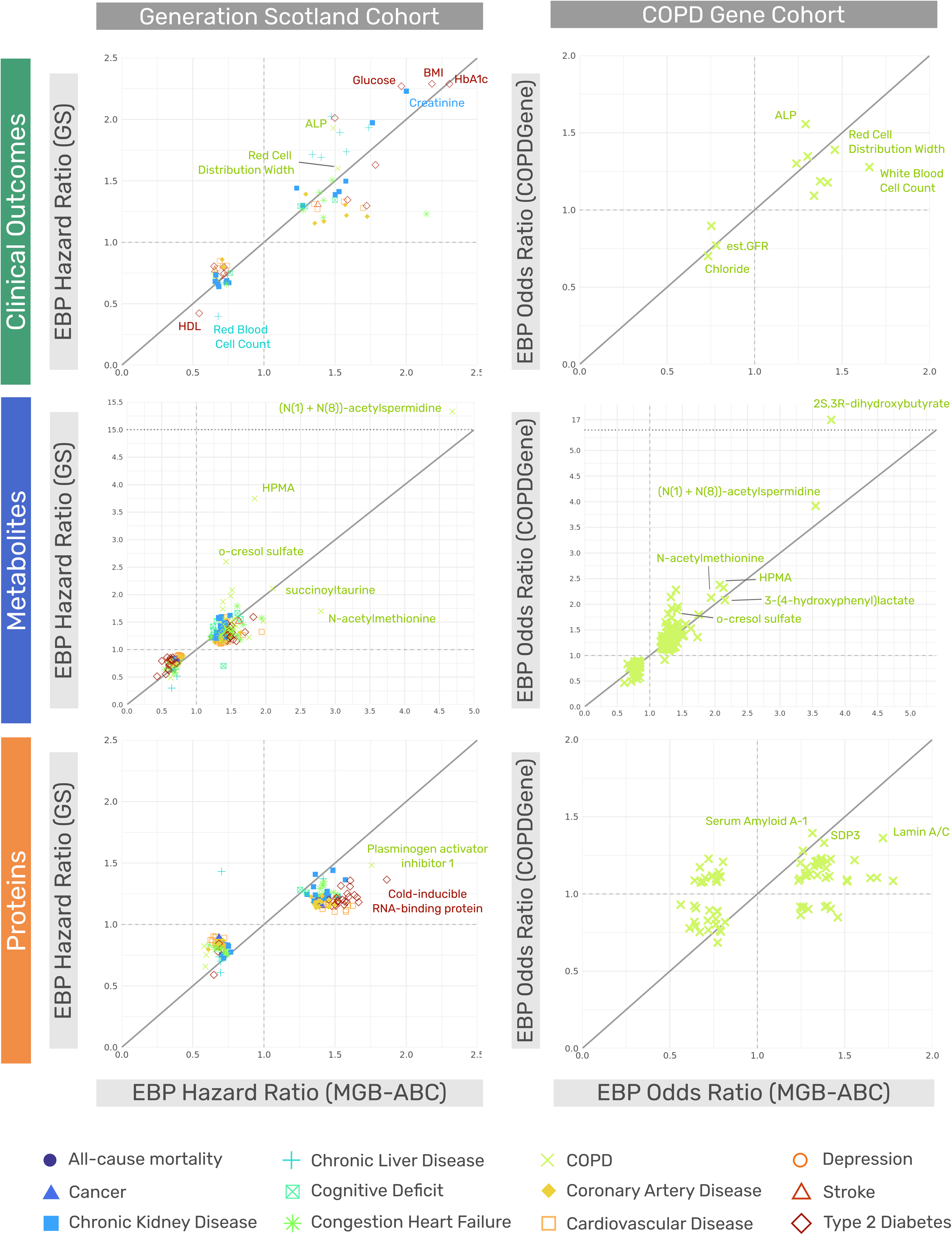

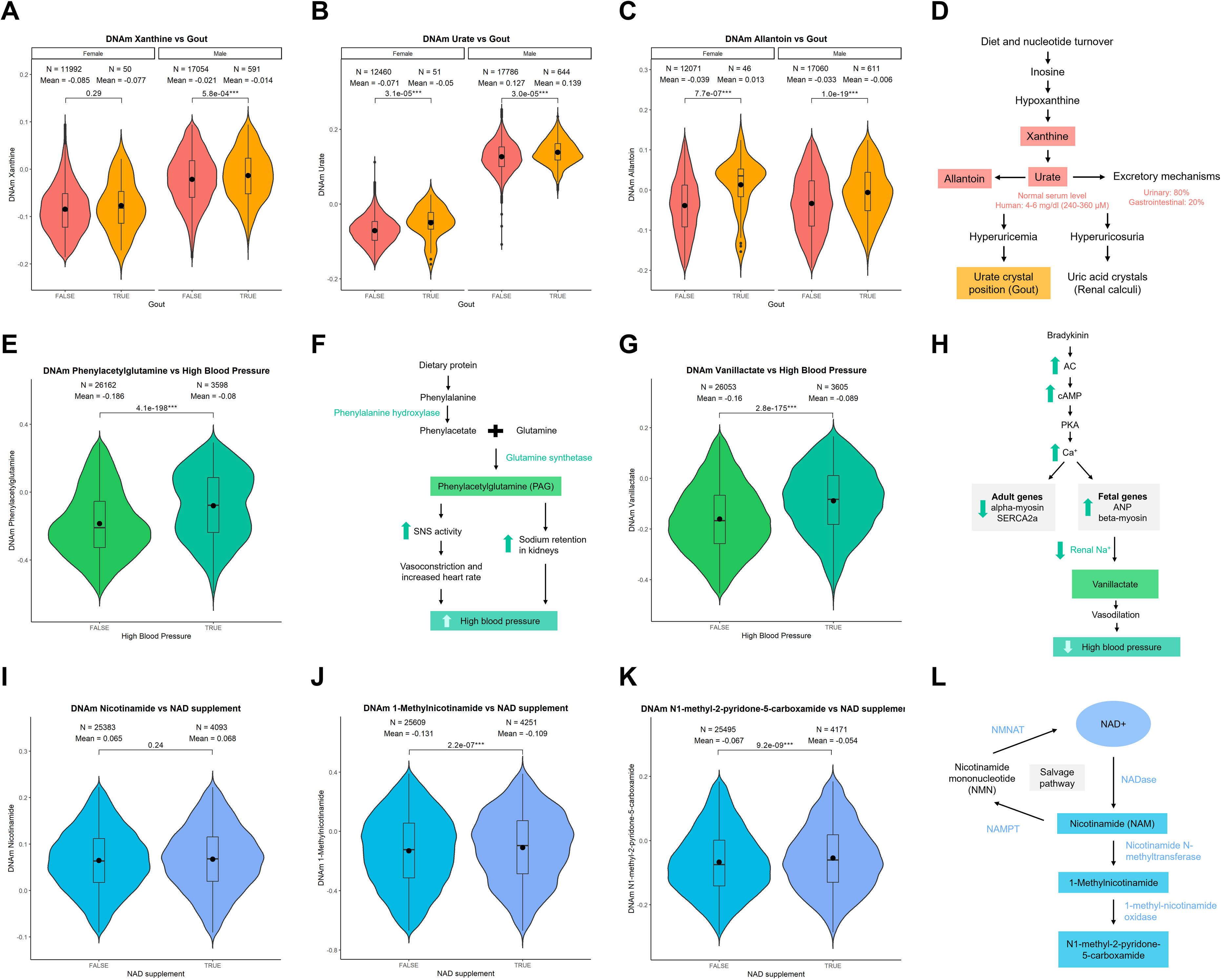

## Notes

### Summary of Updates

Since the initial medRxiv posting, the manuscript has been substantially revised. The title and abstract were updated, and the study was reframed from a two-cohort analysis to a multi-cohort development and validation framework. EBP models developed in 4,418 MGB-ABC participants are now evaluated across seven independent external cohorts (57,454 samples total), including MGB-ABC Narrow, MGB-LEOCC, CAMP, Costa Rica, Generation Scotland, COPDGene, and the TruDiagnostic Biobank. New external-validation analyses assess replication of EBP-biomarker correlations, disease associations, and classification of out-of-range clinical laboratory values across diverse ages, disease burdens, biospecimens, and DNAm platforms. Disease-association analyses and clinical-range performance metrics were recalculated, and statistical methods, multiple-testing procedures, longitudinal analyses, and limitations were clarified. The Discussion and Conclusions were revised to emphasize that EBPs are complementary population-level molecular proxies rather than replacements for direct clinical measurements, and that further validation is required before individual-level clinical use. The prior exploratory gene-ontology analysis was removed to focus the manuscript on external validation. Figures 1-6 and Table 1 were revised to reflect the expanded analyses; new external-validation results were added, and the longitudinal analysis was moved to the Supplement. Supplementary materials were reorganized and expanded to 33 supplementary figures and 13 supplementary tables. The author list, contribution statements, affiliations, disclosures, data/code availability, and references were also updated.

