## Supplemental Data 1 for "Leveraging epigenetic biomarker proxies for precision medicine"

**Additional file 1.** Supplementary Figures

**A**

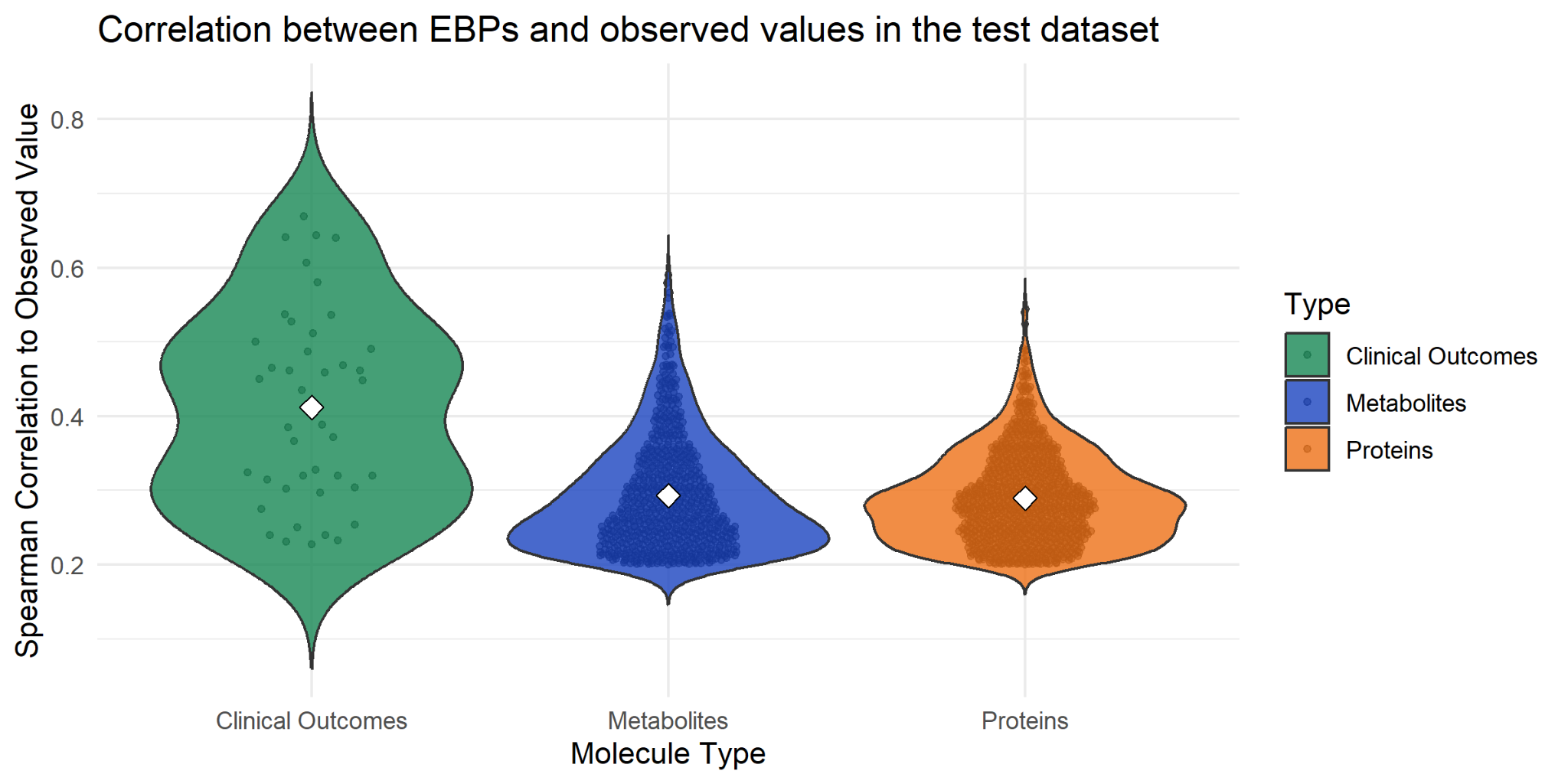

**B**

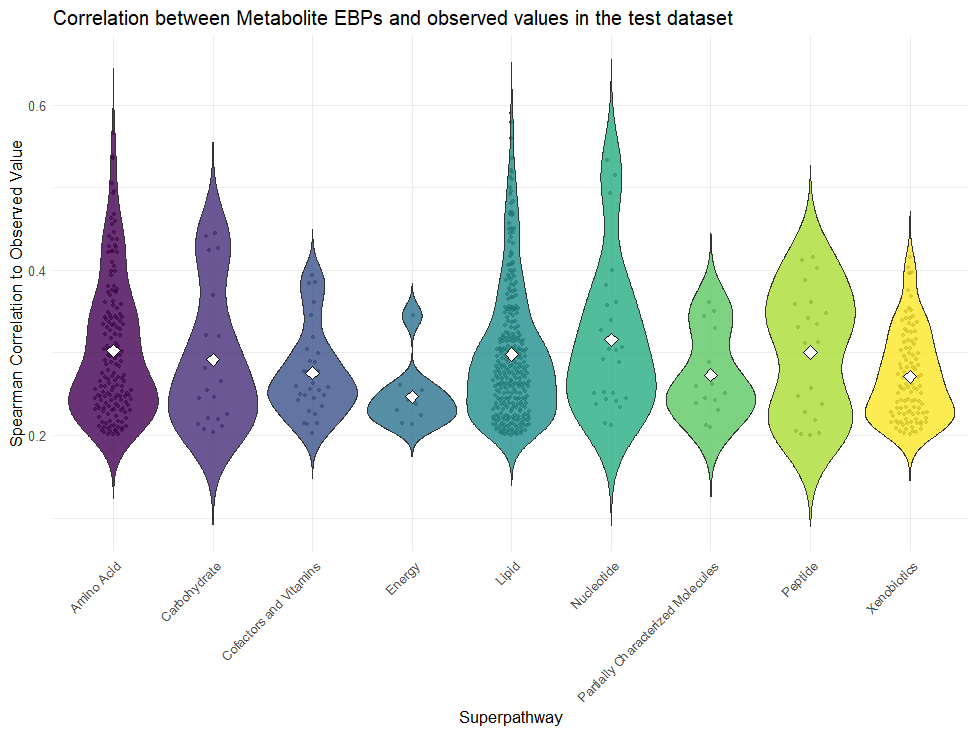

**Supplementary Figure 1.** Violin plots showing the correlation between EBPs and the observed counterparts in each type of molecule **(A)** and metabolites split based on an annotated superpathway **(B)**. To calculate these correlations, we used all samples from the testing dataset.

**A	B	C**

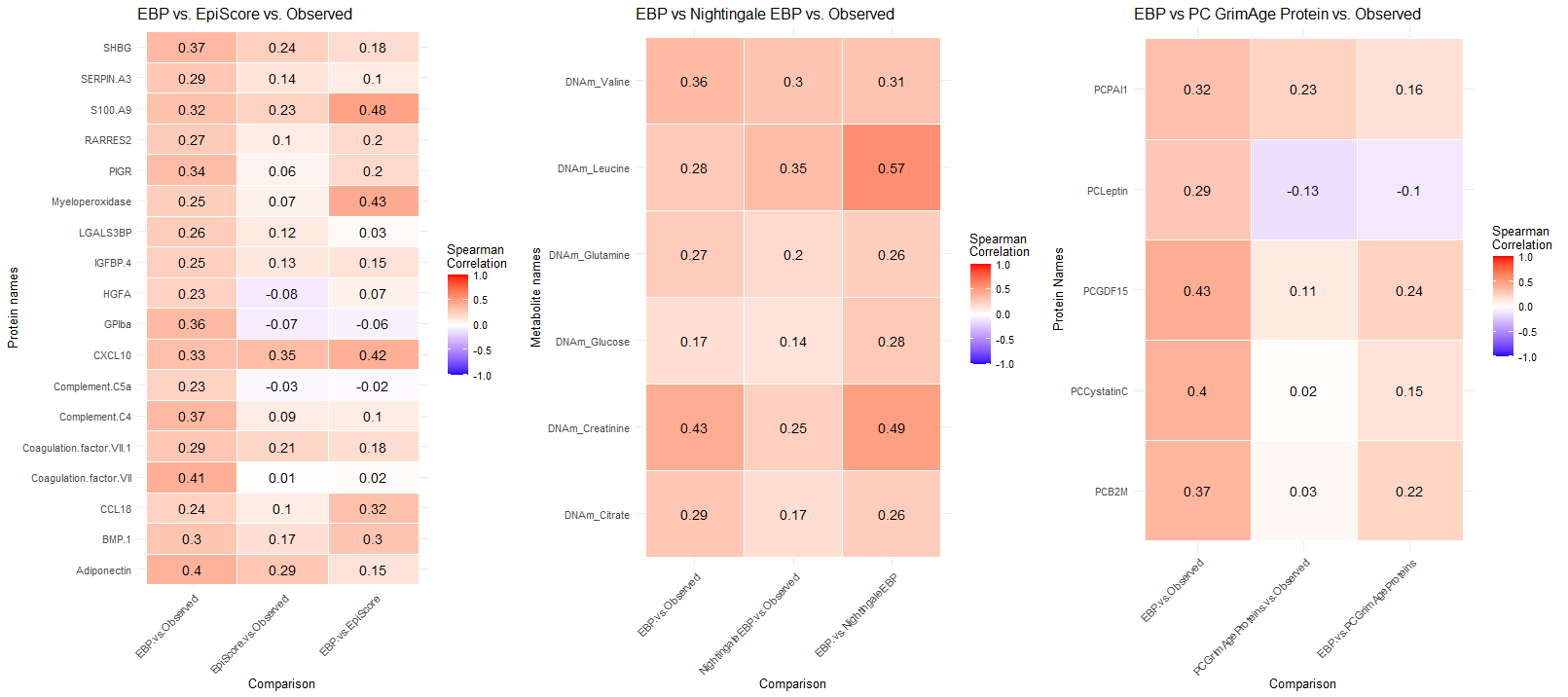

**Supplementary Figure 2.** Correlation plots illustrating the relationships between (A) developed protein EBPs, Marioni EpiScores, and their observed counterparts measured with the Seer platform; (B) metabolite EBPs, matched Nightingale epigenetic proxies, and their observed counterparts measured with Metabolon metabolomics; and (C) protein EBPs, principal component (PC)-based surrogate proteins from GrimAge, and their observed counterparts measured with the Seer platform.

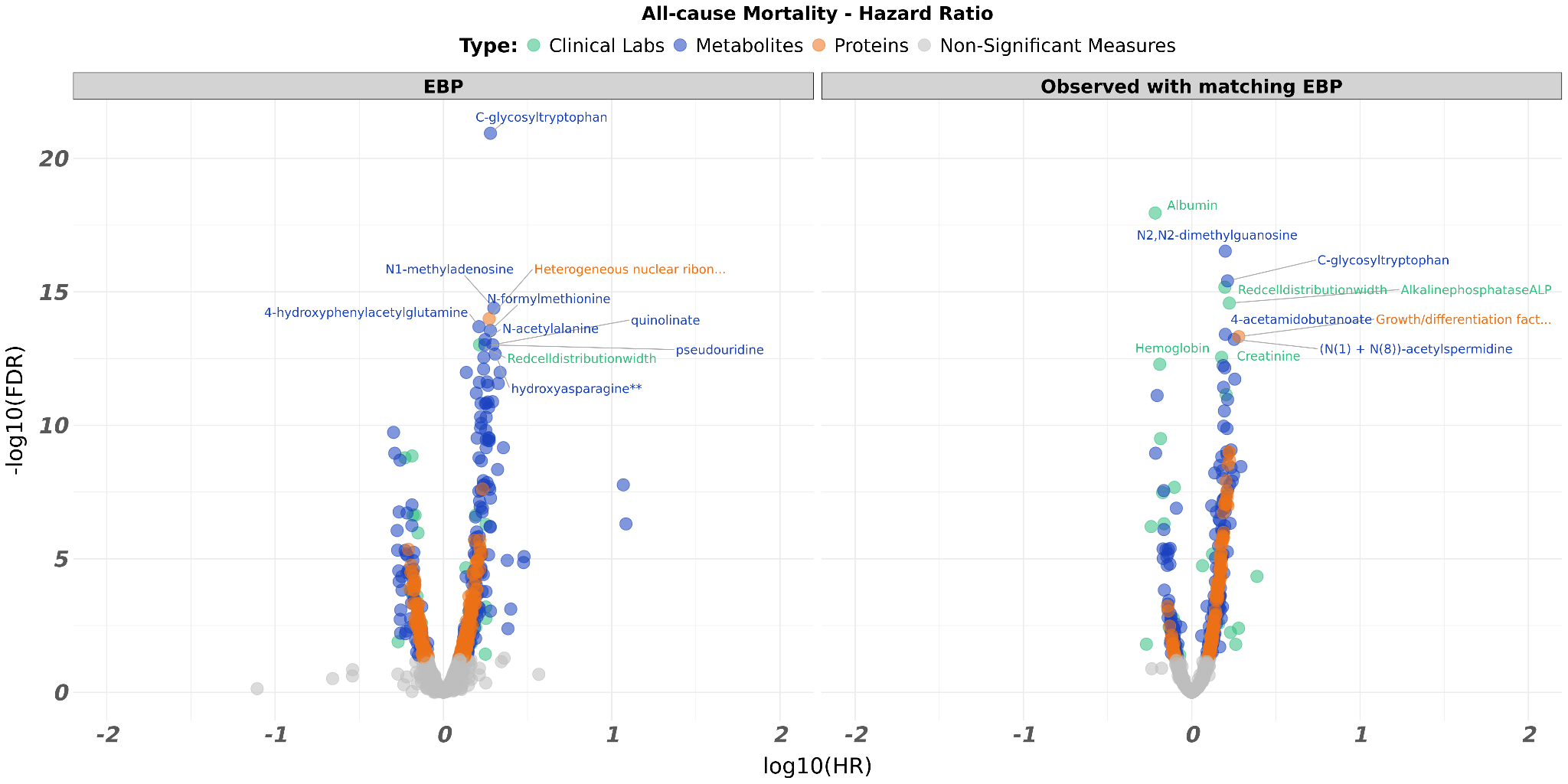

**Supplementary Figure 3.** Volcano plot comparing the EBP values to the matched observed values for all-cause mortality.

**A**

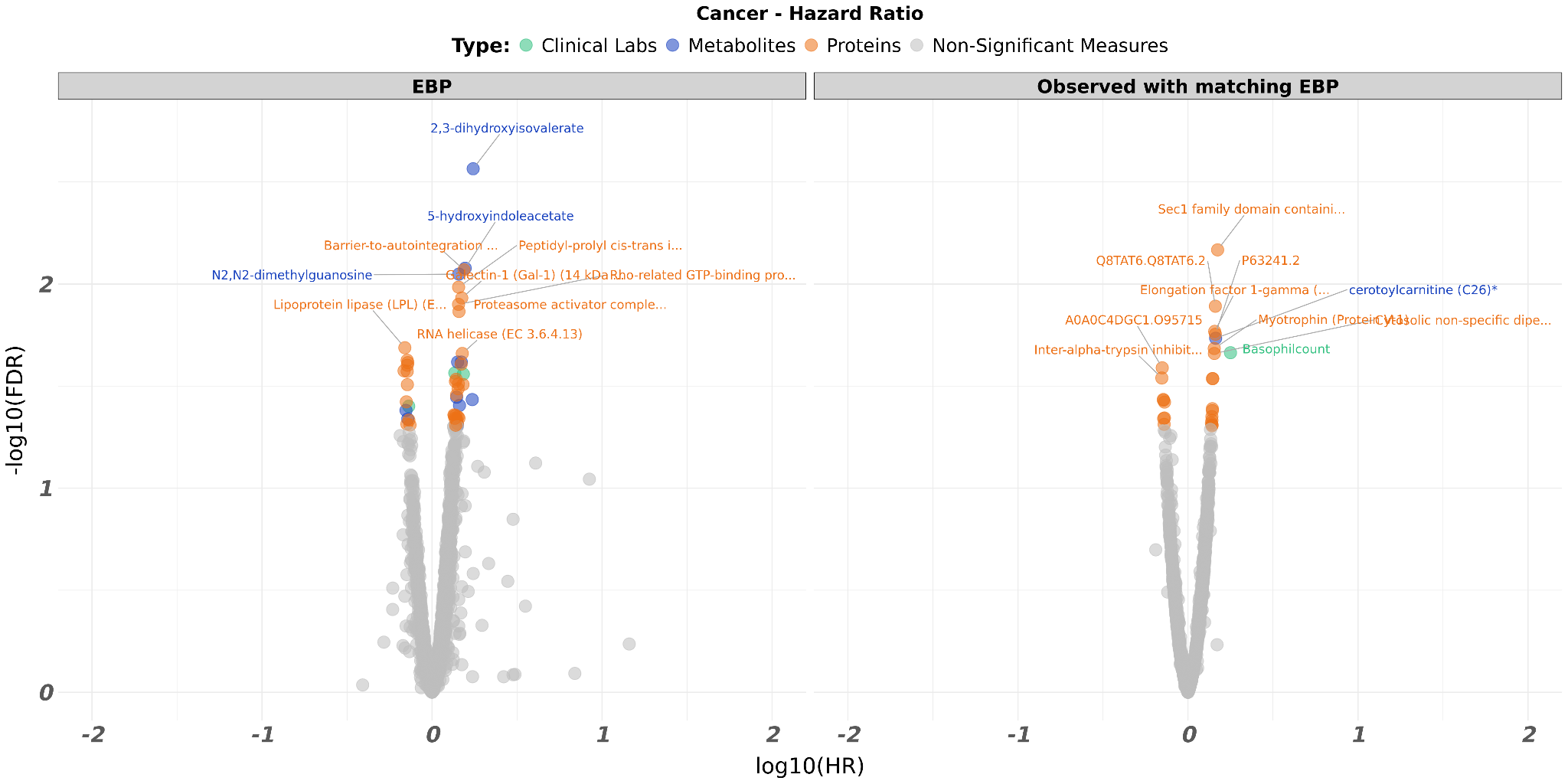

**B**

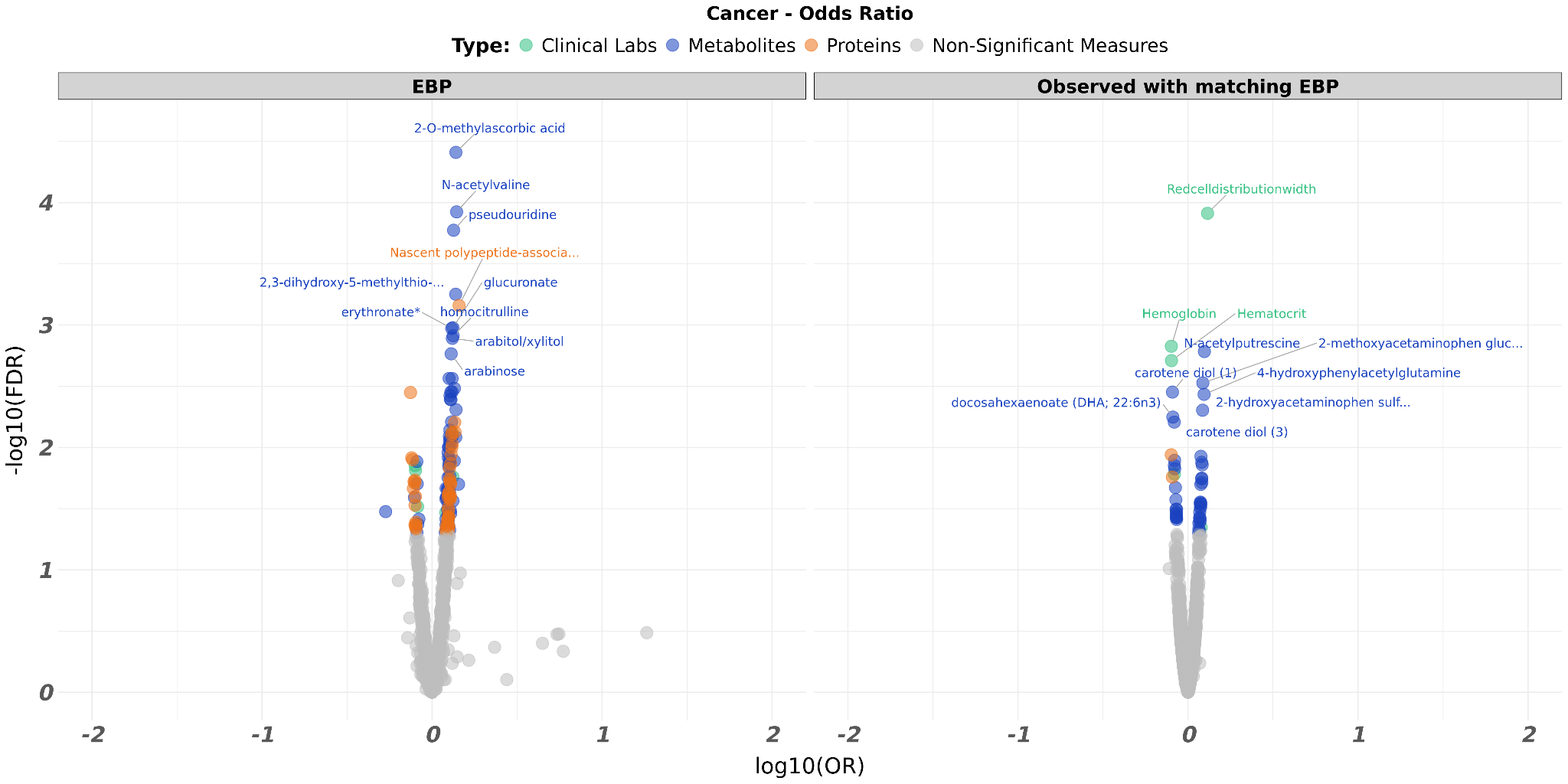

**Supplementary Figure 4.** Volcano plot comparing the EBP values to the matched observed values for cancer. (**A**) The plot is according to Hazard Ratios. (**B**) The plot is according to Odd Ratios.

**A**

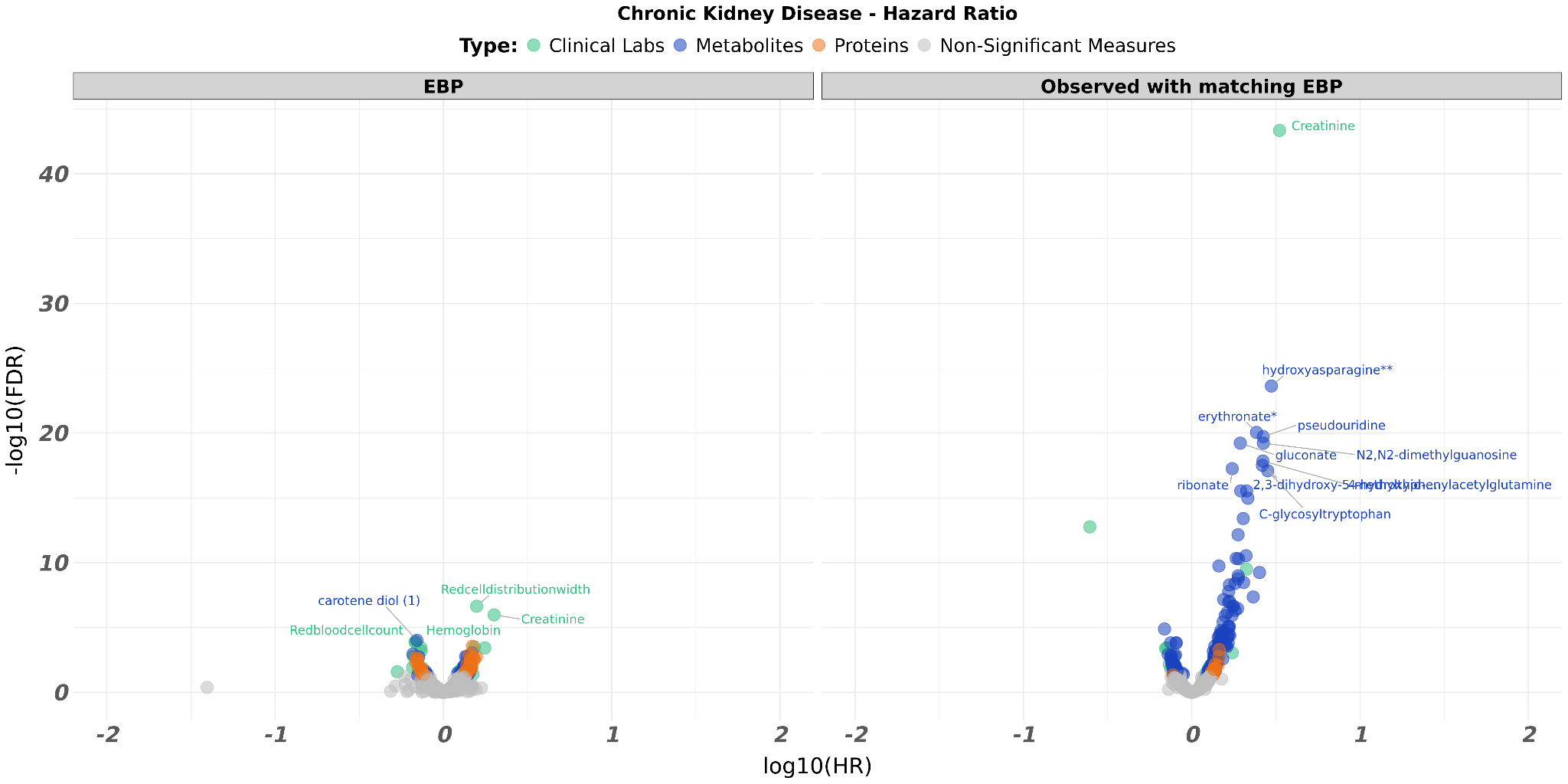

**B**

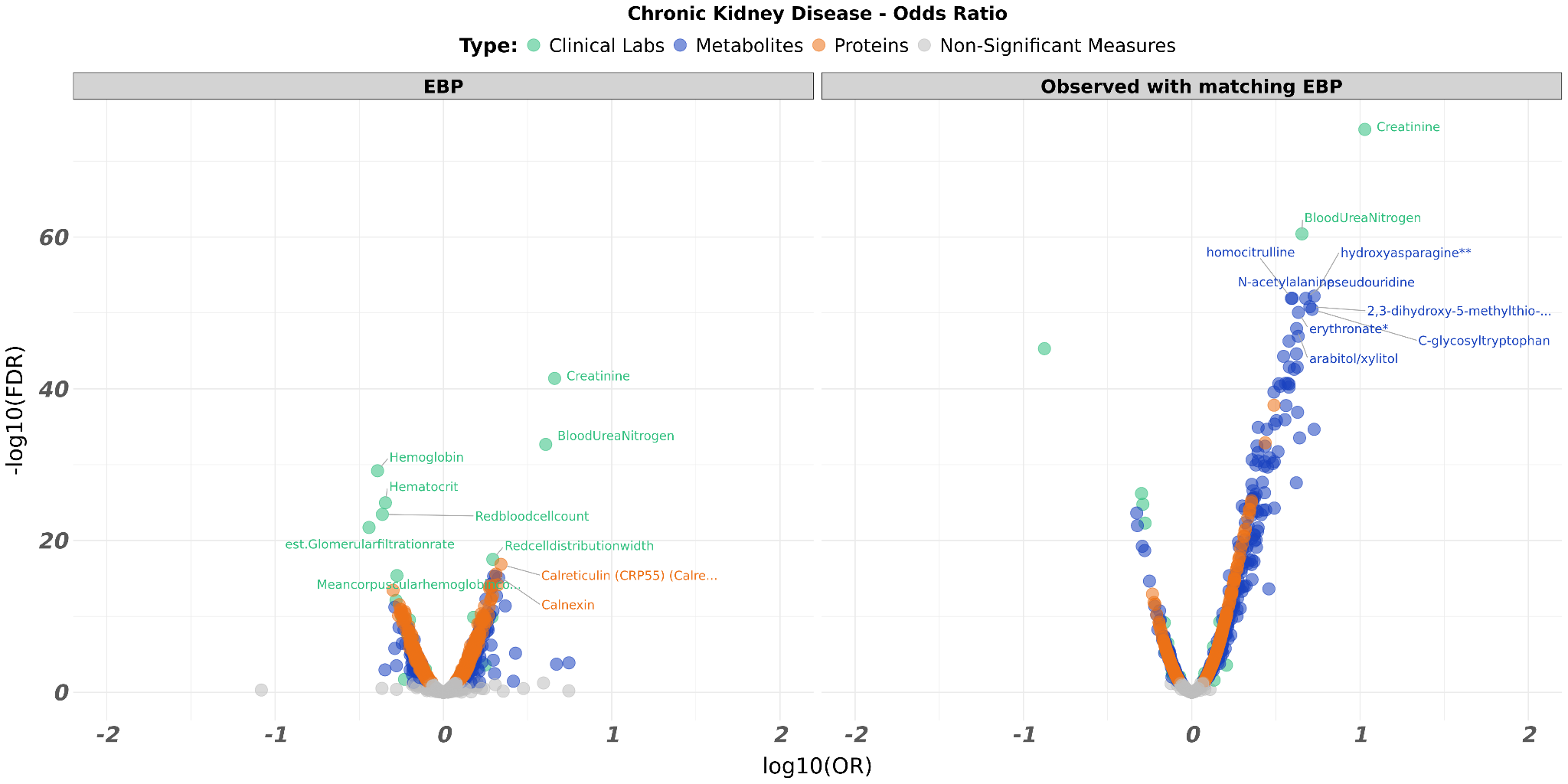

**Supplementary Figure 5.** Volcano plot comparing the EBP values to the matched observed values for chronic kidney disease. (**A**) The plot is according to Hazard Ratios. (**B**) The plot is according to Odd Ratios.

**A**

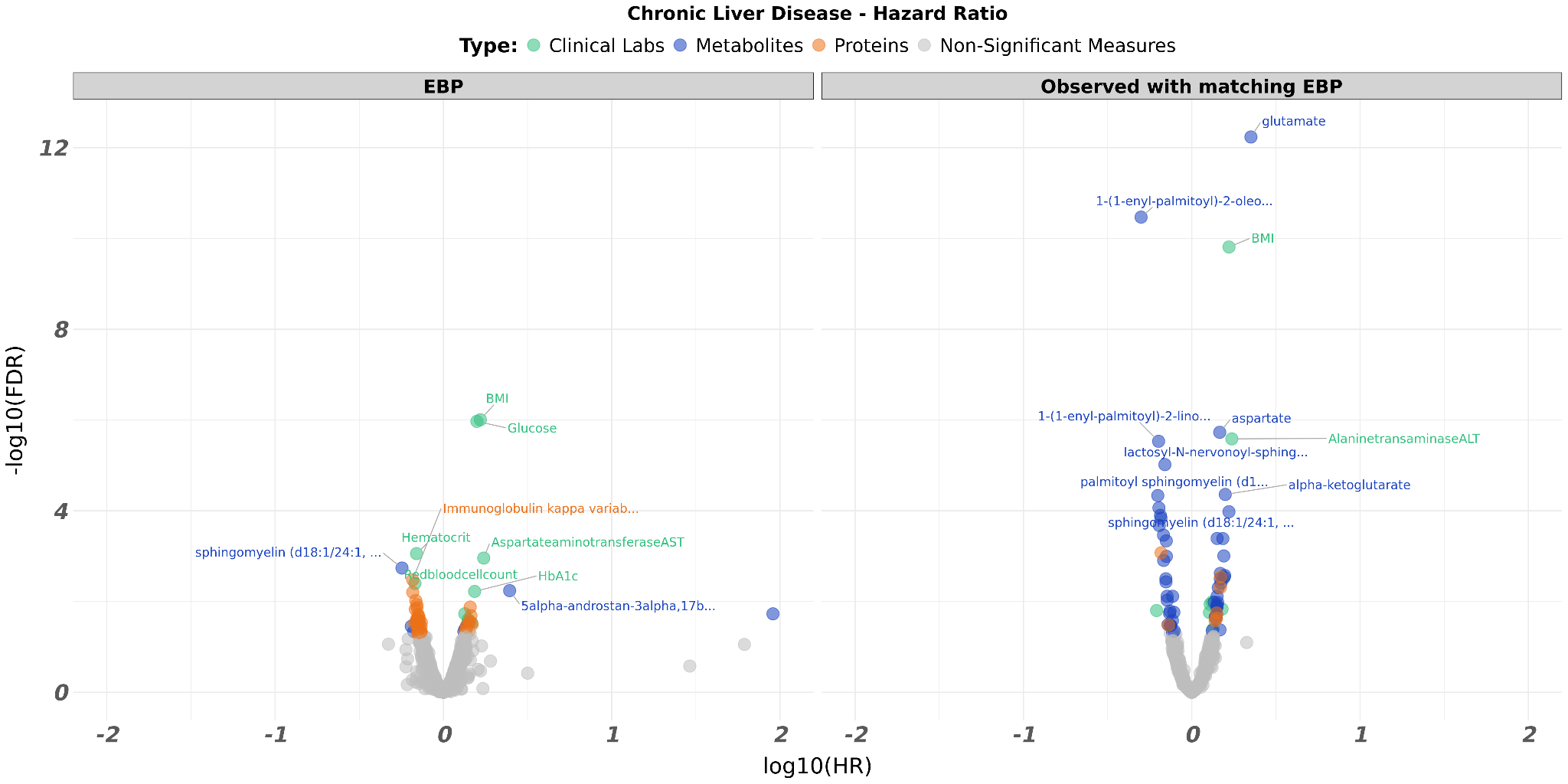

**B**

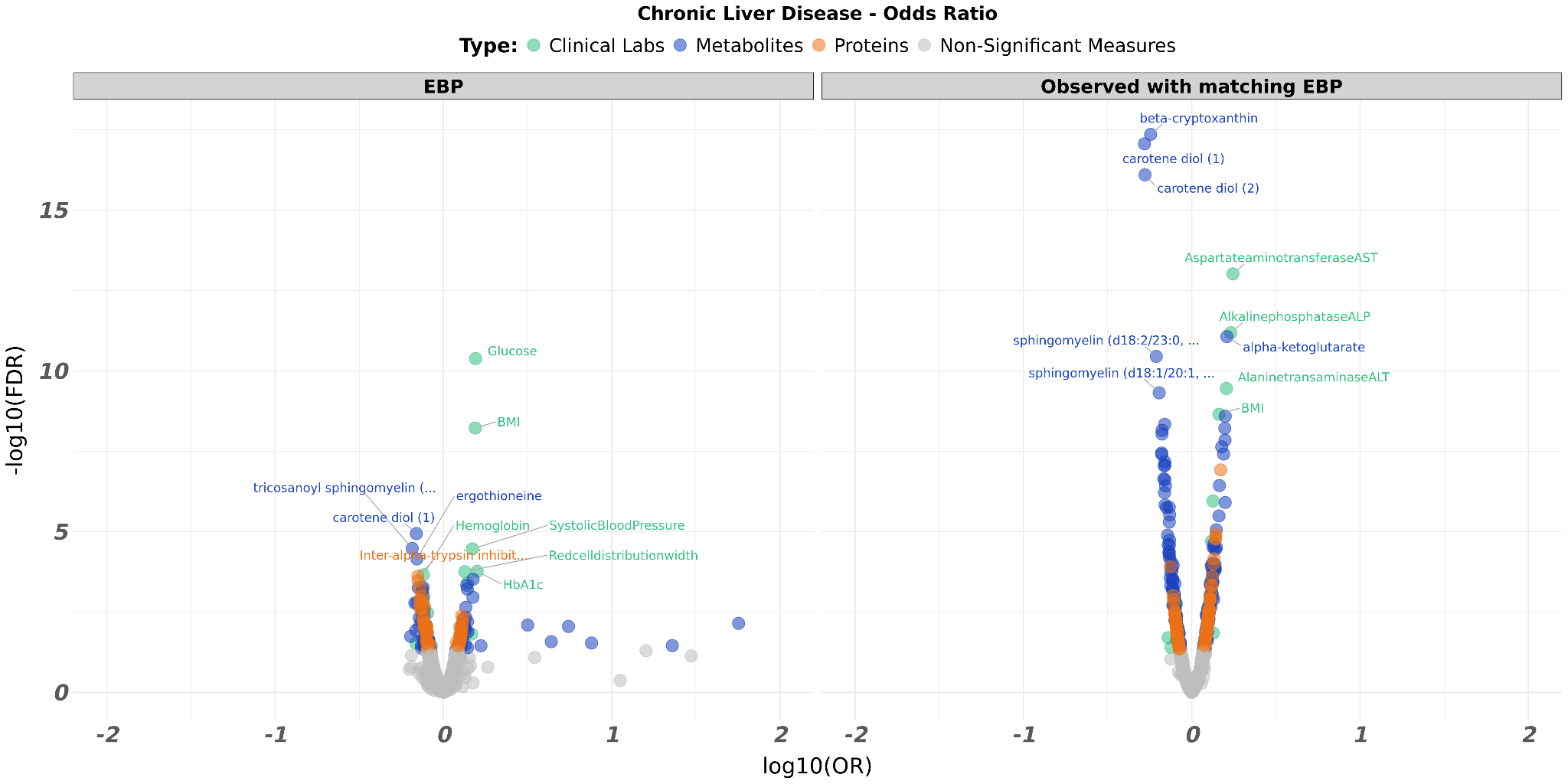

**Supplementary Figure 6.** Volcano plot comparing the EBP values to the matched observed values for chronic liver disease. (**A**) The plot is according to Hazard Ratios. (**B**) The plot is according to Odd Ratios.

**A**

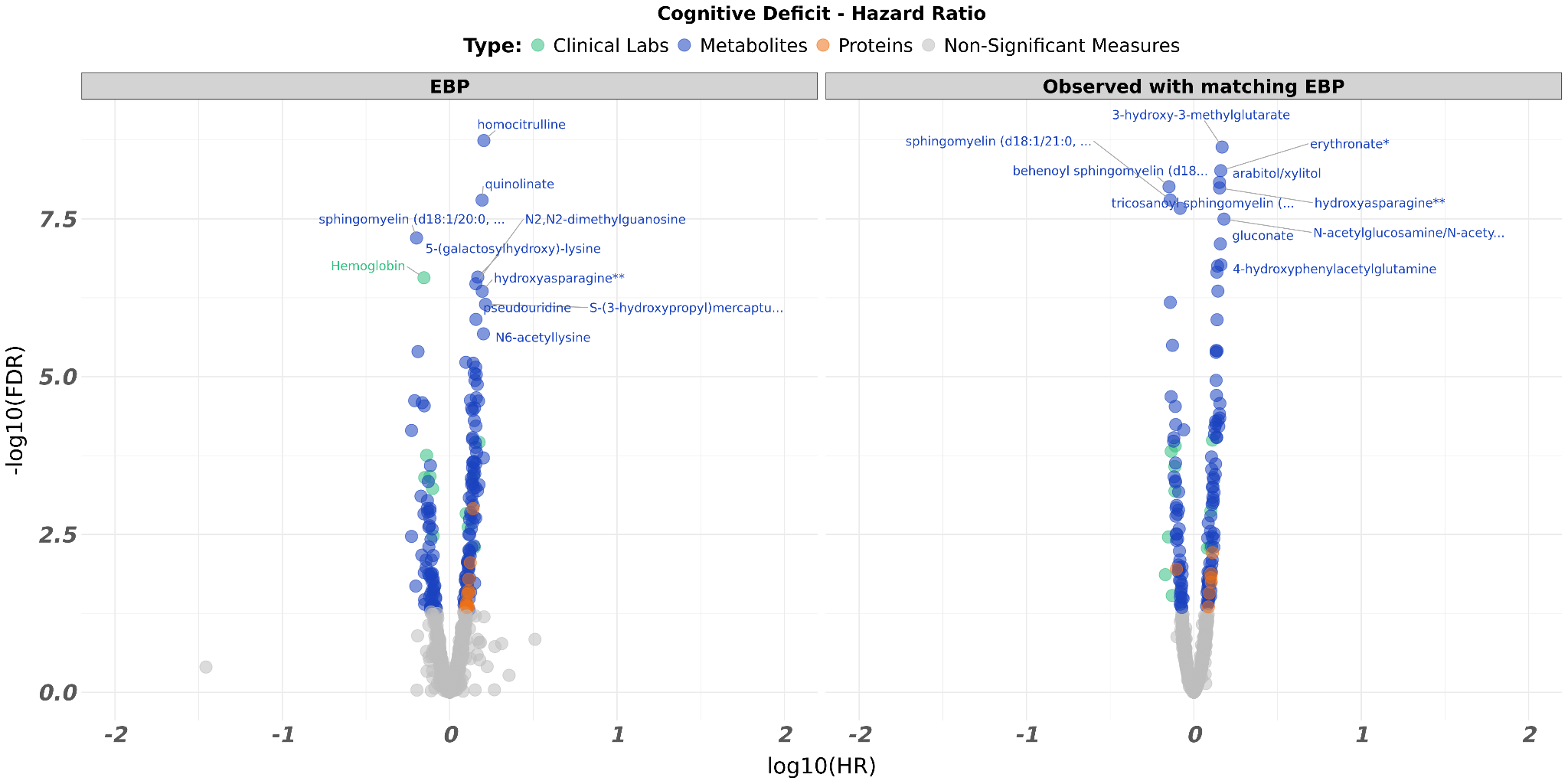

**B**

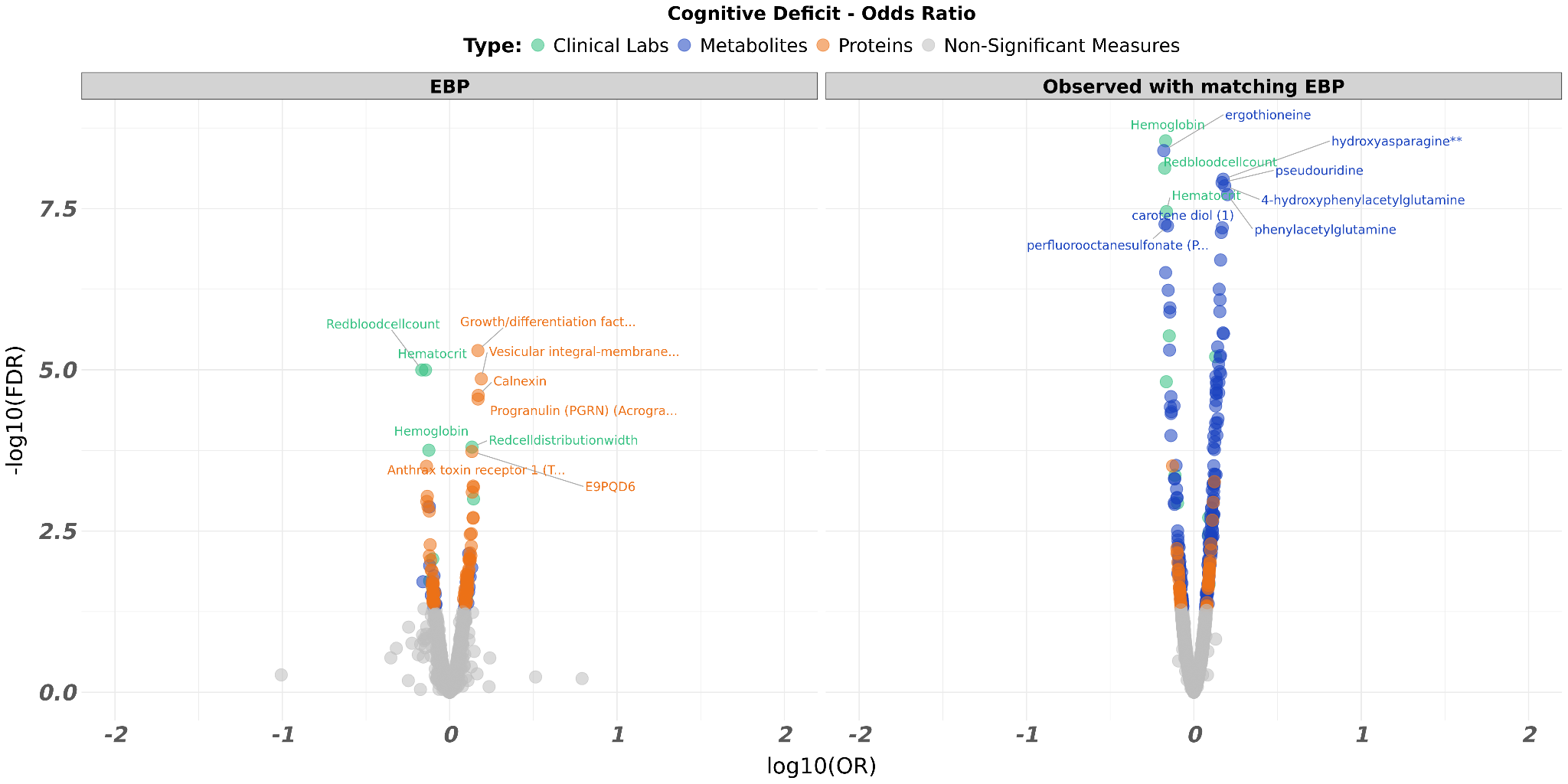

**Supplementary Figure 7.** Volcano plot comparing the EBP values to the matched observed values for cognitive deficit. (**A**) The plot is according to Hazard Ratios. (**B**) The plot is according to Odd Ratios.

**A**

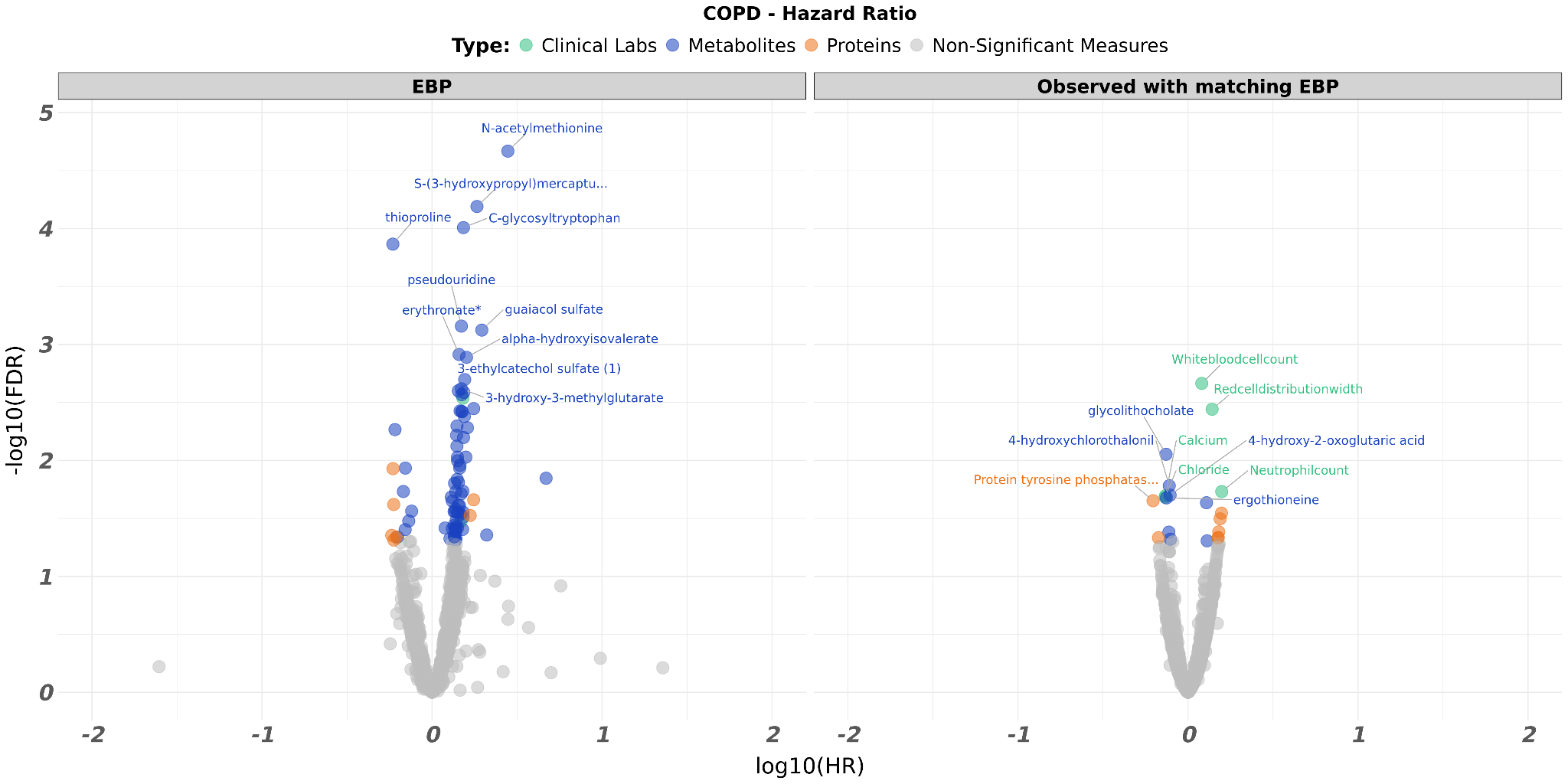

**B**

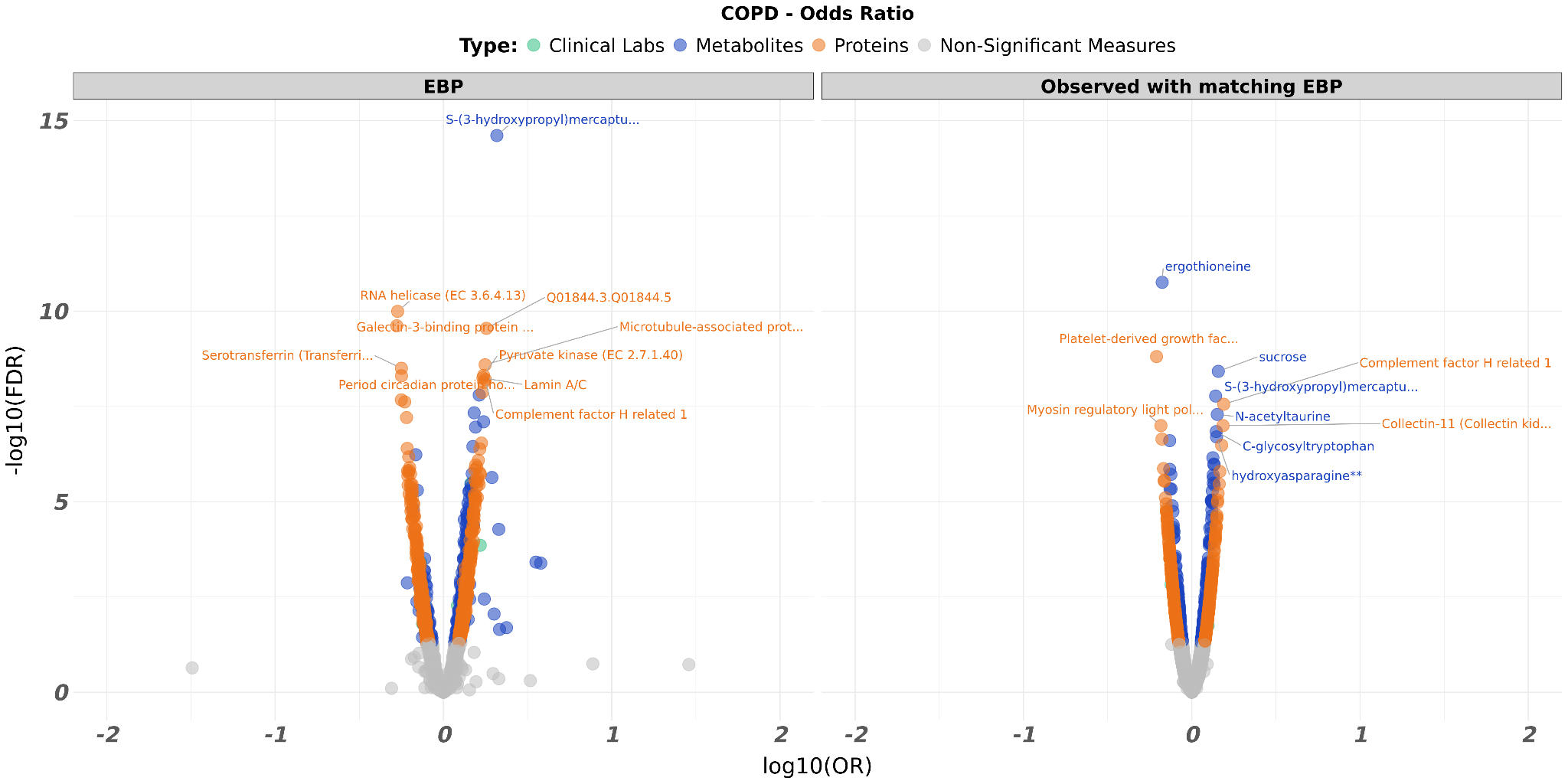

**Supplementary Figure 8.** Volcano plot comparing the EBP values to the matched observed values for COPD. (**A**) The plot is according to Hazard Ratios. (**B**) The plot is according to Odd Ratios.

**A**

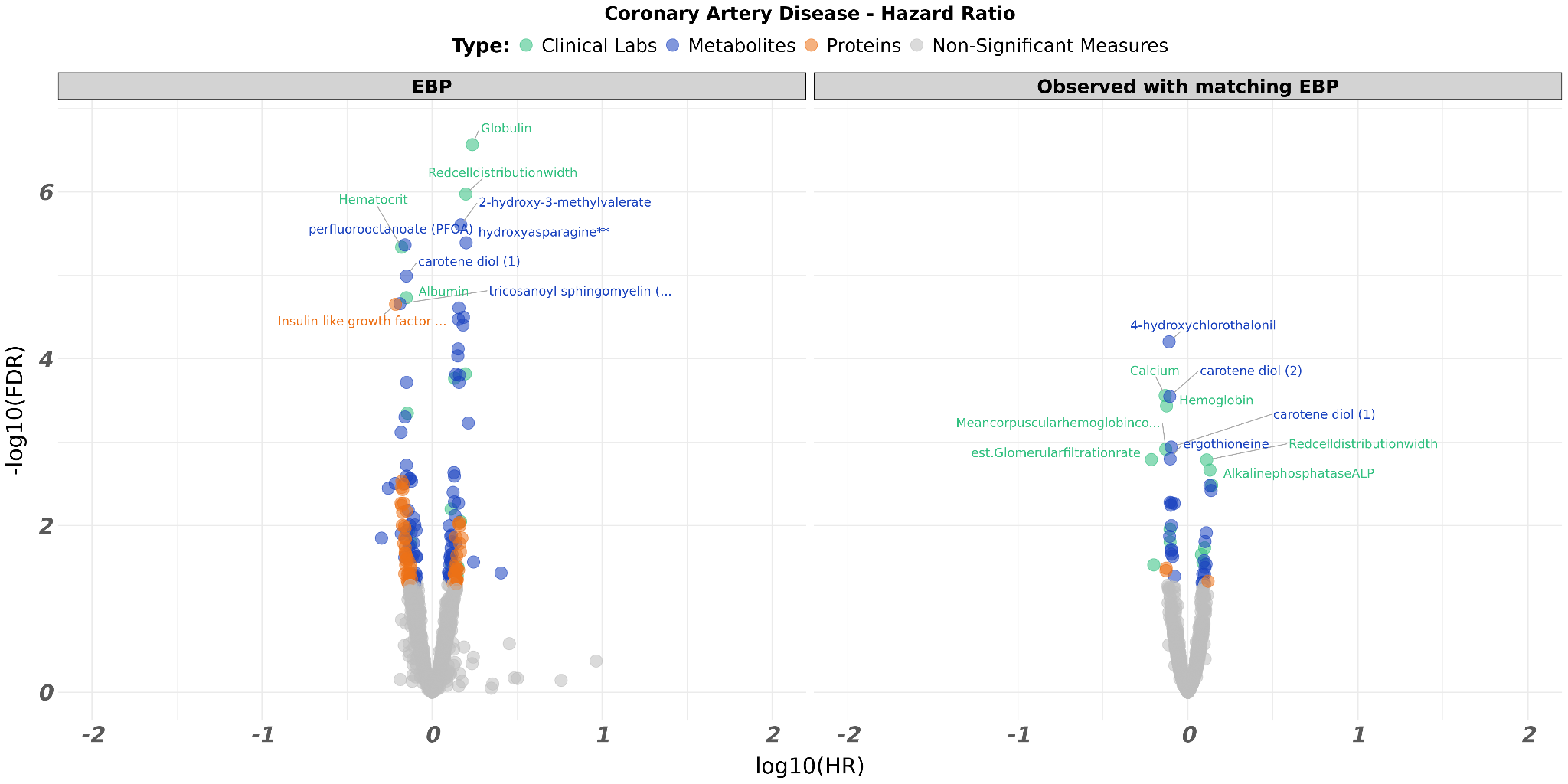

**B**

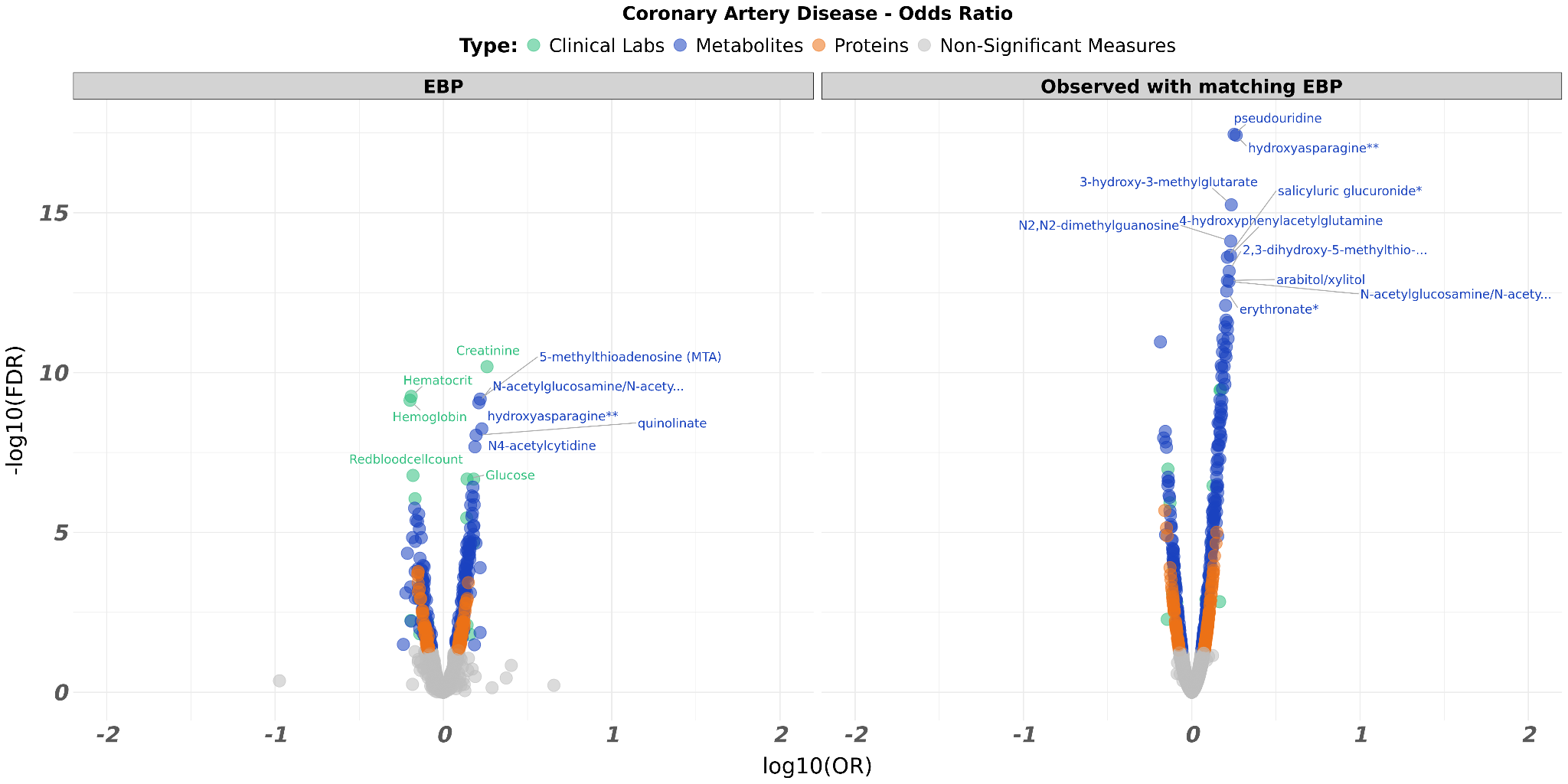

**Supplementary Figure 9.** Volcano plot comparing the EBP values to the matched observed values for coronary artery disease. (**A**) The plot is according to Hazard Ratios. (**B**) The plot is according to Odd Ratios.

**A**

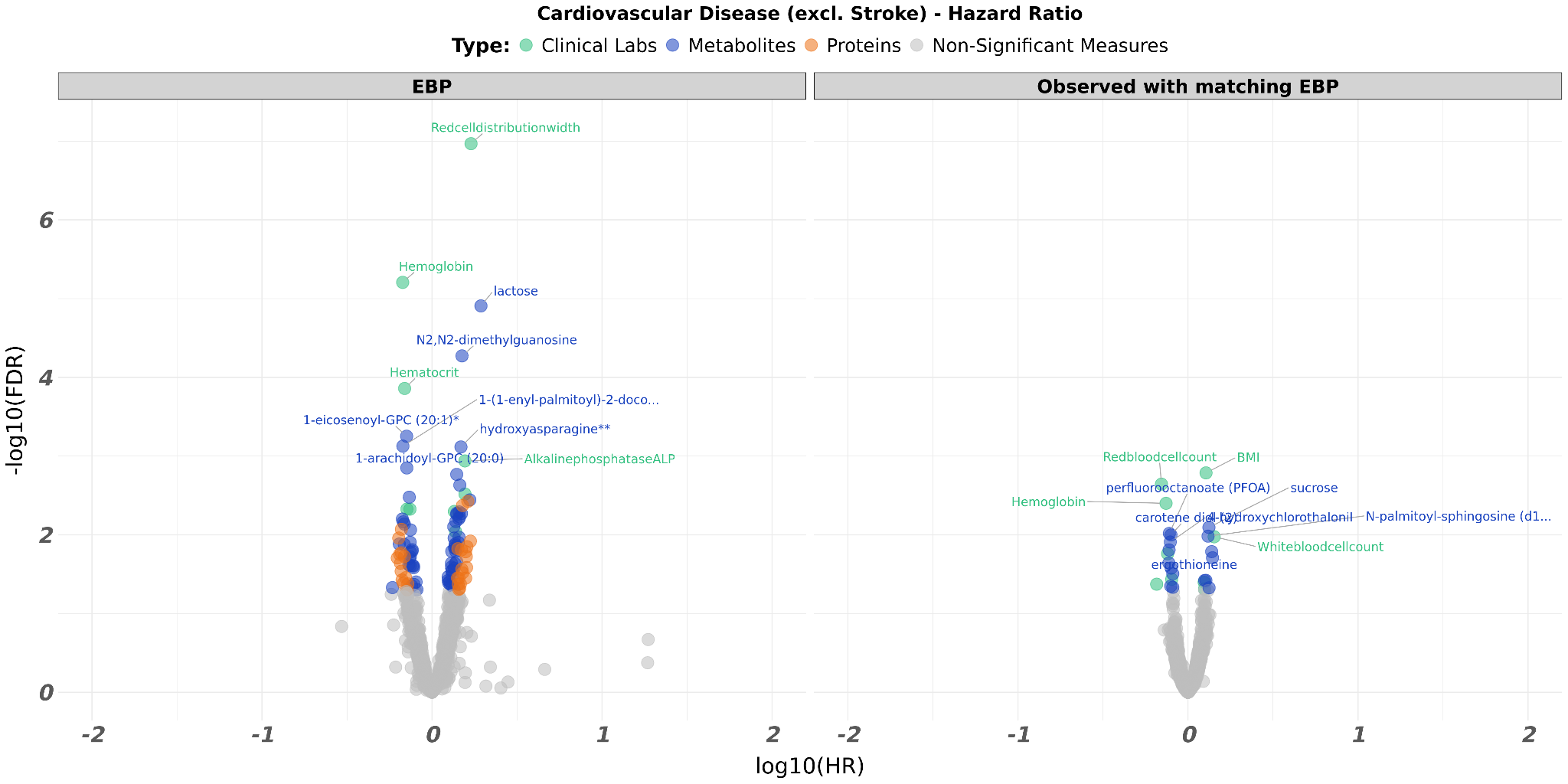

**B**

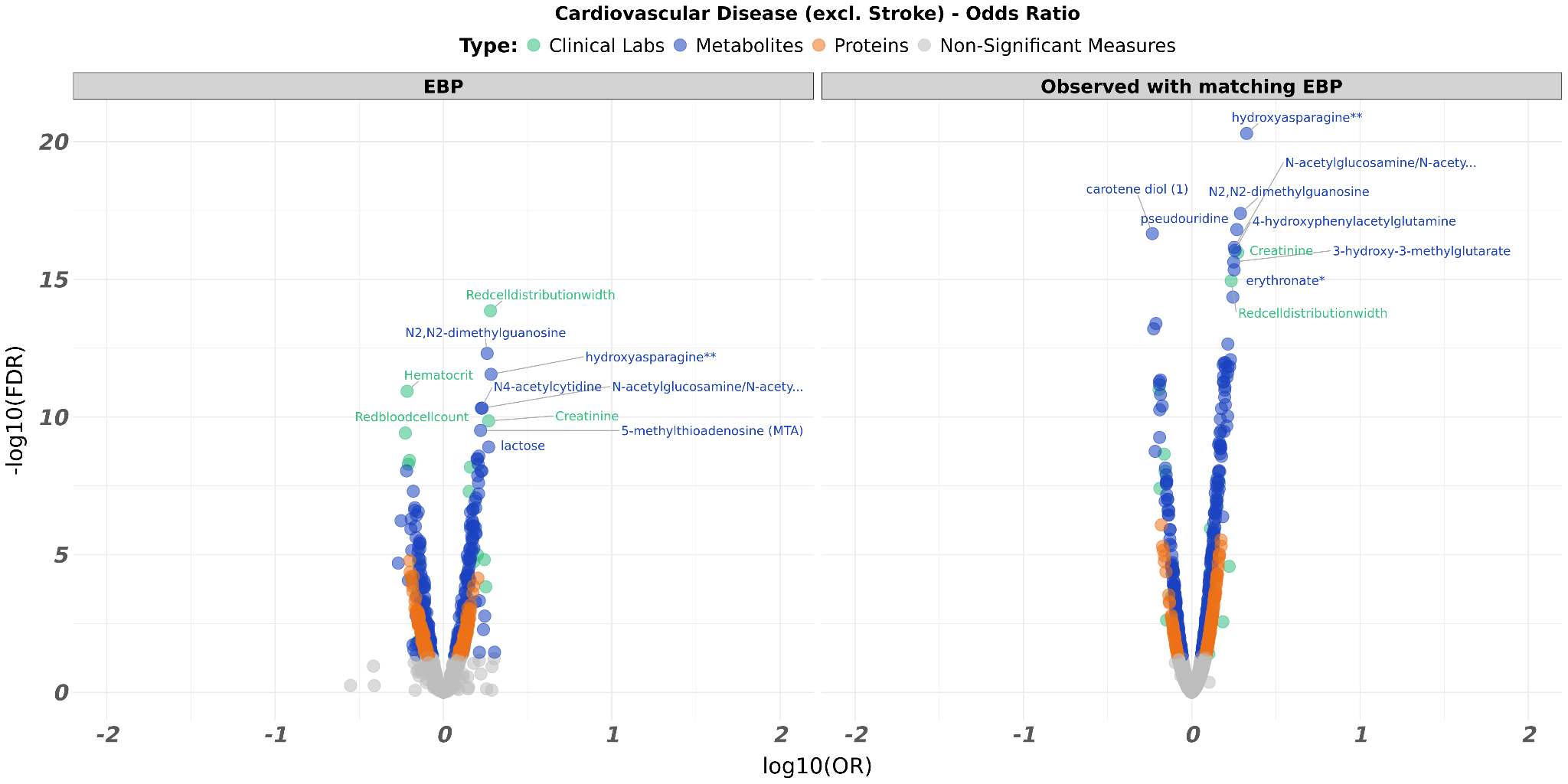

**Supplementary Figure 10.** Volcano plot comparing the EBP values to the matched observed values for cardiovascular disease excluding stroke. (**A**) The plot is according to Hazard Ratios. (**B**) The plot is according to Odd Ratios.

**A**

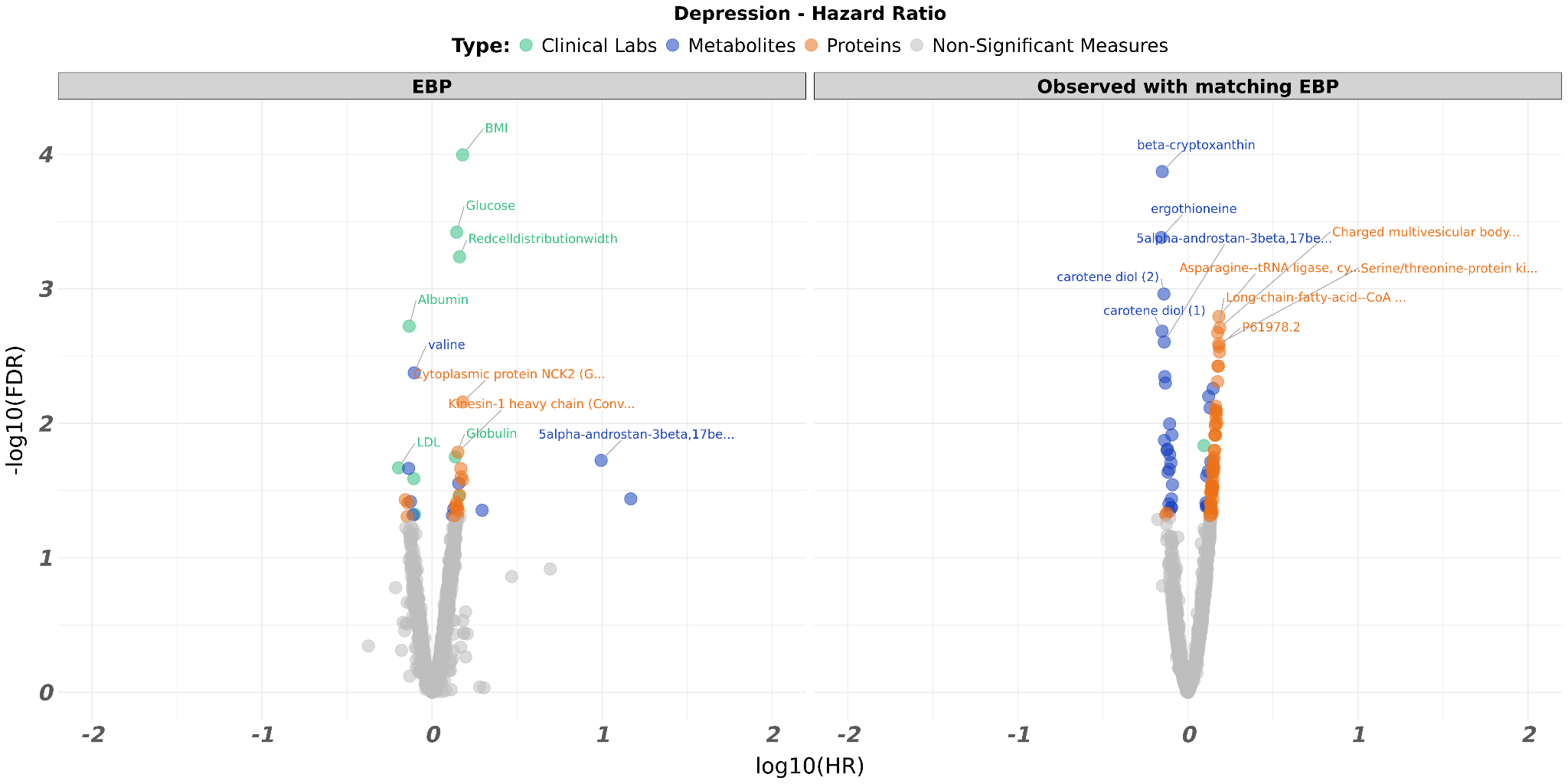

**B**

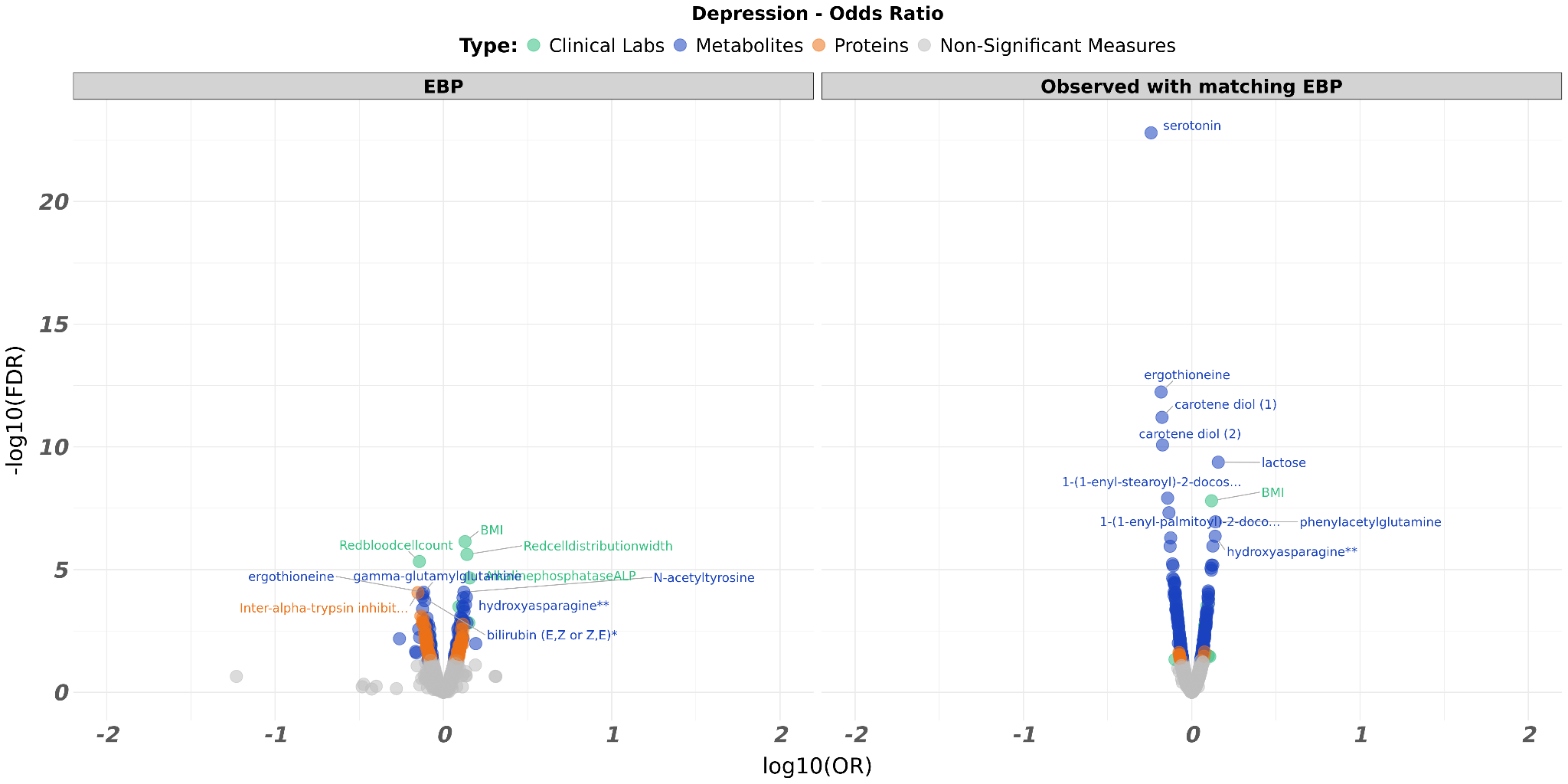

**Supplementary Figure 11.** Volcano plot comparing the EBP values to the matched observed values for depression. (**A**) The plot is according to Hazard Ratios. (**B**) The plot is according to Odd Ratios.

**A**

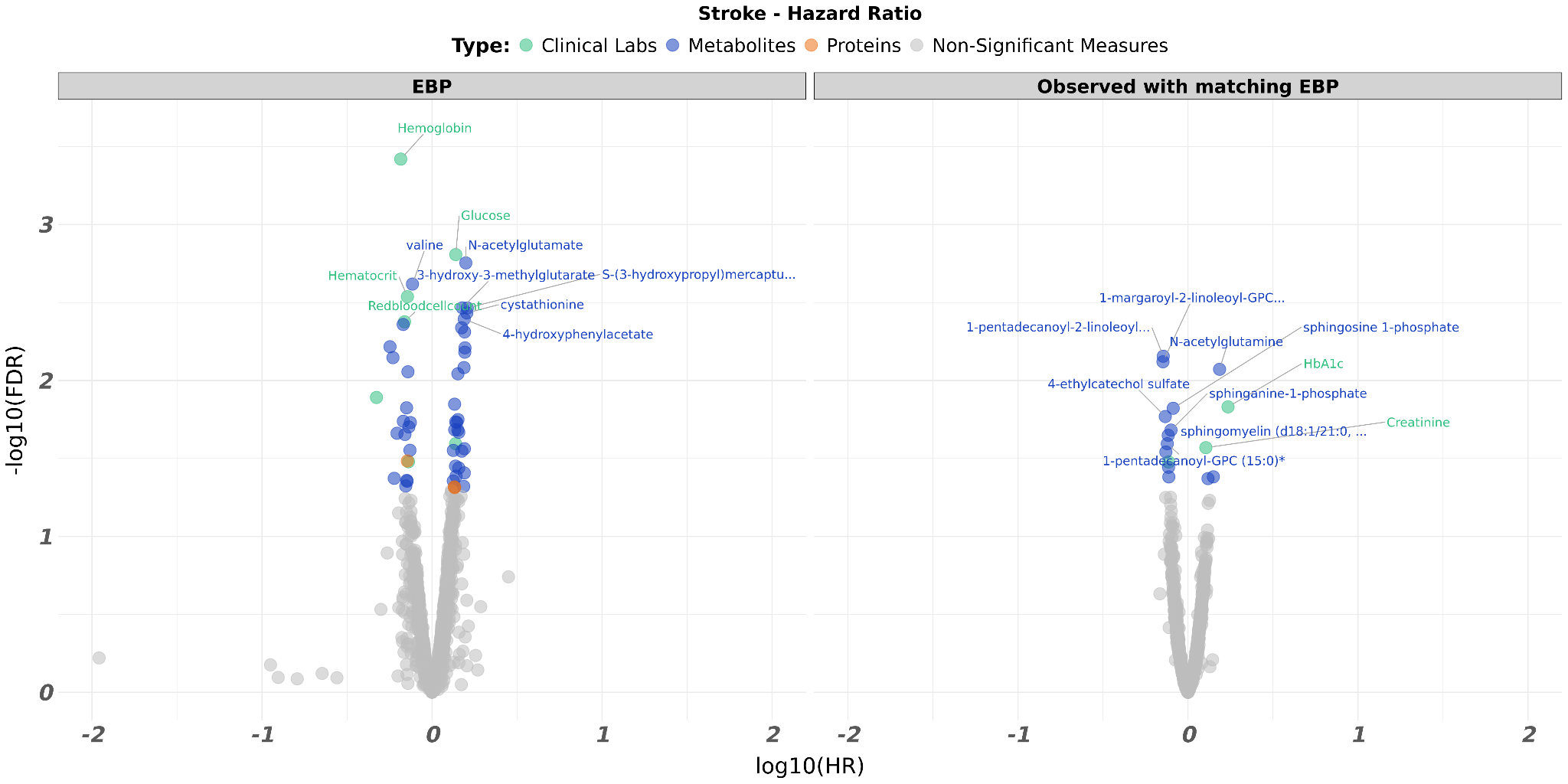

**B**

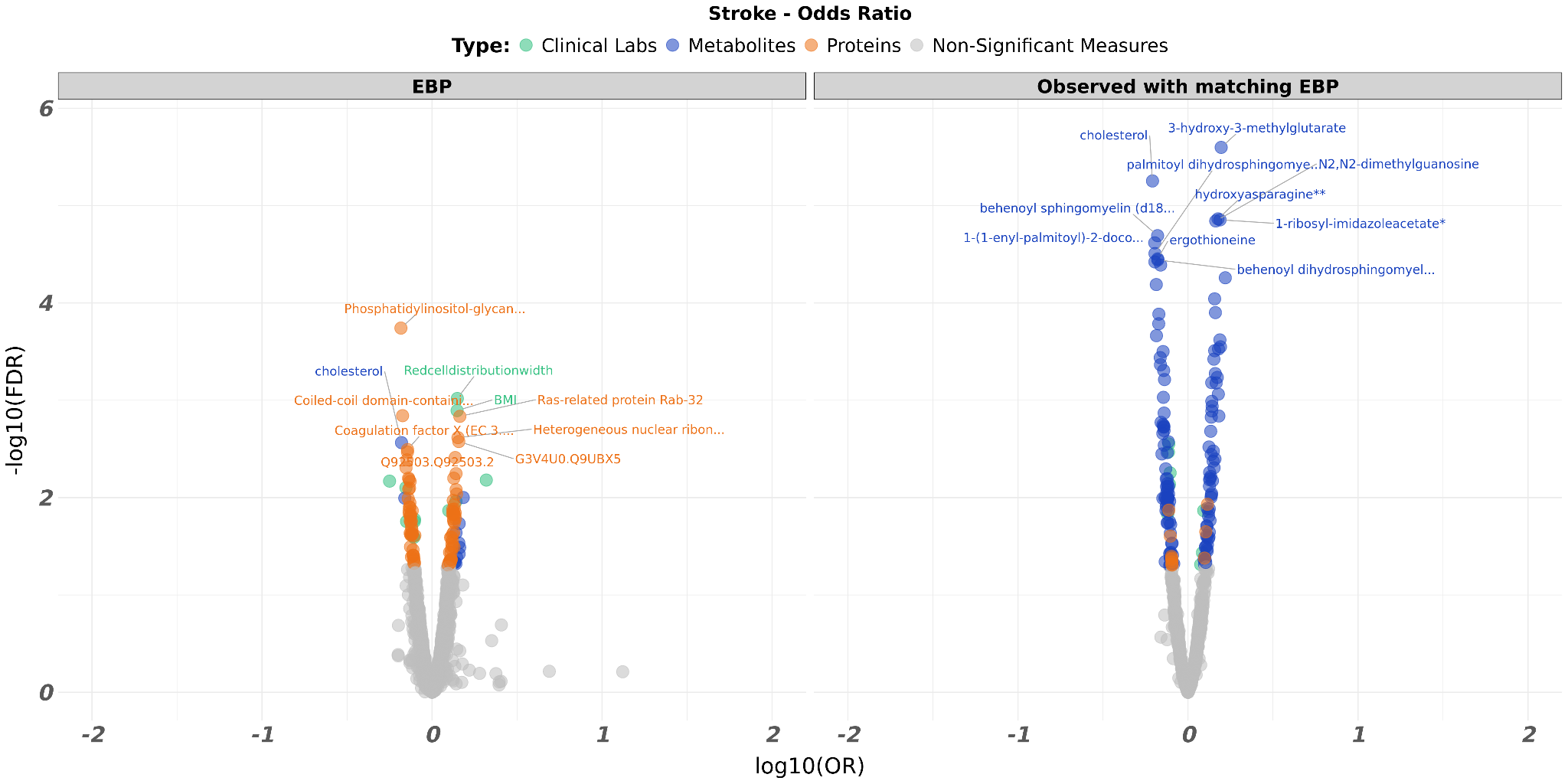

**Supplementary Figure 12.** Volcano plot comparing the EBP values to the matched observed values for stroke. (**A**) The plot is according to Hazard Ratios. (**B**) The plot is according to Odd Ratios.

**A**

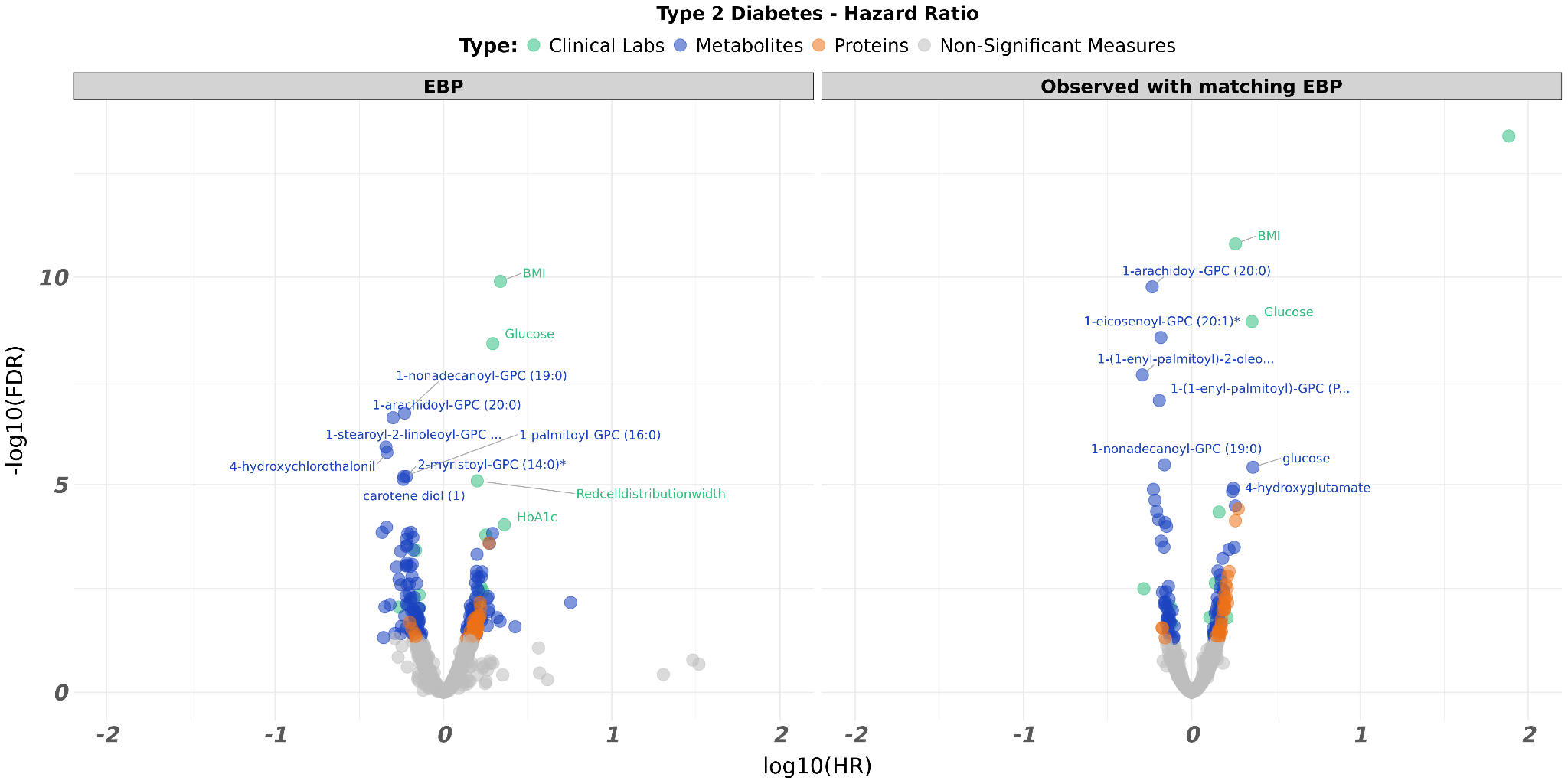

**B**

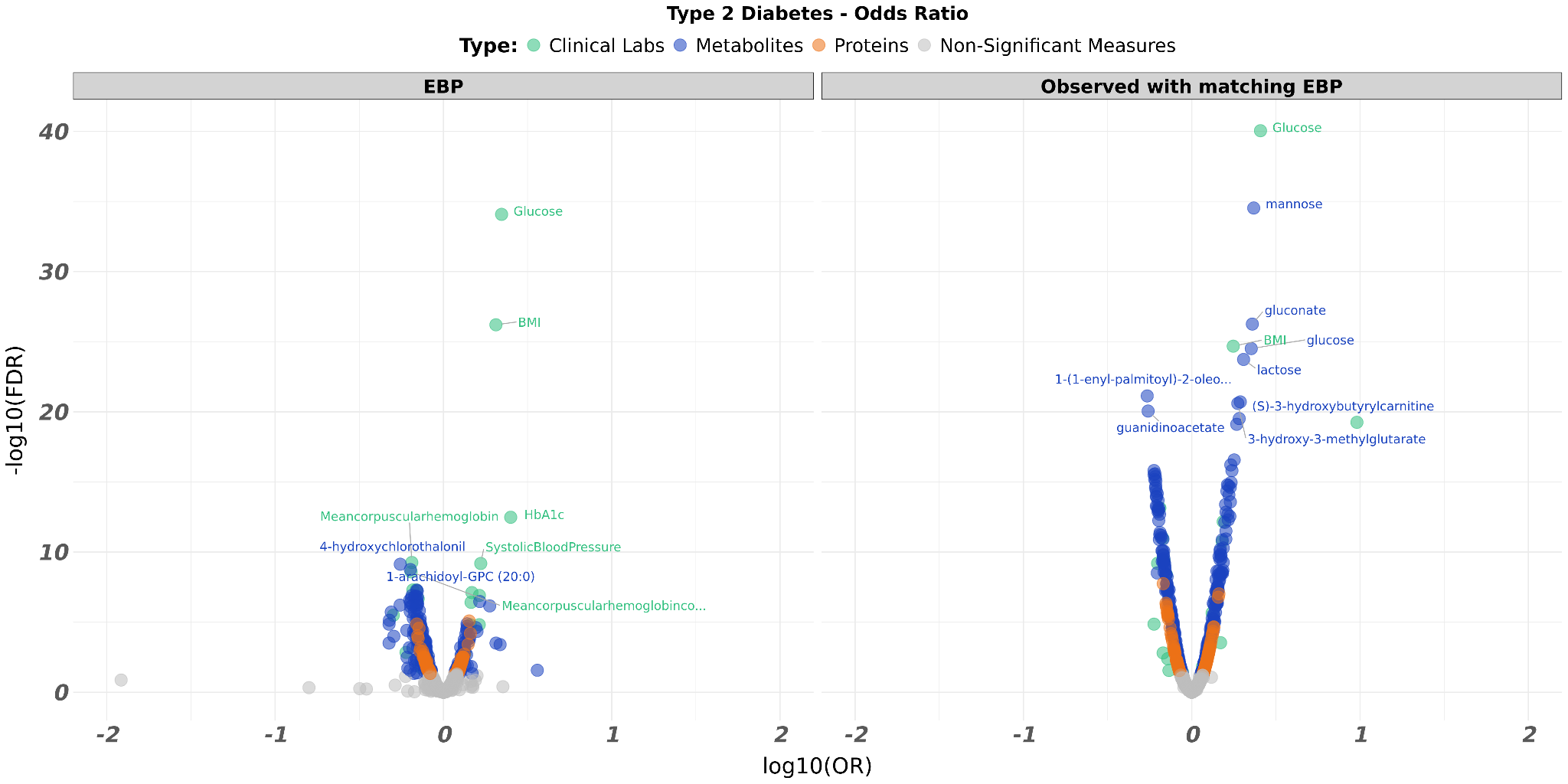

**Supplementary Figure 13.** Volcano plot comparing the EBP values to the matched observed values for Type 2 diabetes. (**A**) The plot is according to Hazard Ratios. (**B**) The plot is according to Odd Ratios.

**A**

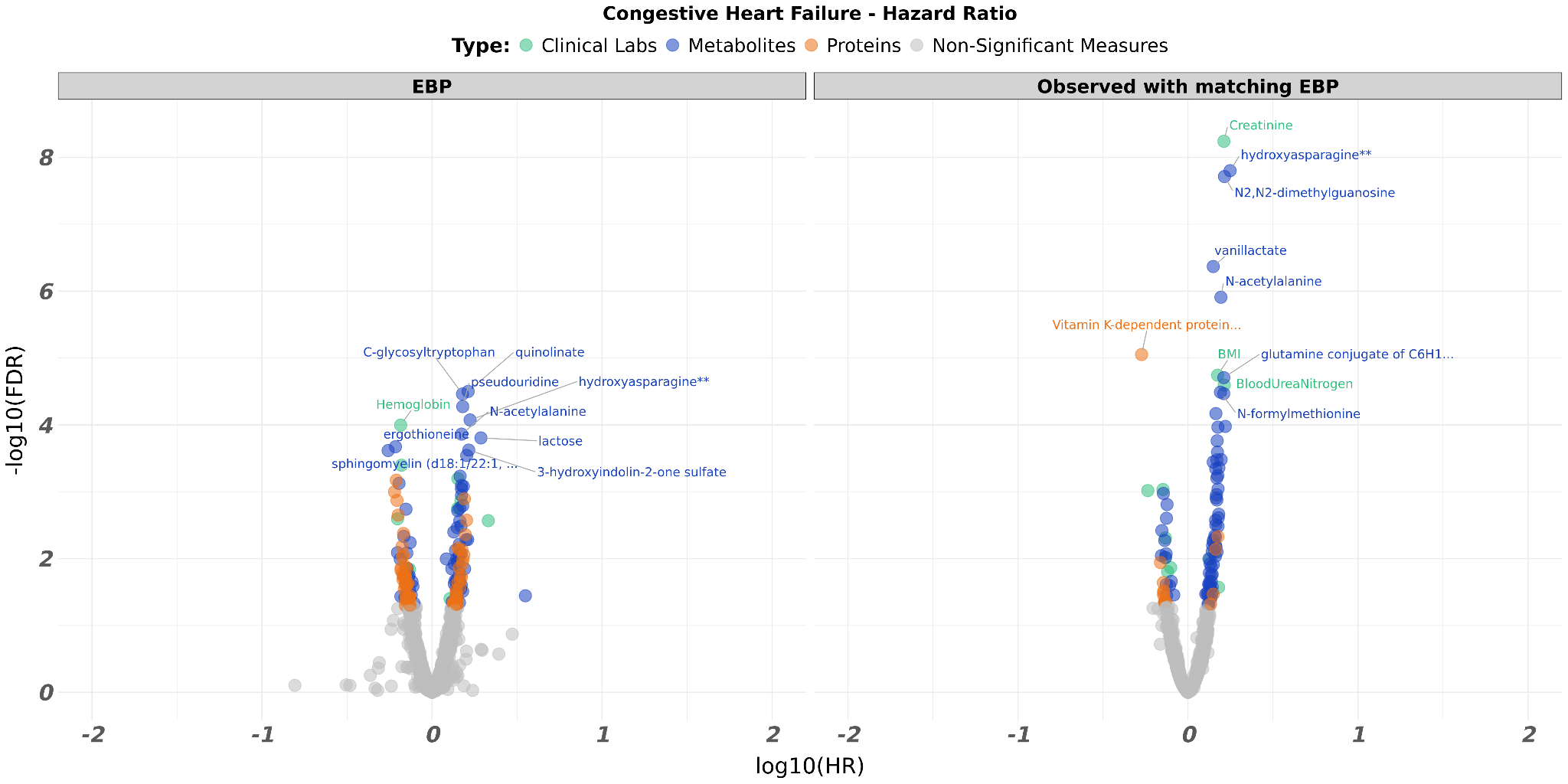

**B**

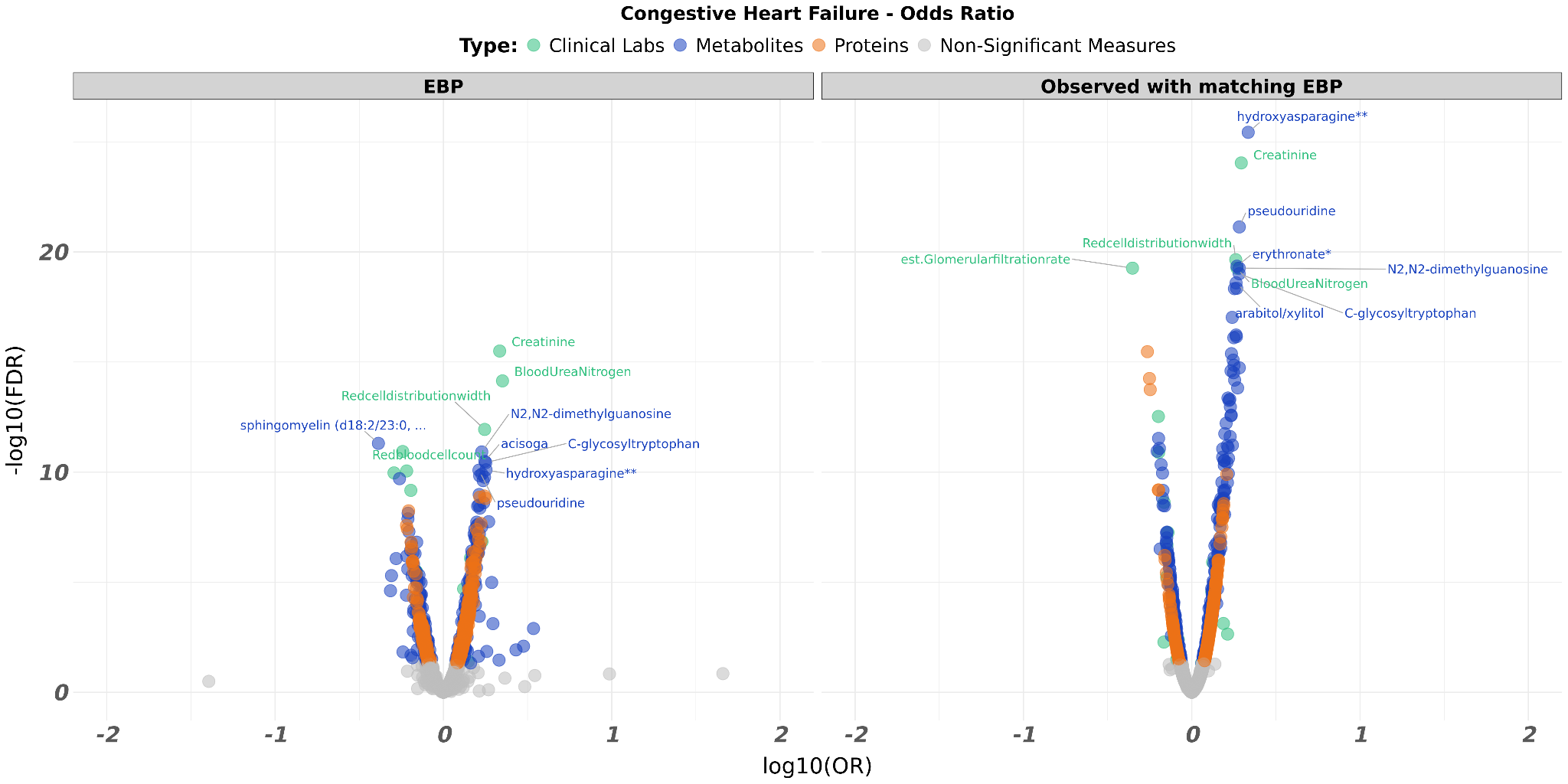

**Supplementary Figure 14.** Volcano plot comparing the EBP values to the matched observed values for congestive heart failure. (**A**) The plot is according to Hazard Ratios. (**B**) The plot is according to Odd Ratios.

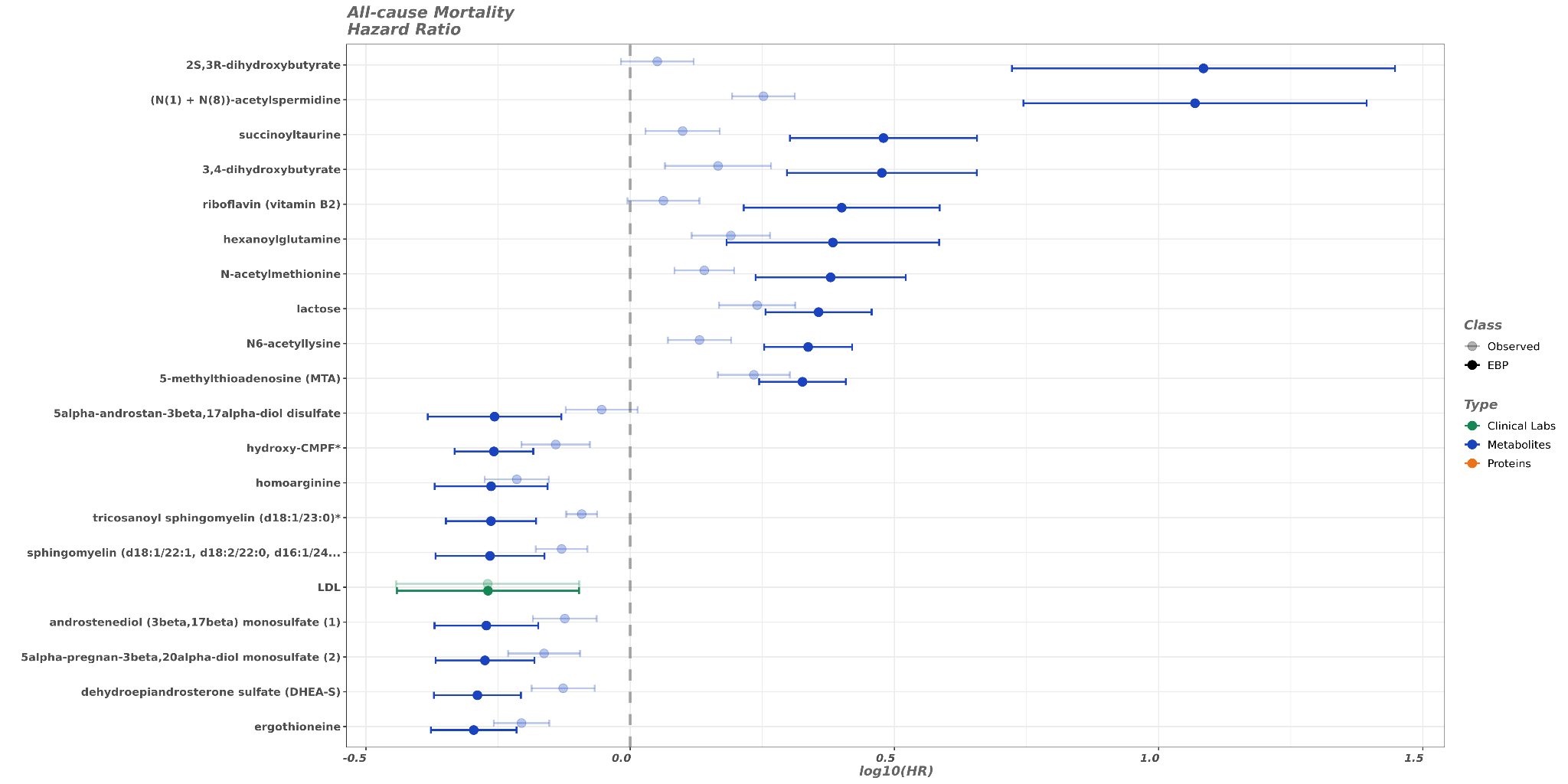

**Supplementary Figure 15.** Forest plot comparing the EBP values to the matched observed values for all-cause mortality.

**A**

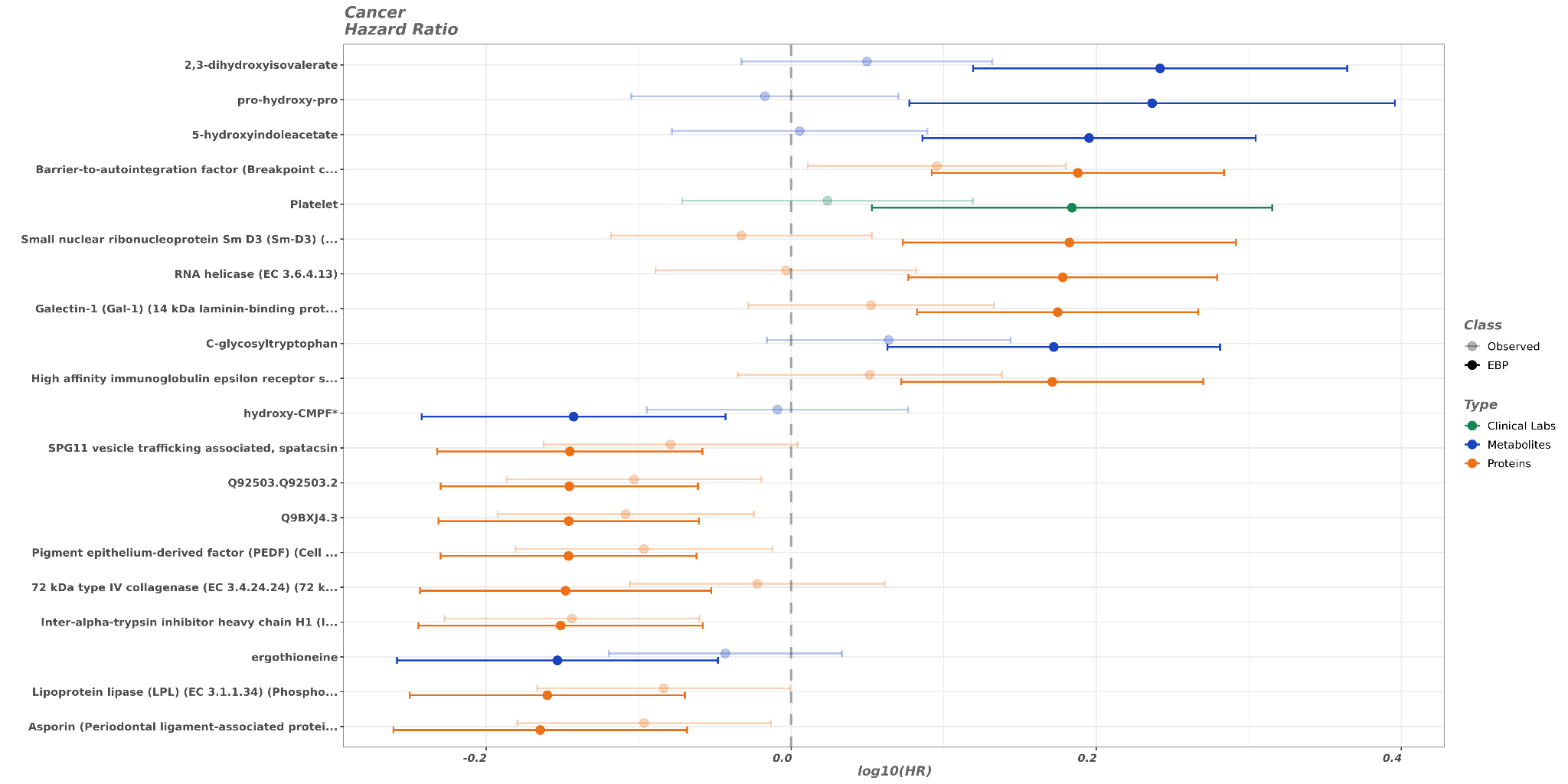

**B**

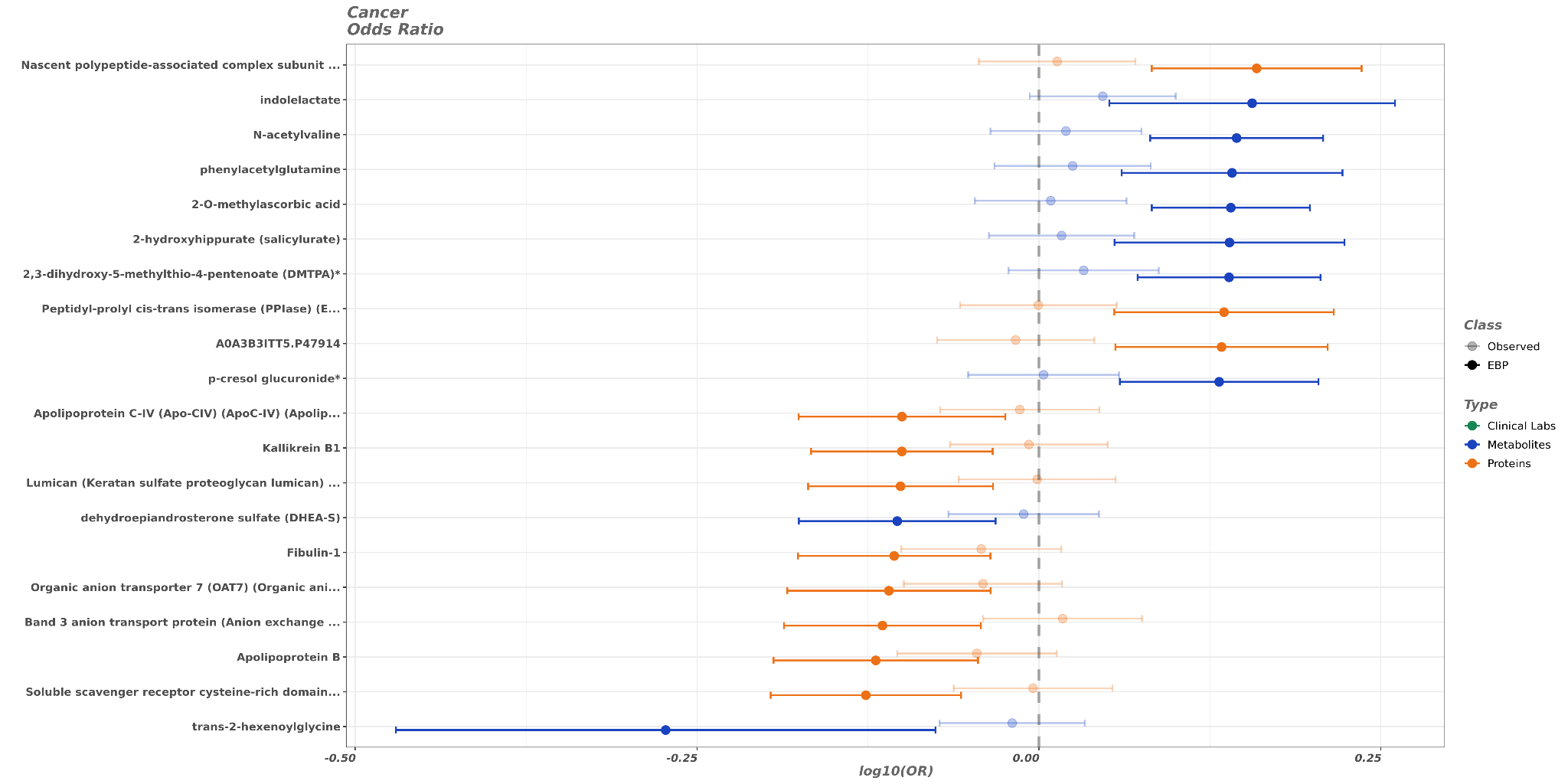

**Supplementary Figure 16.** Forest plot comparing the EBP values to the matched observed values for cancer. (**A**) The plot is according to Hazard Ratios. (**B**) The plot is according to Odd Ratios.

**A**

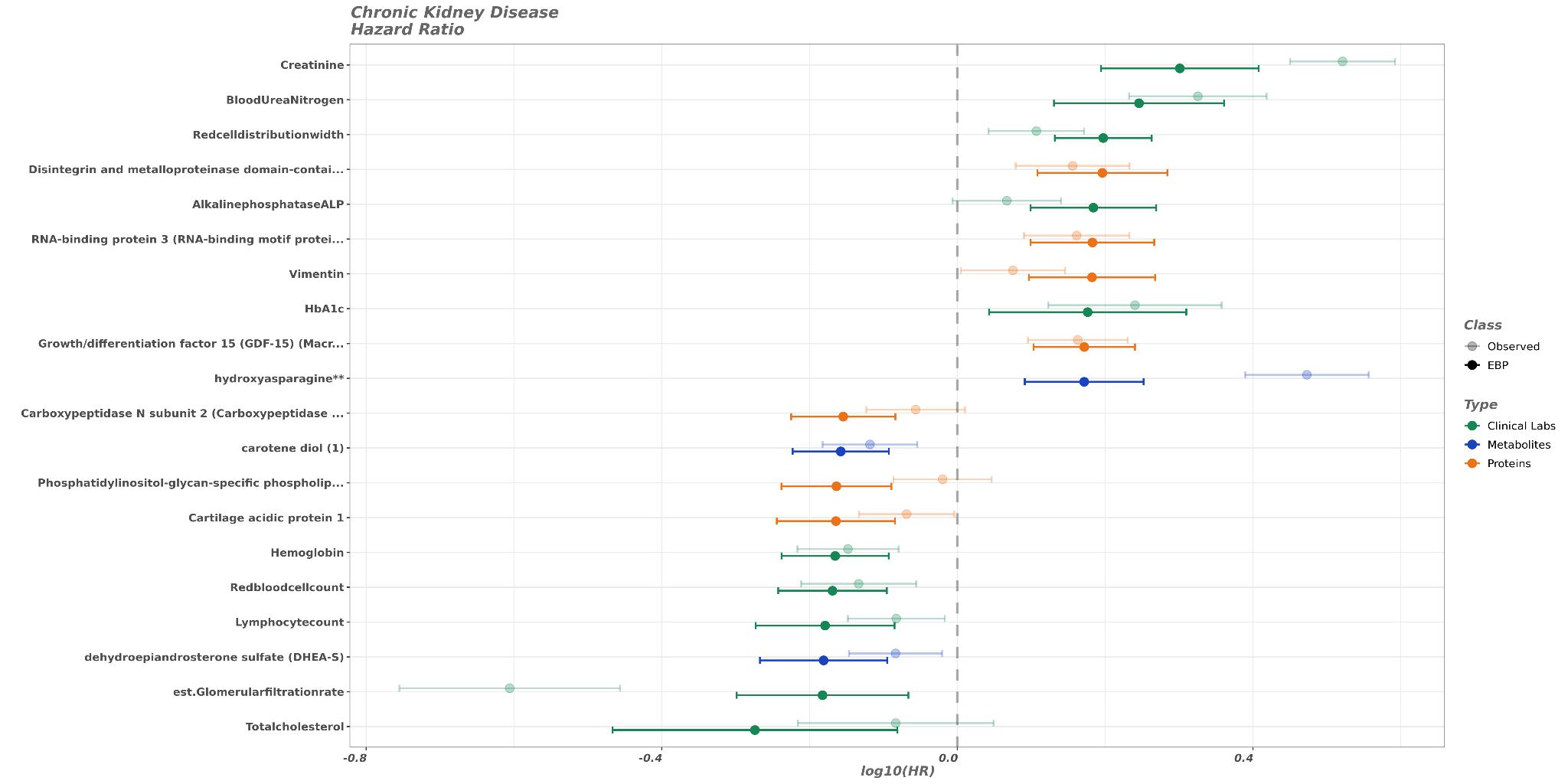

**B**

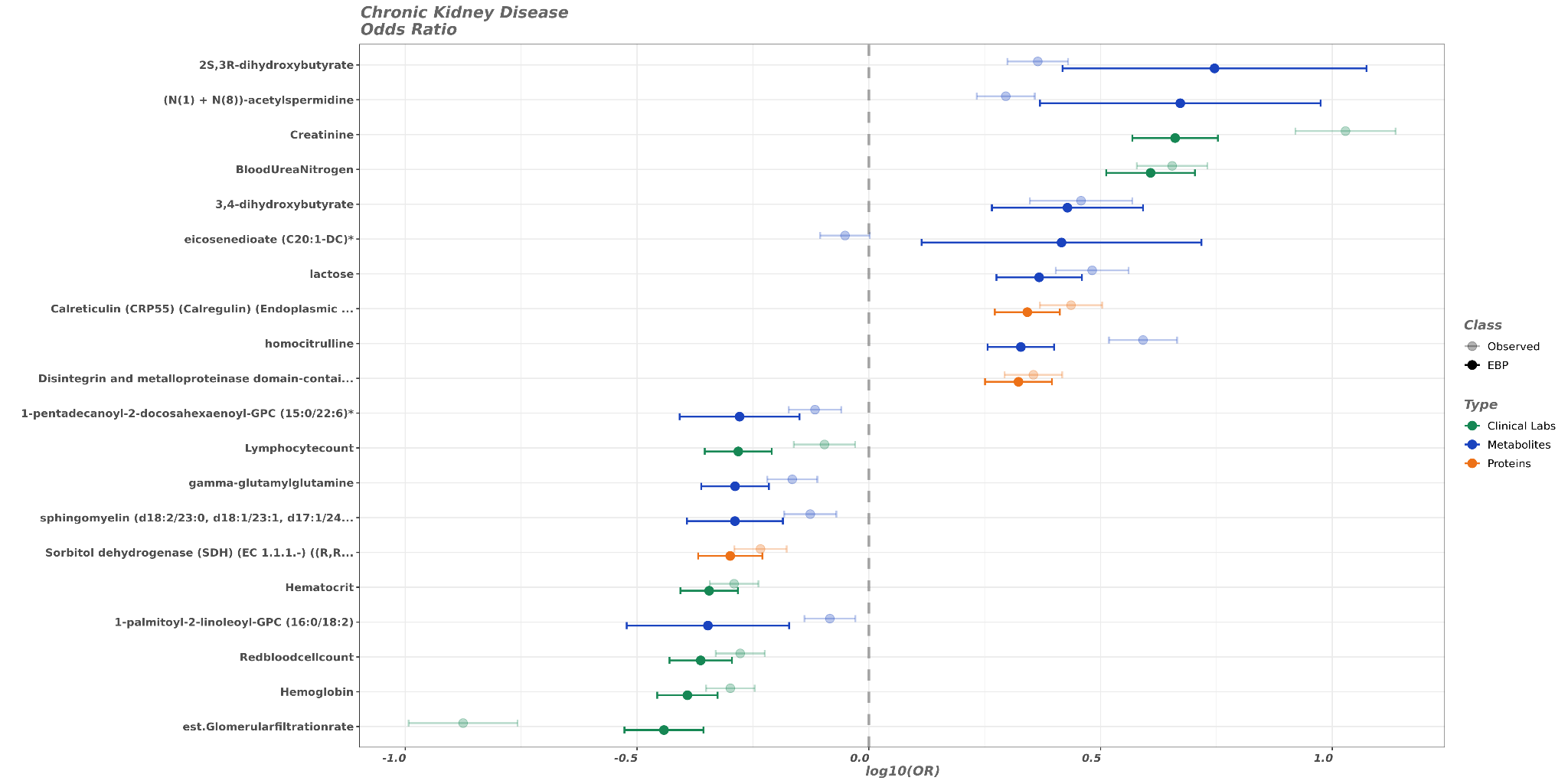

**Supplementary Figure 17.** Forest plot comparing the EBP values to the matched observed values for chronic kidney disease. (**A**) The plot is according to Hazard Ratios. (**B**) The plot is according to Odd Ratios.

**A**

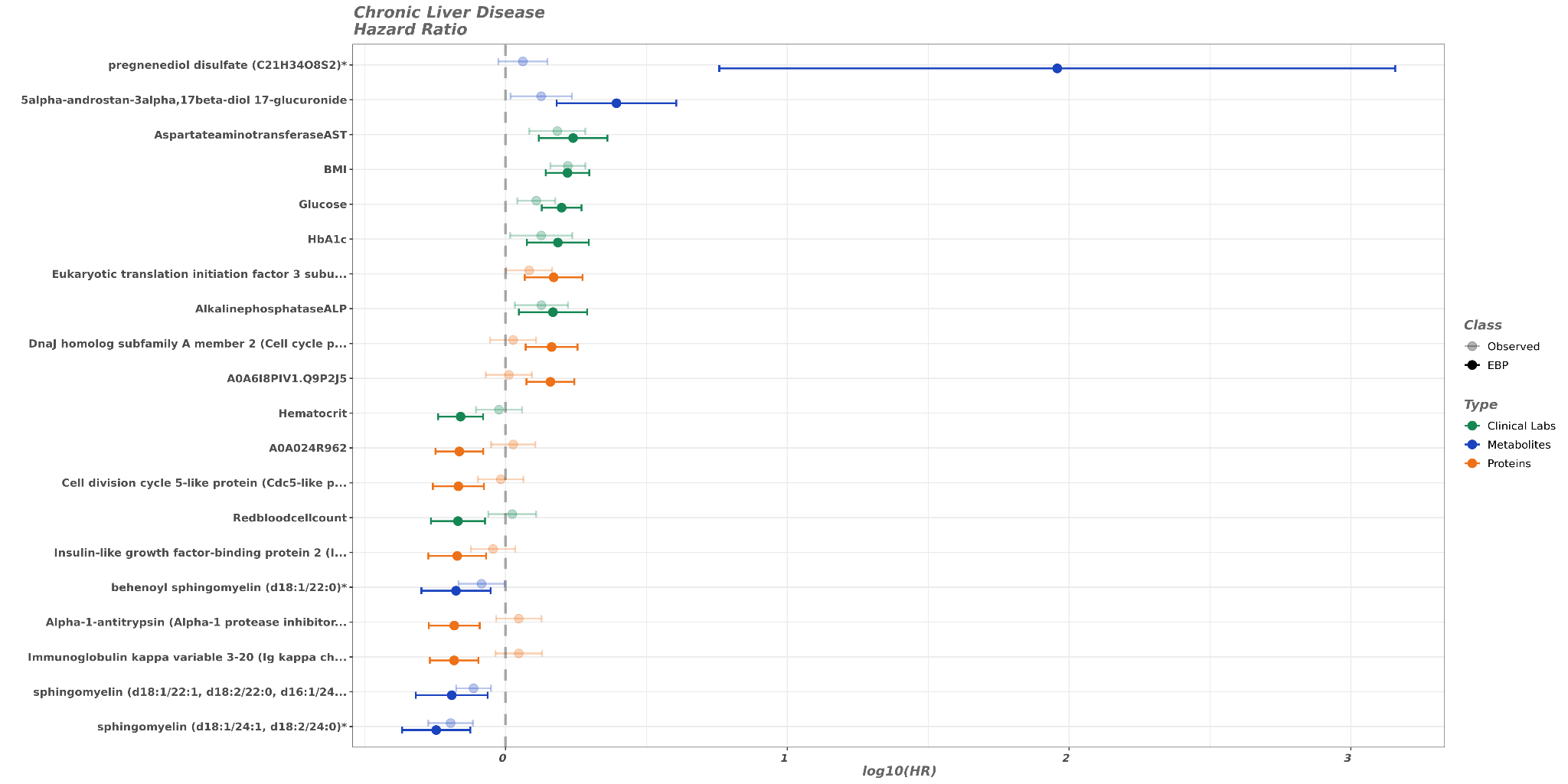

**B**

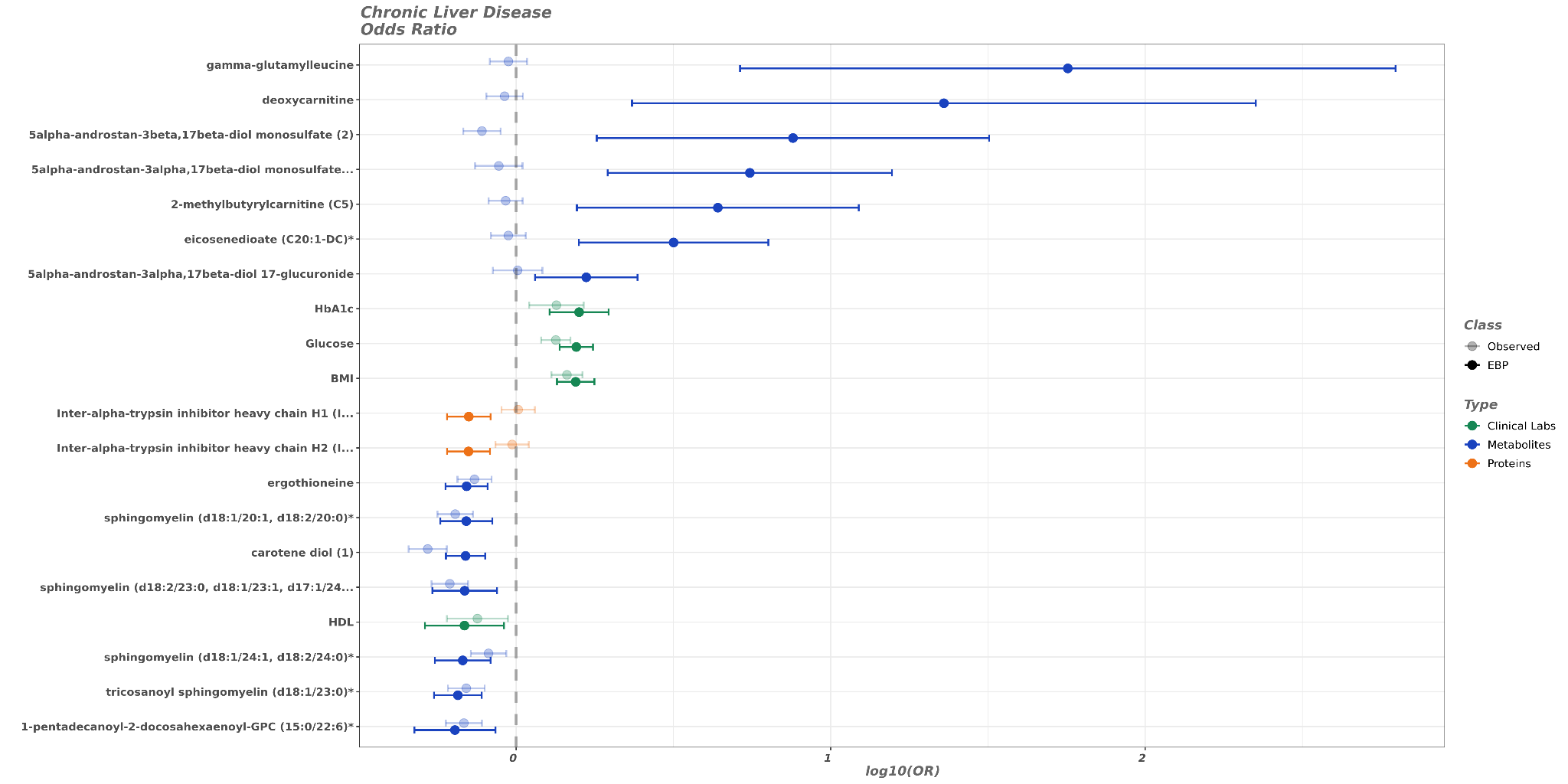

**Supplementary Figure 18.** Forest plot comparing the EBP values to the matched observed values for chronic liver disease. (**A**) The plot is according to Hazard Ratios. (**B**) The plot is according to Odd Ratios.

**A**

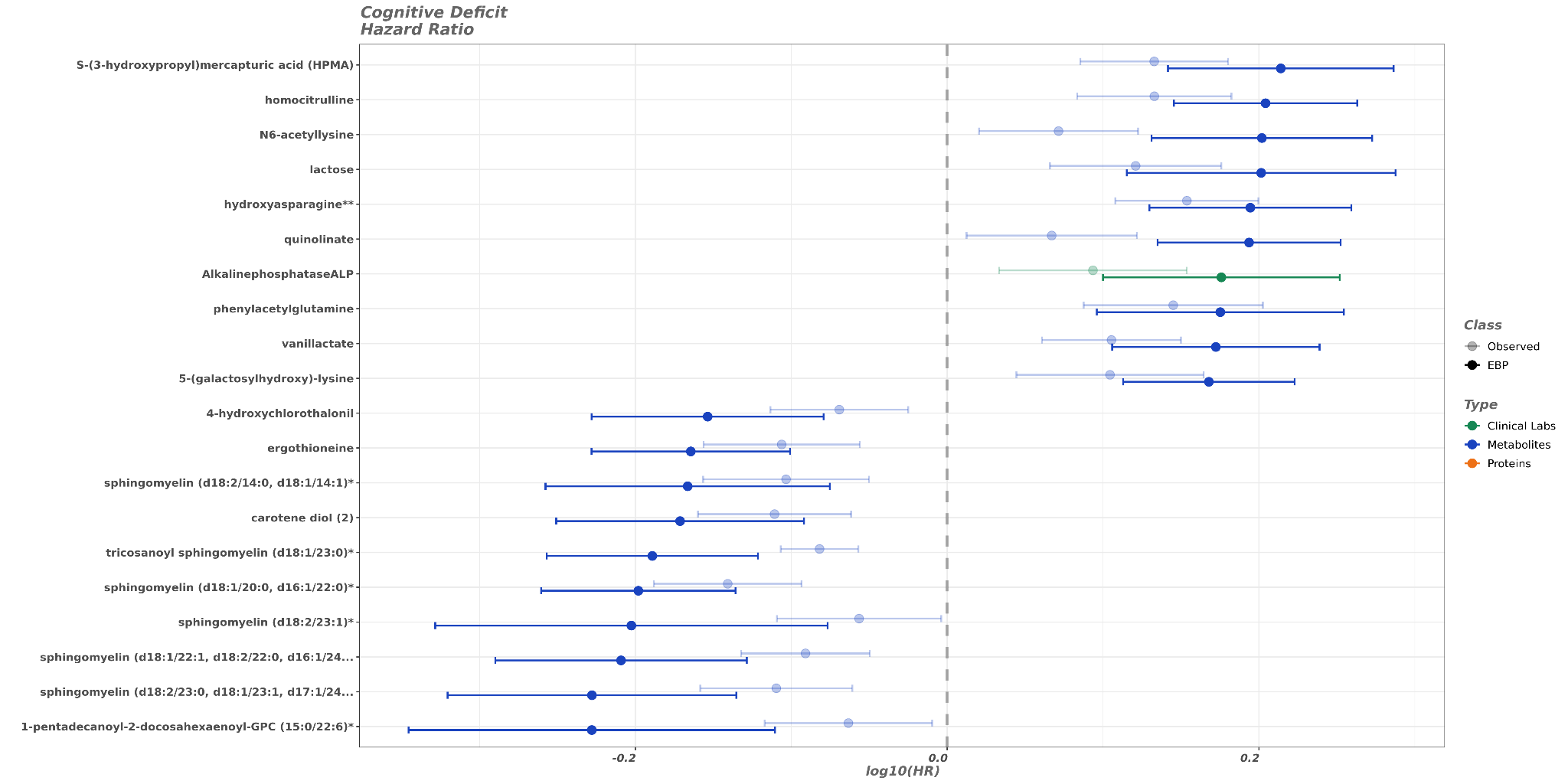

**B**

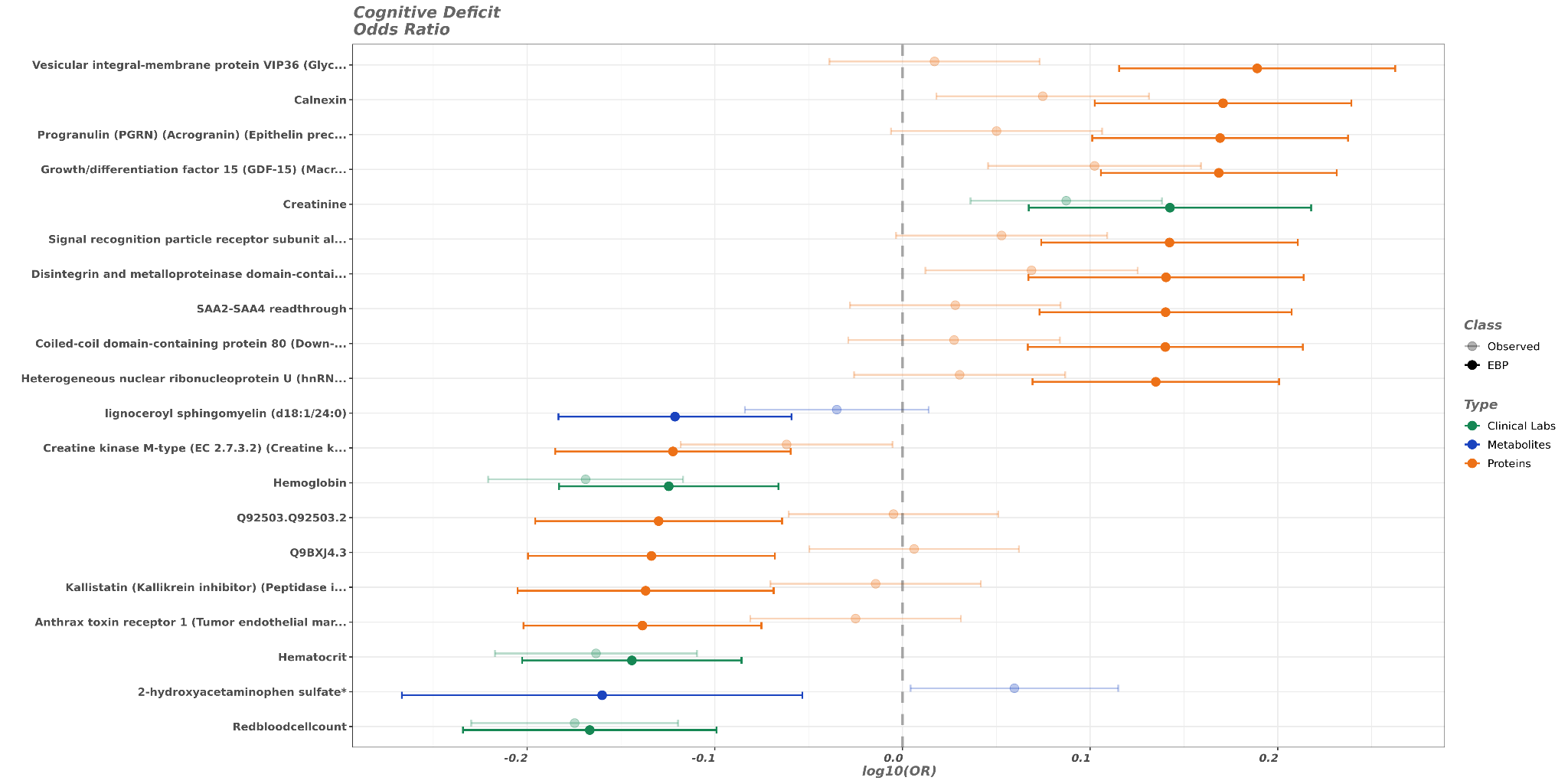

**Supplementary Figure 19.** Forest plot comparing the EBP values to the matched observed values for cognitive deficit. (**A**) The plot is according to Hazard Ratios. (**B**) The plot is according to Odd Ratios.

**A**

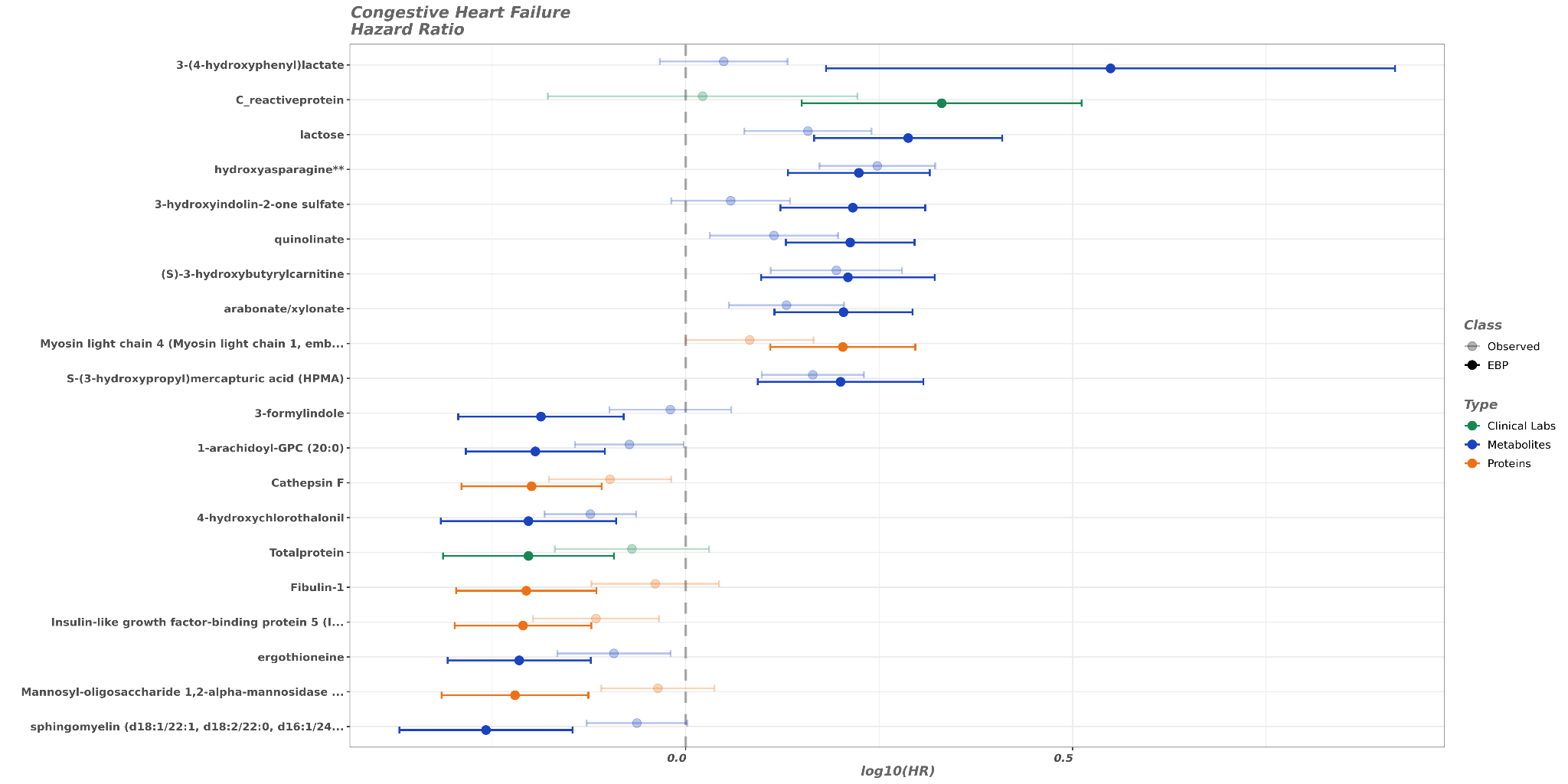

**B**

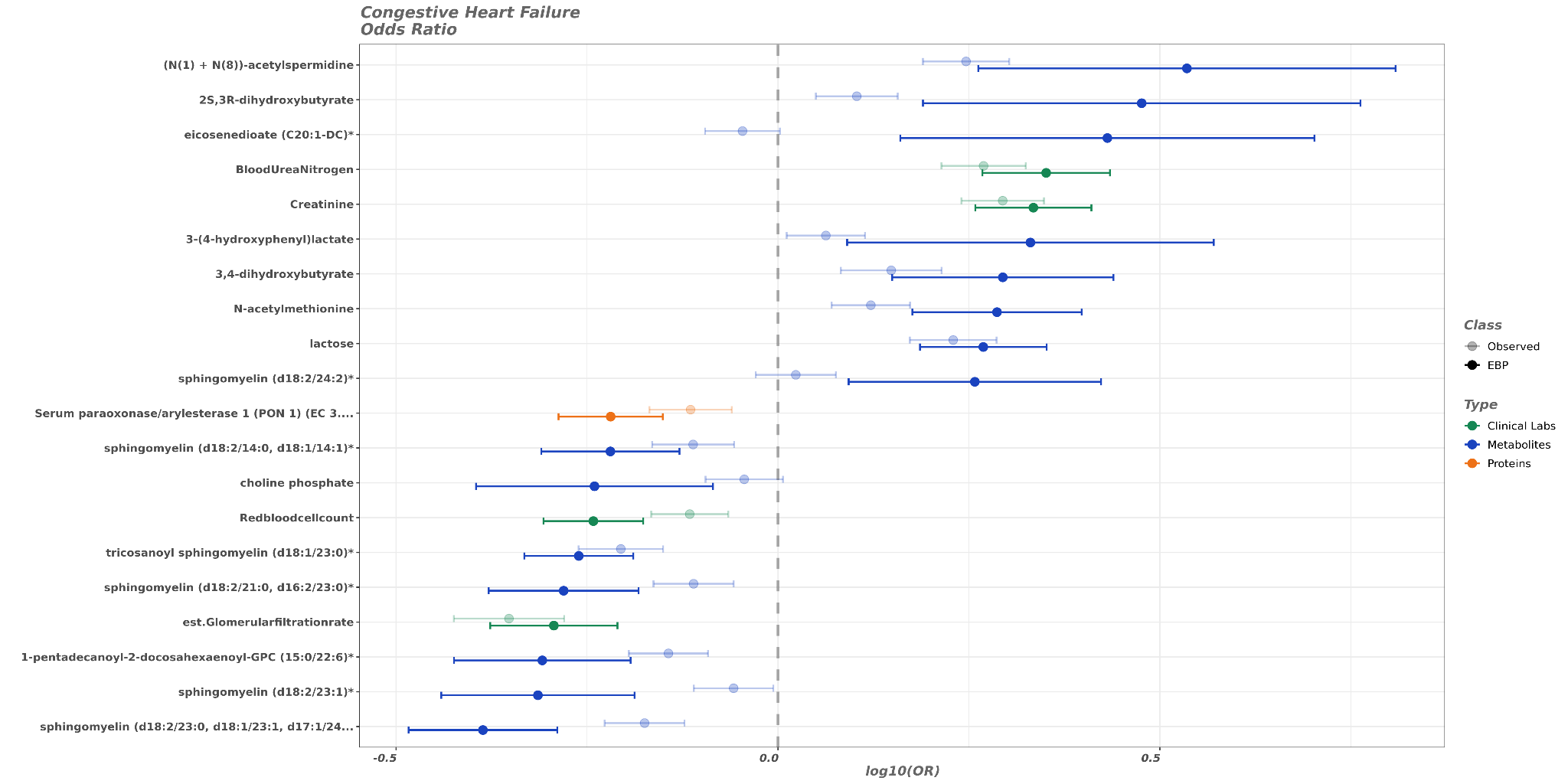

**Supplementary Figure 20.** Forest plot comparing the EBP values to the matched observed values for congestive heart failure. (**A**) The plot is according to Hazard Ratios. (**B**) The plot is according to Odd Ratios.

**A**

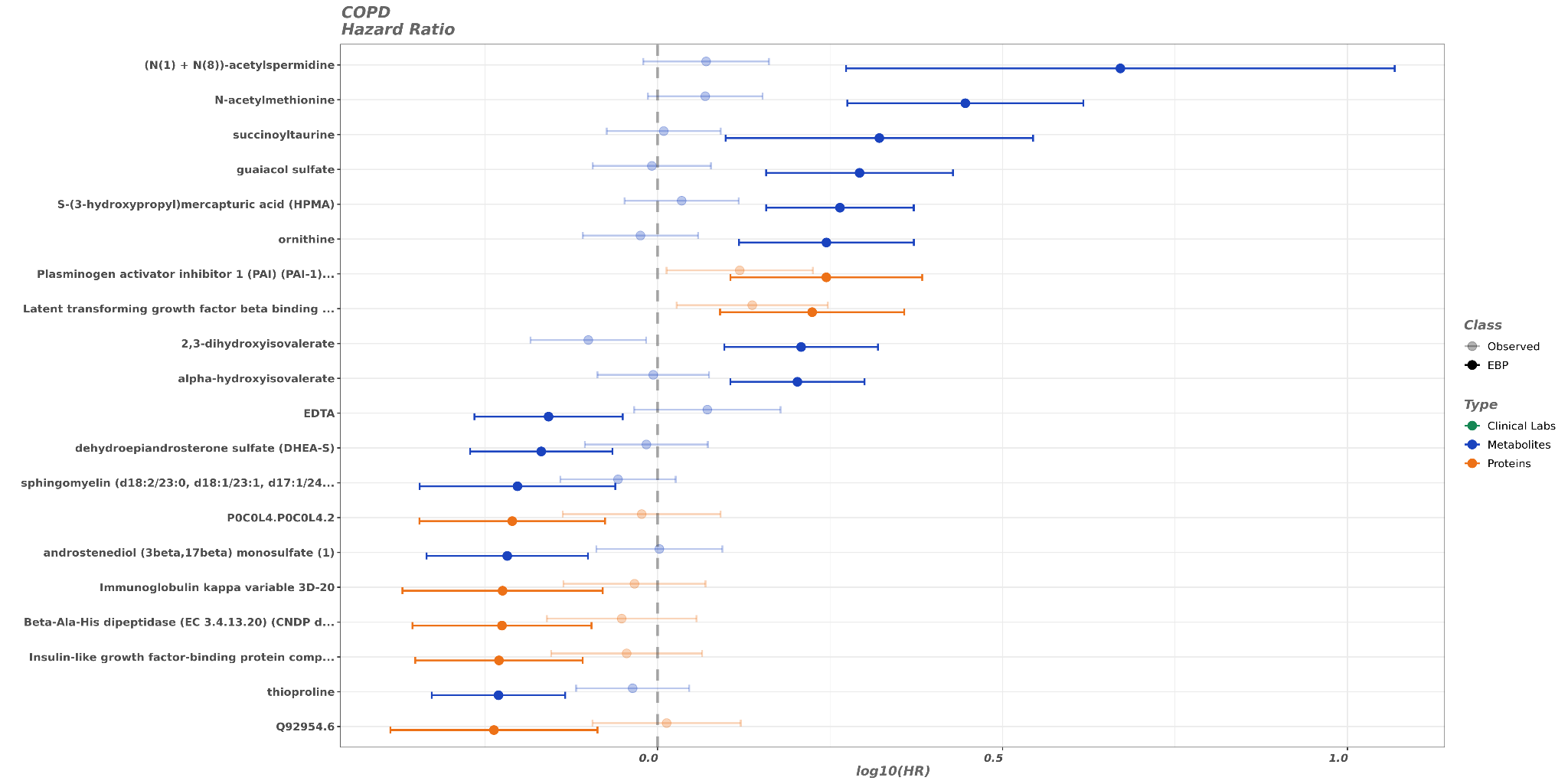

**B**

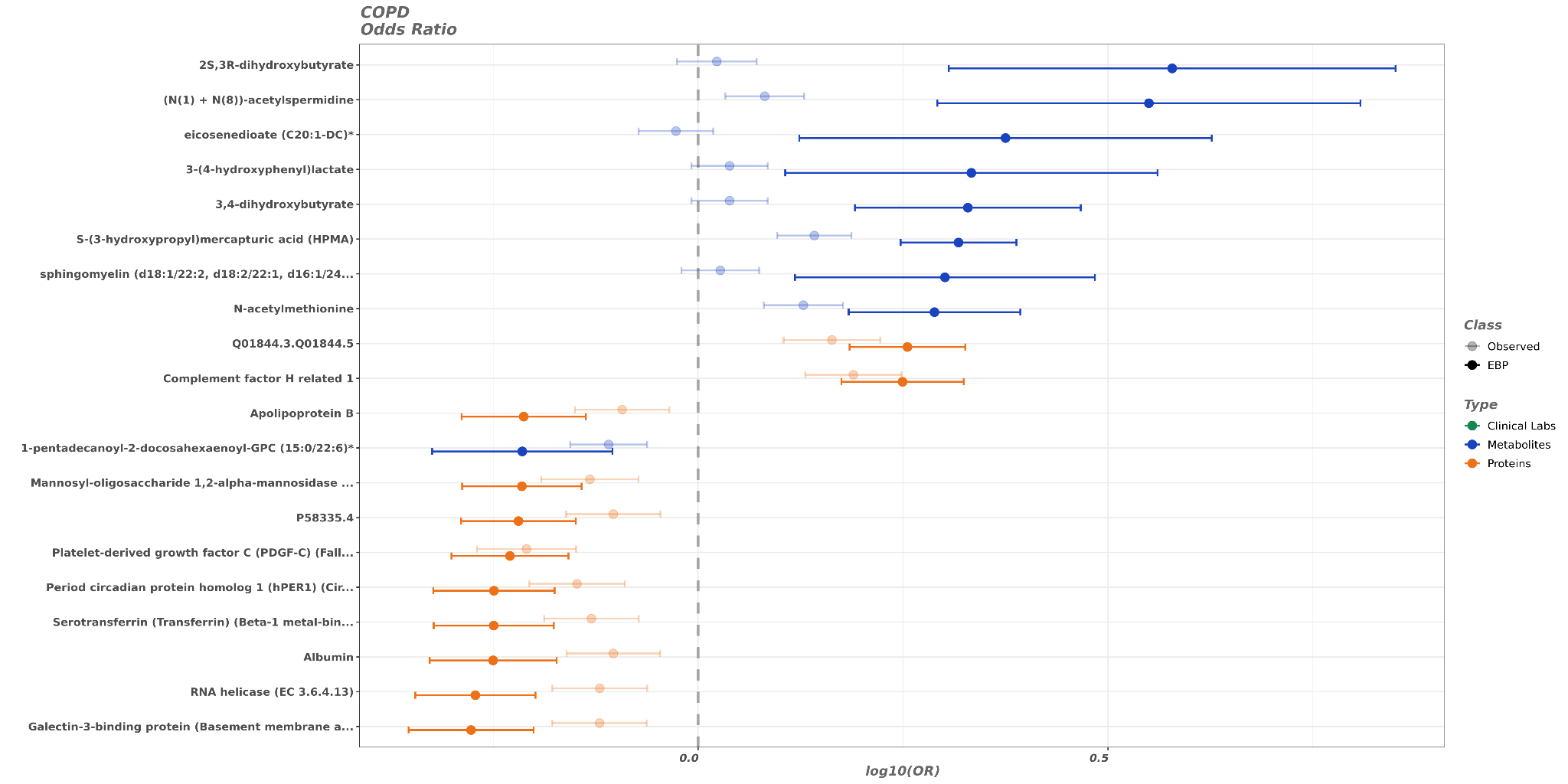

**Supplementary Figure 21.** Forest plot comparing the EBP values to the matched observed values for COPD. (**A**) The plot is according to Hazard Ratios. (**B**) The plot is according to Odd Ratios.

**A**

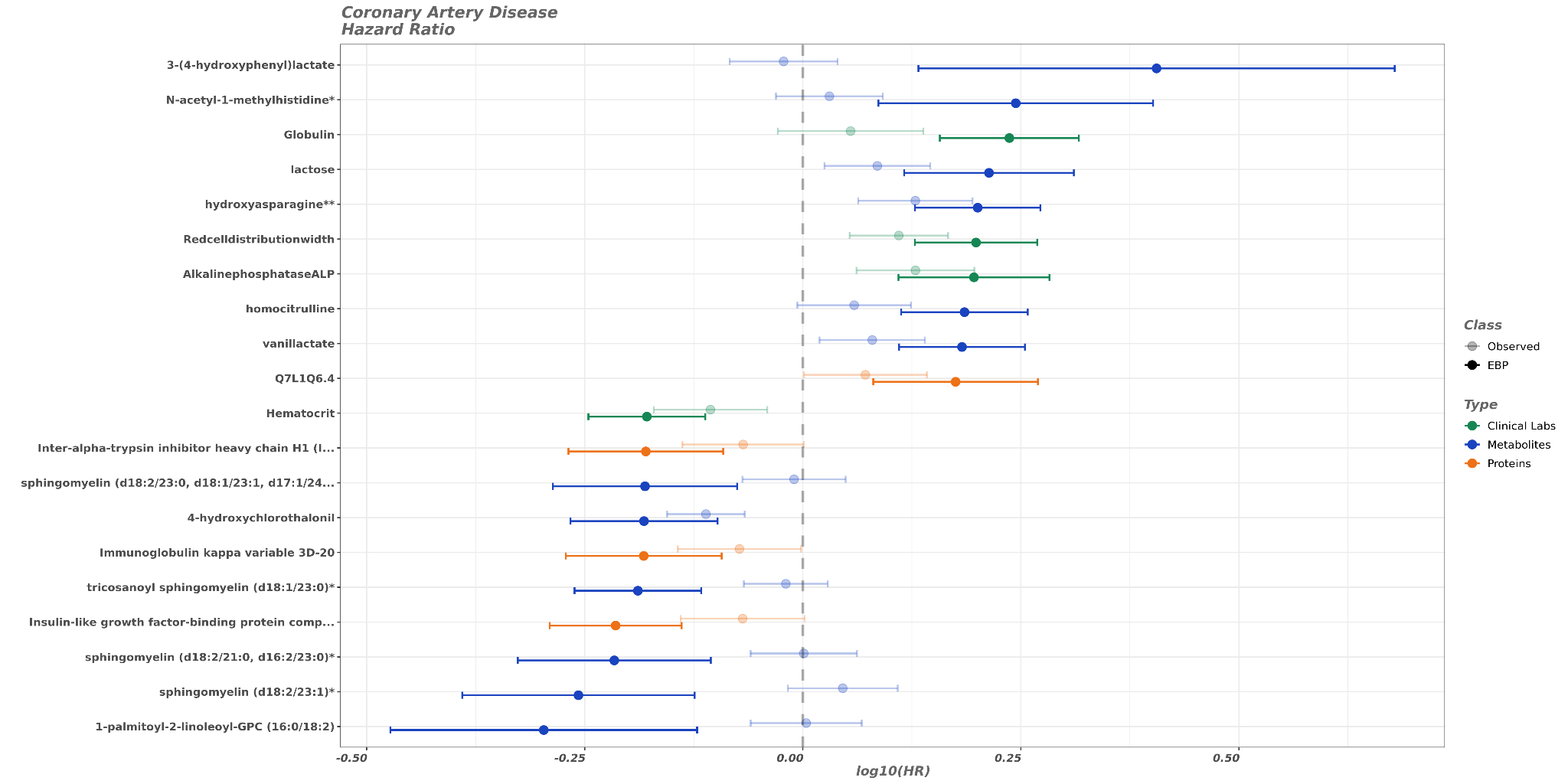

**B**

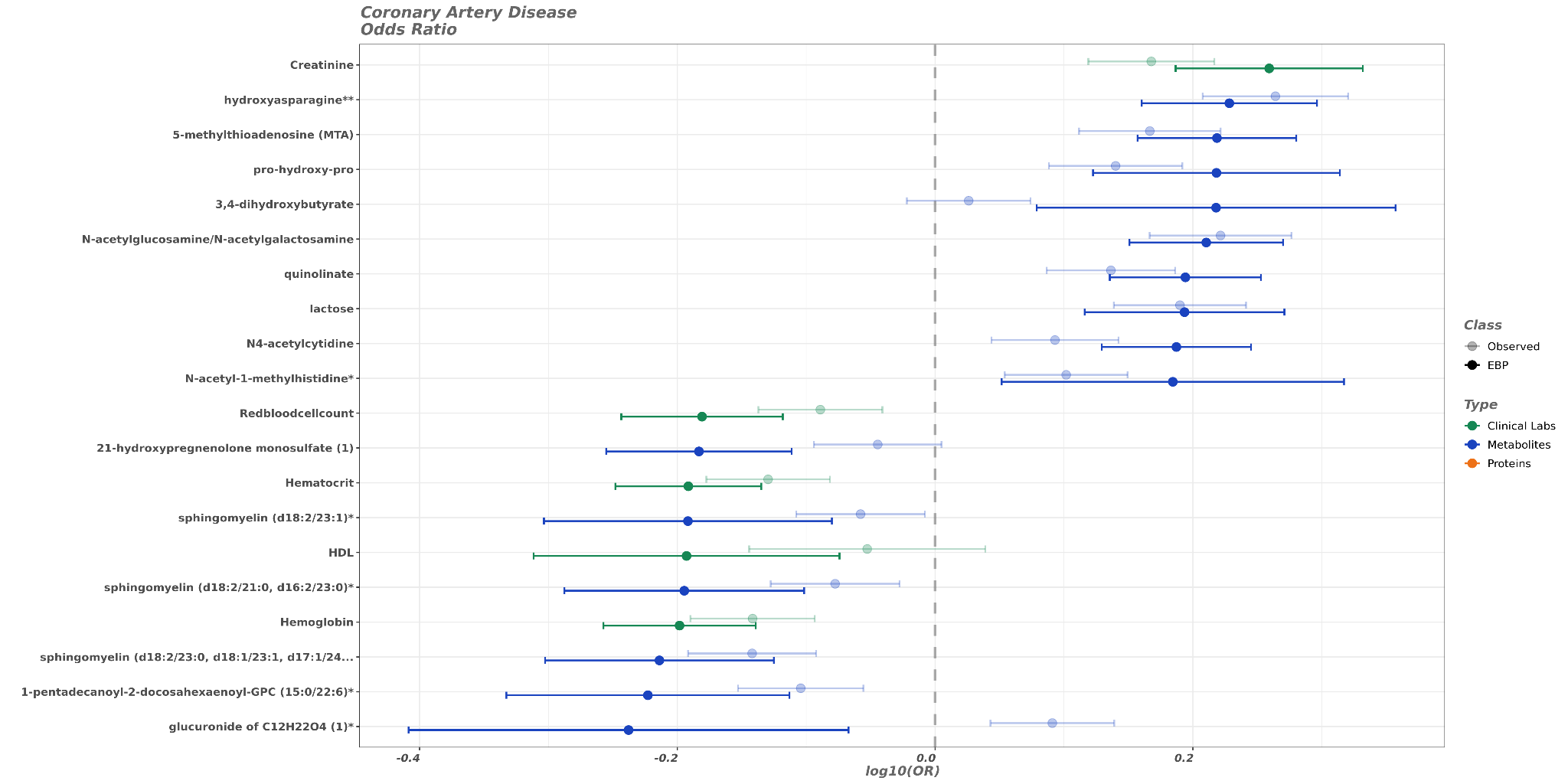

**Supplementary Figure 22.** Forest plot comparing the EBP values to the matched observed values for coronary artery disease. (**A**) The plot is according to Hazard Ratios. (**B**) The plot is according to Odd Ratios.

**A**

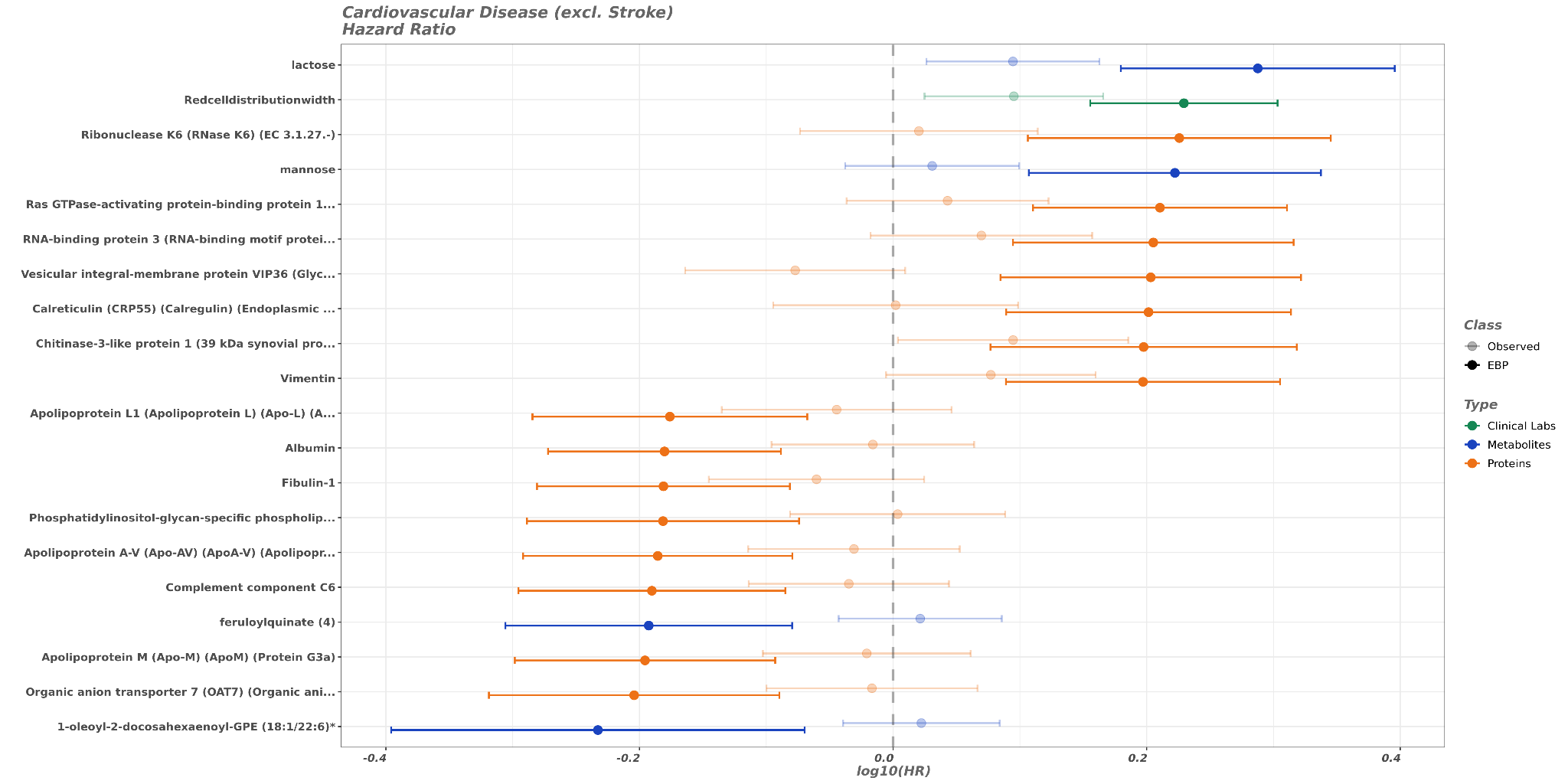

**B**

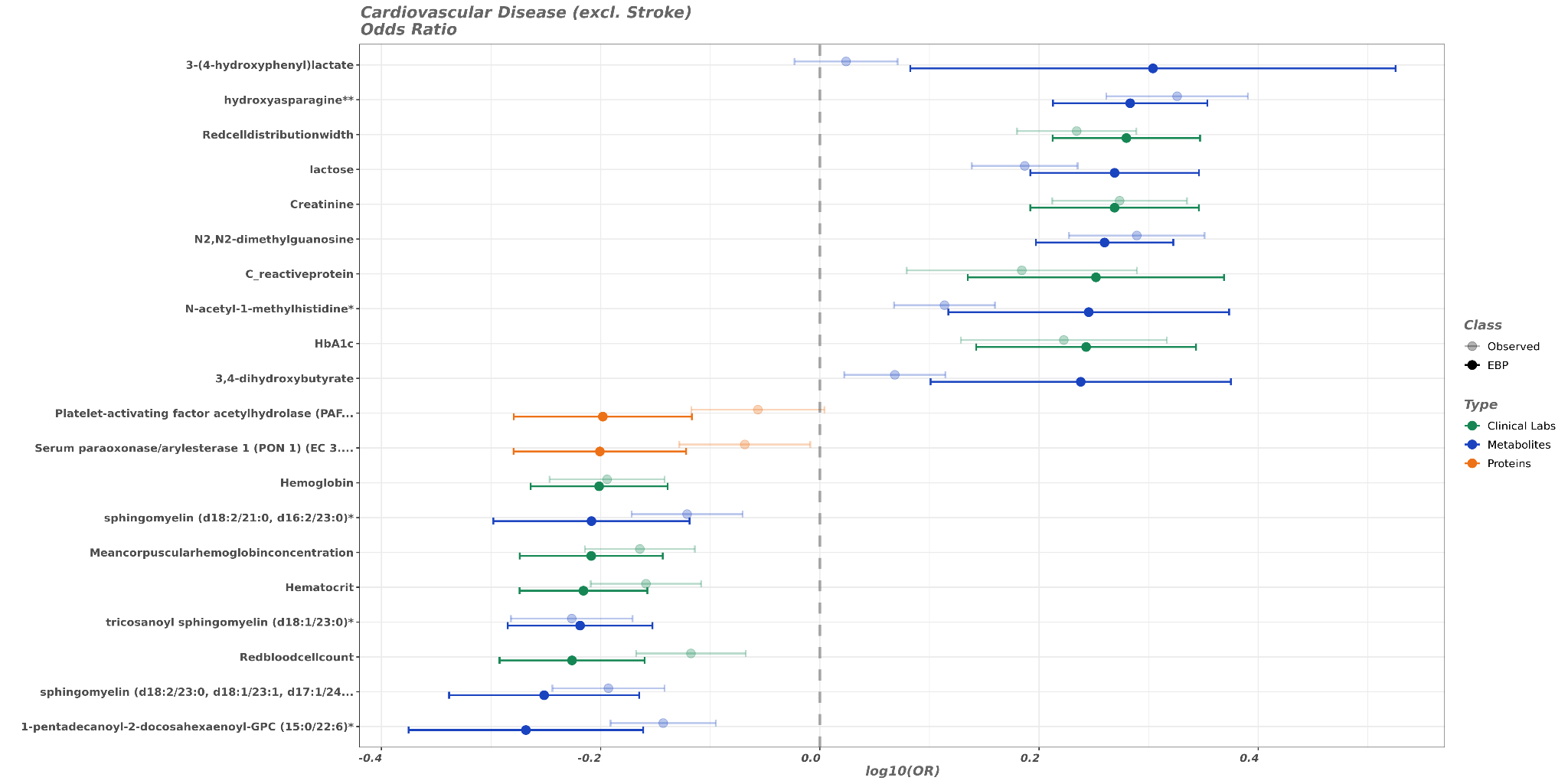

**Supplementary Figure 23.** Forest plot comparing the EBP values to the matched observed values for cardiovascular disease excluding stroke. (**A**) The plot is according to Hazard Ratios. (**B**) The plot is according to Odd Ratios.

**A**

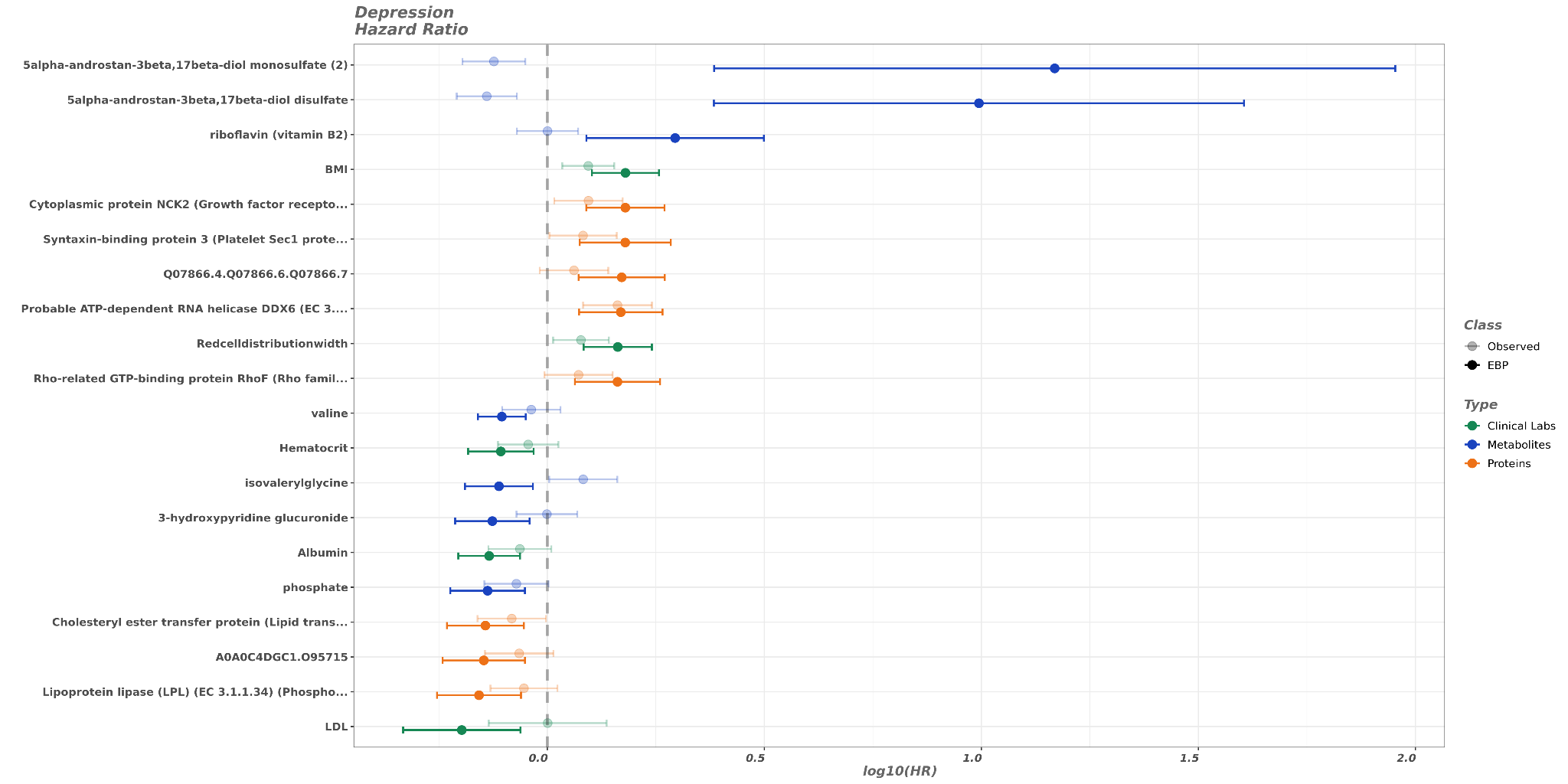

**B**

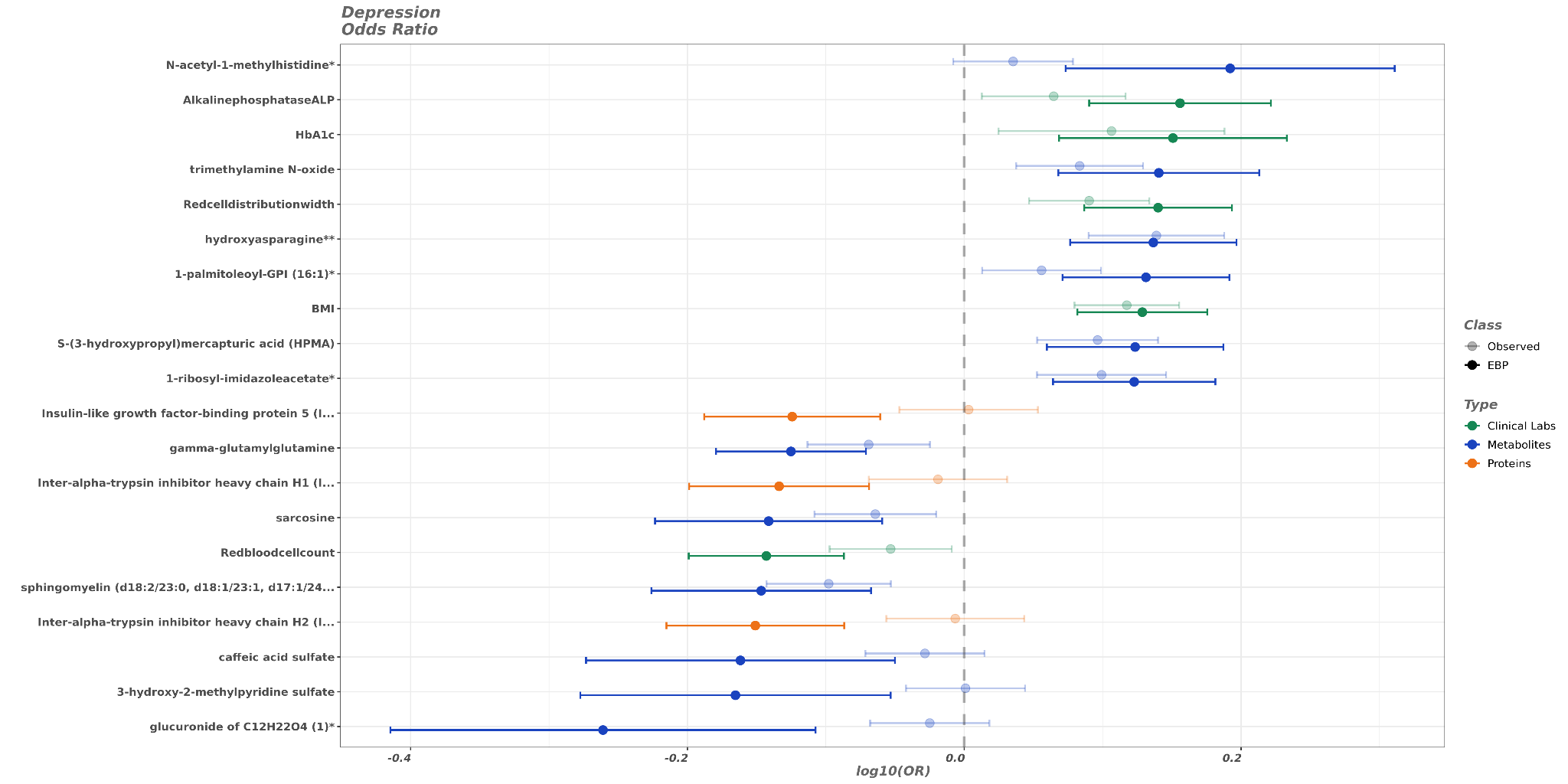

**Supplementary Figure 24.** Forest plot comparing the EBP values to the matched observed values for depression. (**A**) The plot is according to Hazard Ratios. (**B**) The plot is according to Odd Ratios.

**A**

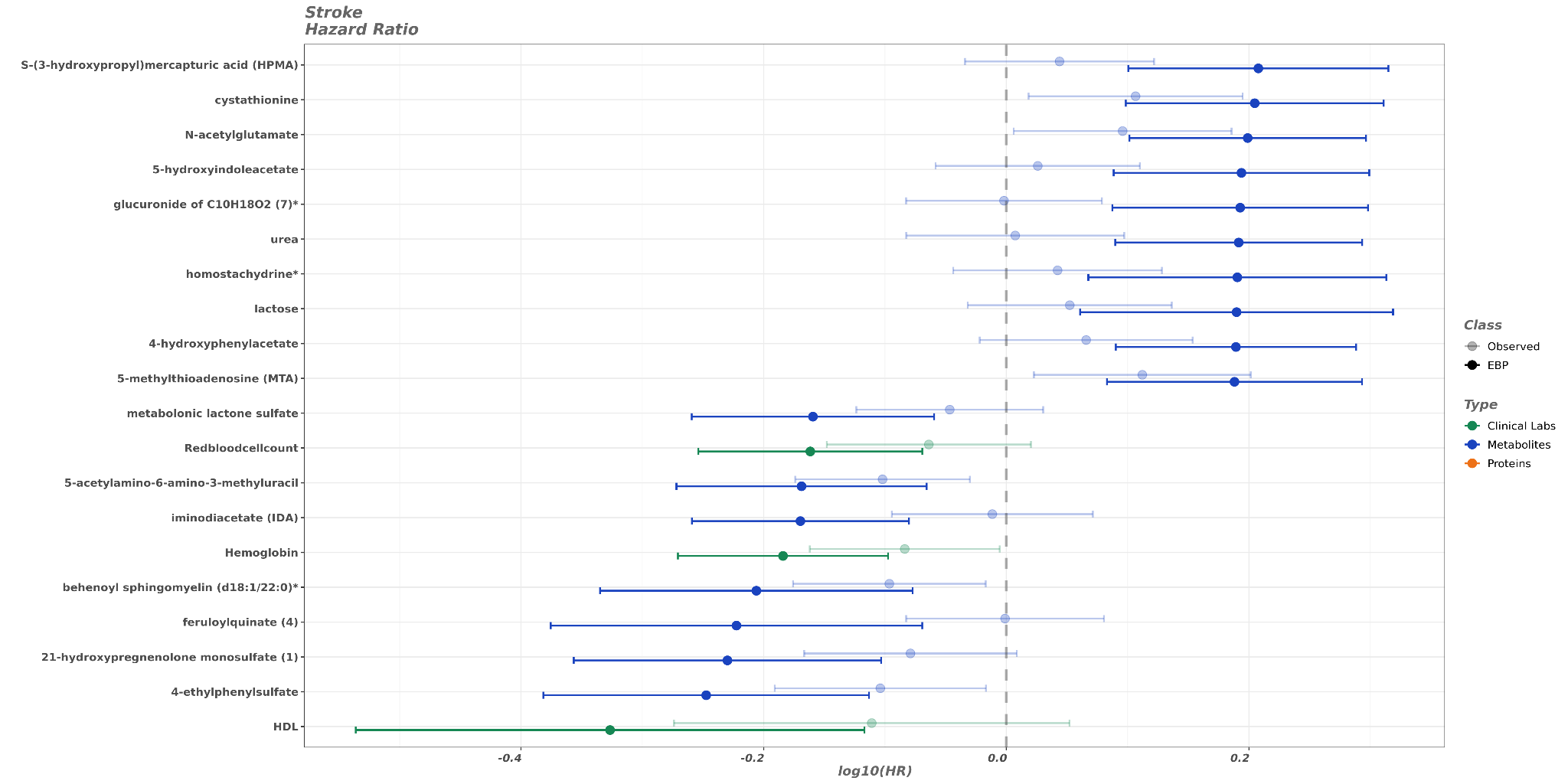

**B**

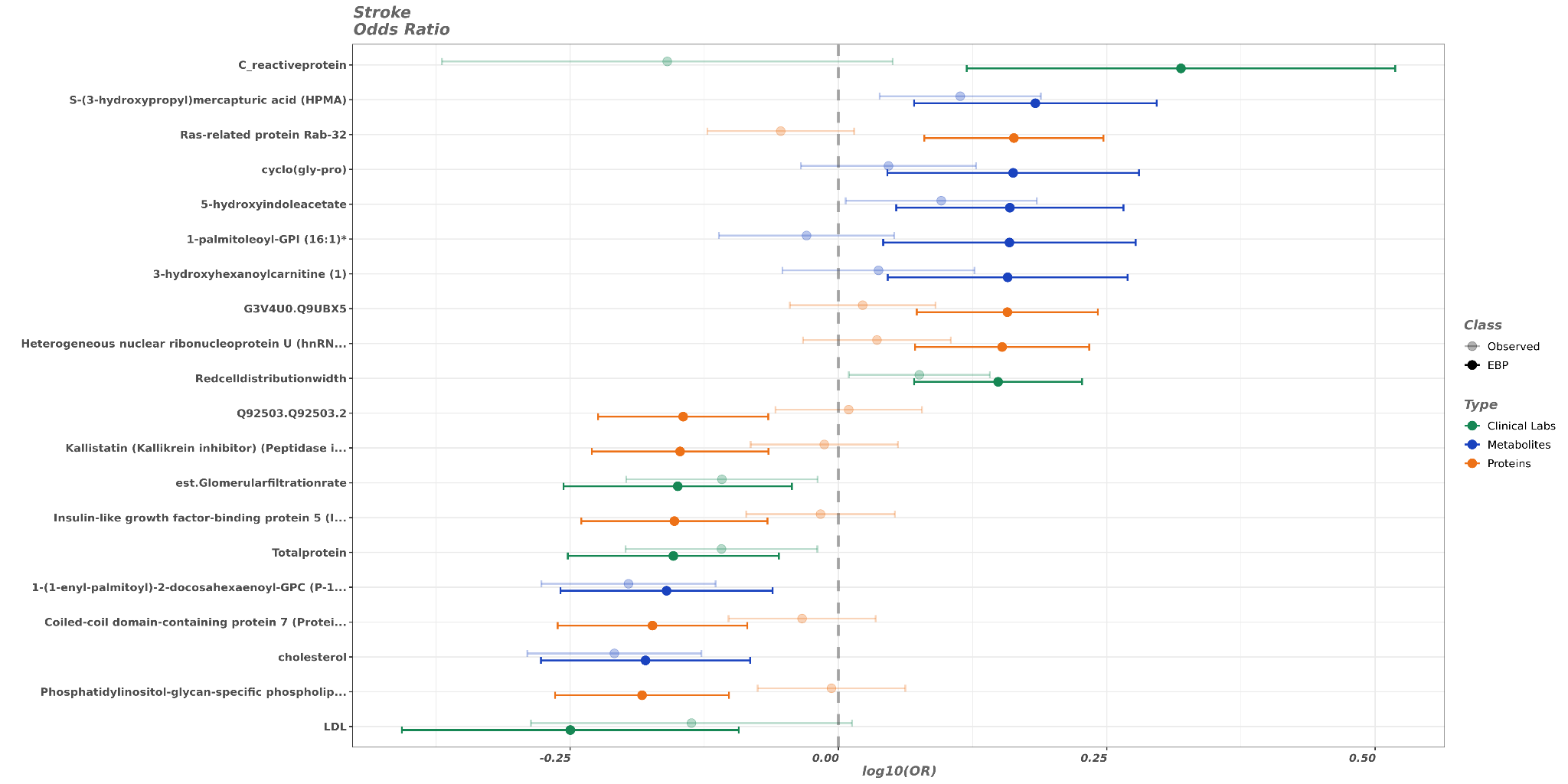

**Supplementary Figure 25.** Forest plot comparing the EBP values to the matched observed values for stroke. (**A**) The plot is according to Hazard Ratios. (**B**) The plot is according to Odd Ratios.

**A**

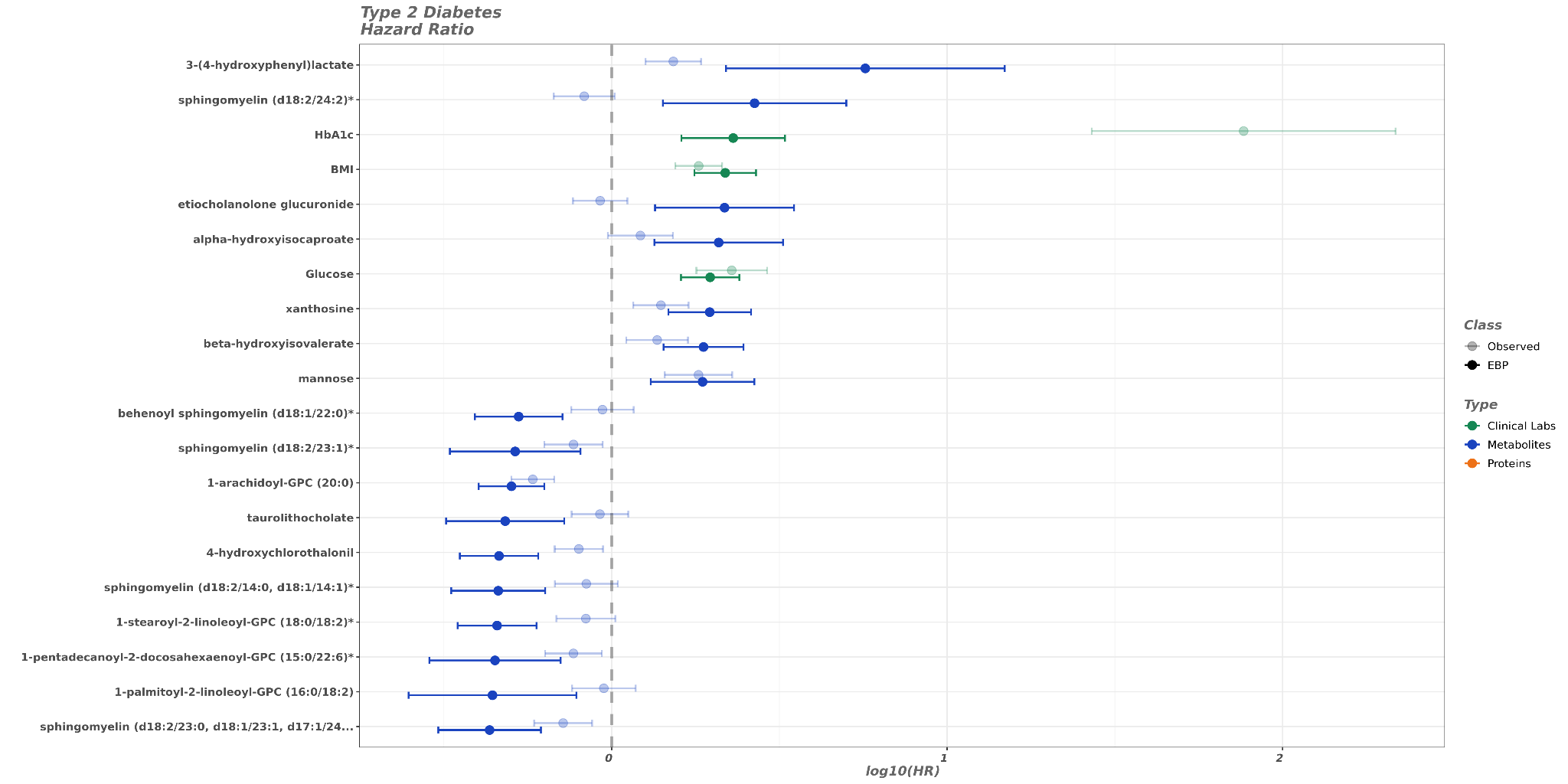

**B**

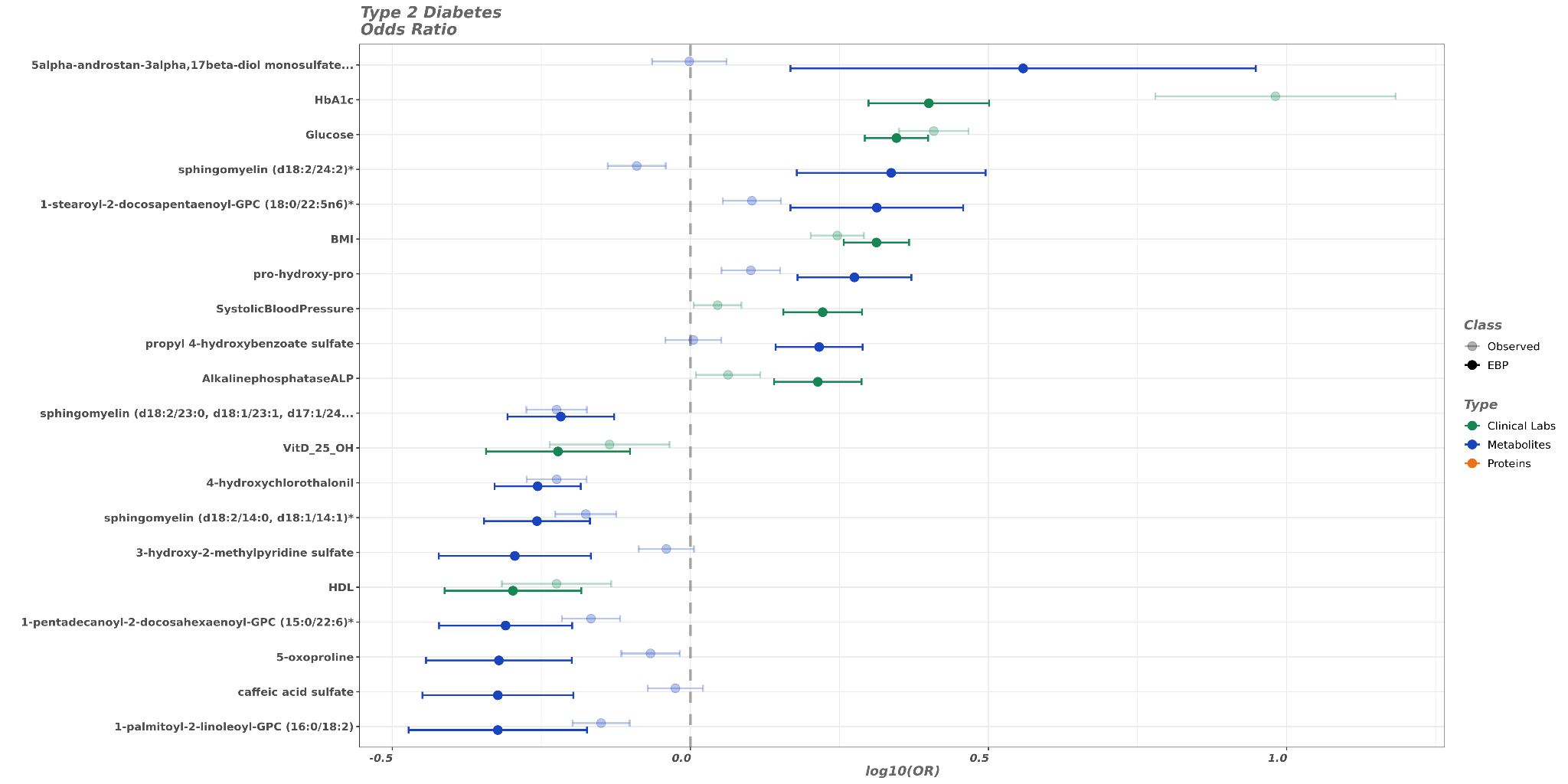

**Supplementary Figure 26.** Forest plot comparing the EBP values to the matched observed values for Type 2 diabetes. (**A**) The plot is according to Hazard Ratios. (**B**) The plot is according to Odd Ratios.

**A**

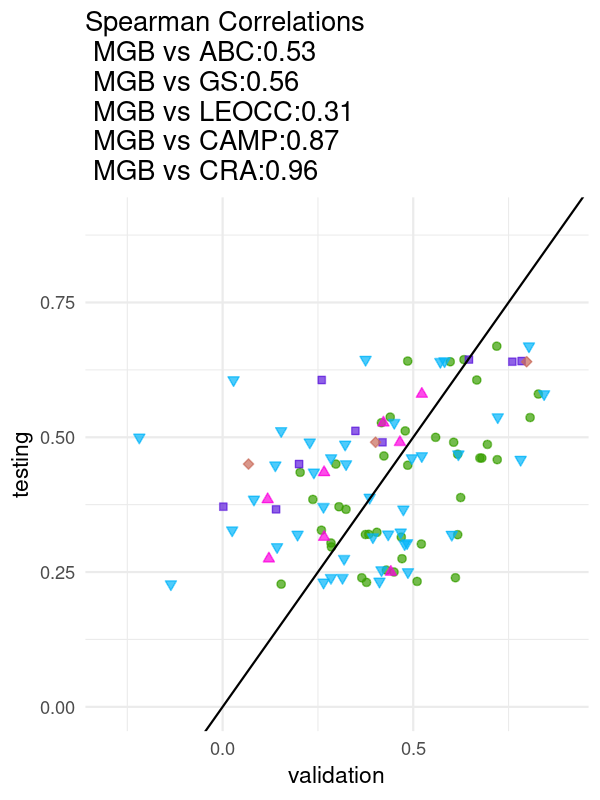

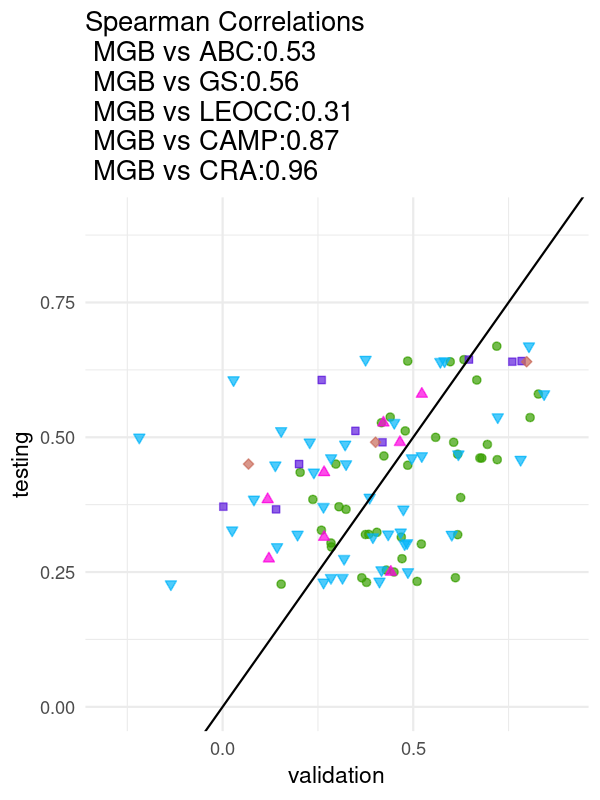

**B**

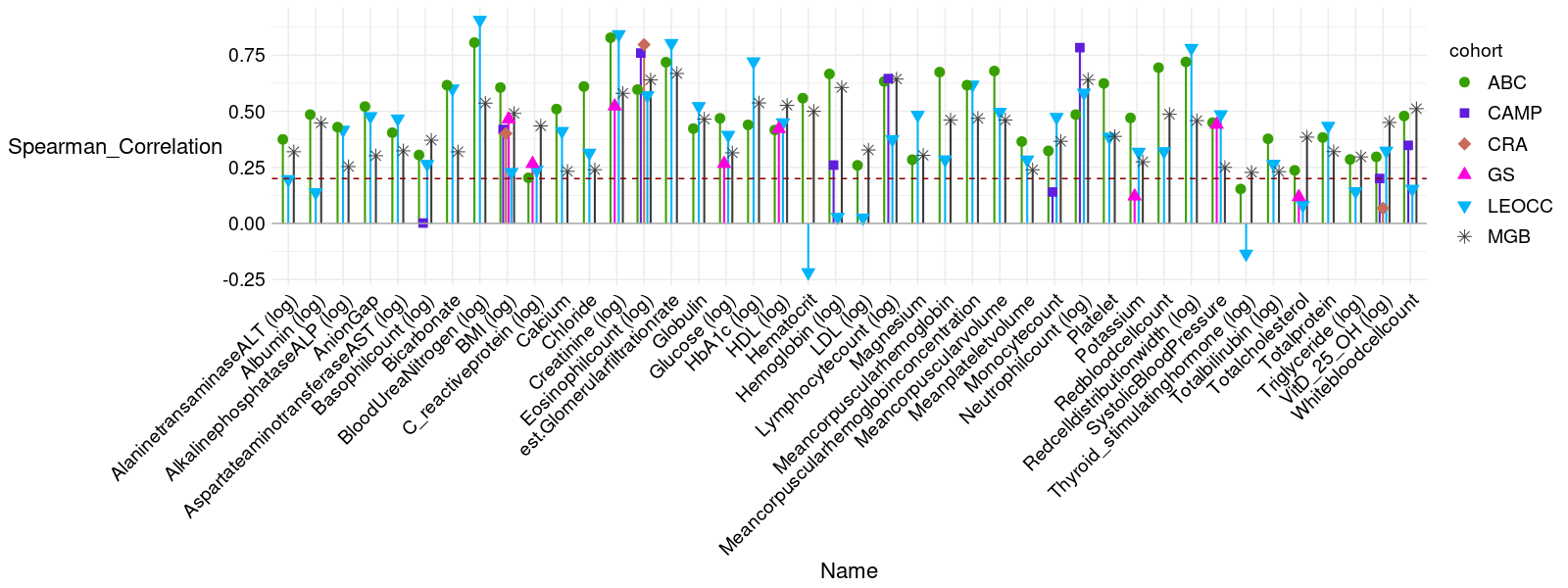

**Supplementary Figure 27. Comparison of validation and testing performances for clinical measures across five independent cohorts**. (**A**) Scatterplot showing the correlation between validation and testing performance metrics at different sites (MGB-ABC Narrow (“ABC”), Generation Scotland (“GS”), LEOCC, CAMP, Costa Rica (“CRA”)) relative to the MGB-ABC test cohort (“MGB”). Each point represents an individual feature. The diagonal line indicates equal performance between validation and testing. Spearman correlation coefficients between MGB and other cohorts are shown. (**B**) Bar plot showing Spearman correlation coefficients for predicted versus observed values across multiple clinical measures, stratified by cohort. Each point represents a different cohort (ABC, CAMP, CRA, GS, LEOCC, and MGB), with colors and symbols denoting cohort identity. The red dashed line indicates zero correlation. Overall, prediction performance was consistent across most measures and cohorts, with the MGB cohort as a reference.

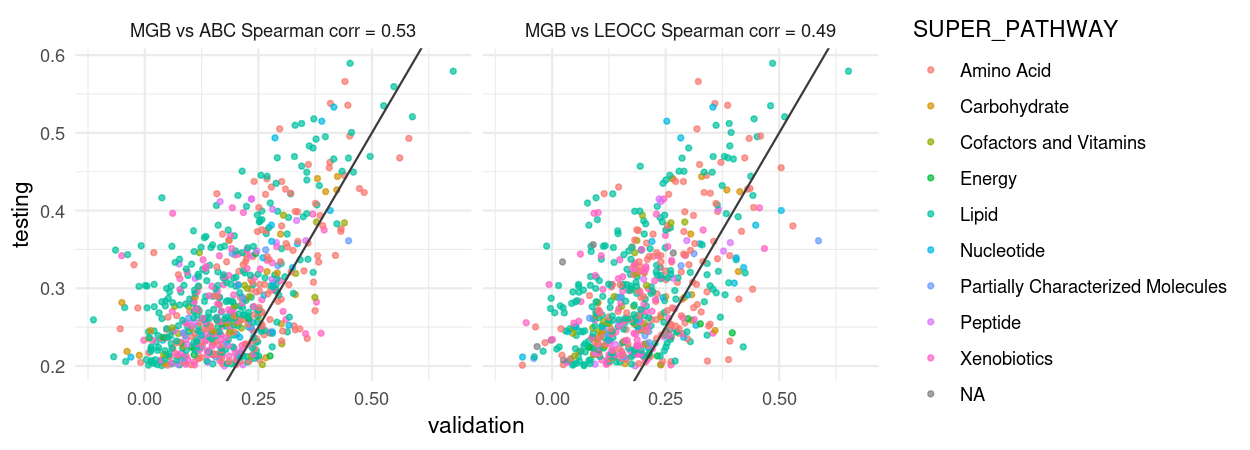

**Supplementary Figure 28. Cross-cohort validation of metabolite model performance by biochemical pathway.** Scatterplots showing correlations between validation and testing performances for metabolite models trained in the MGB cohort and tested in ABC (left) and LEOCC (right) cohorts. Each point represents an individual metabolite, color-coded by its assigned biochemical super pathway. The black diagonal line indicates equal performance between validation and testing. Spearman correlation coefficients are reported in each panel.

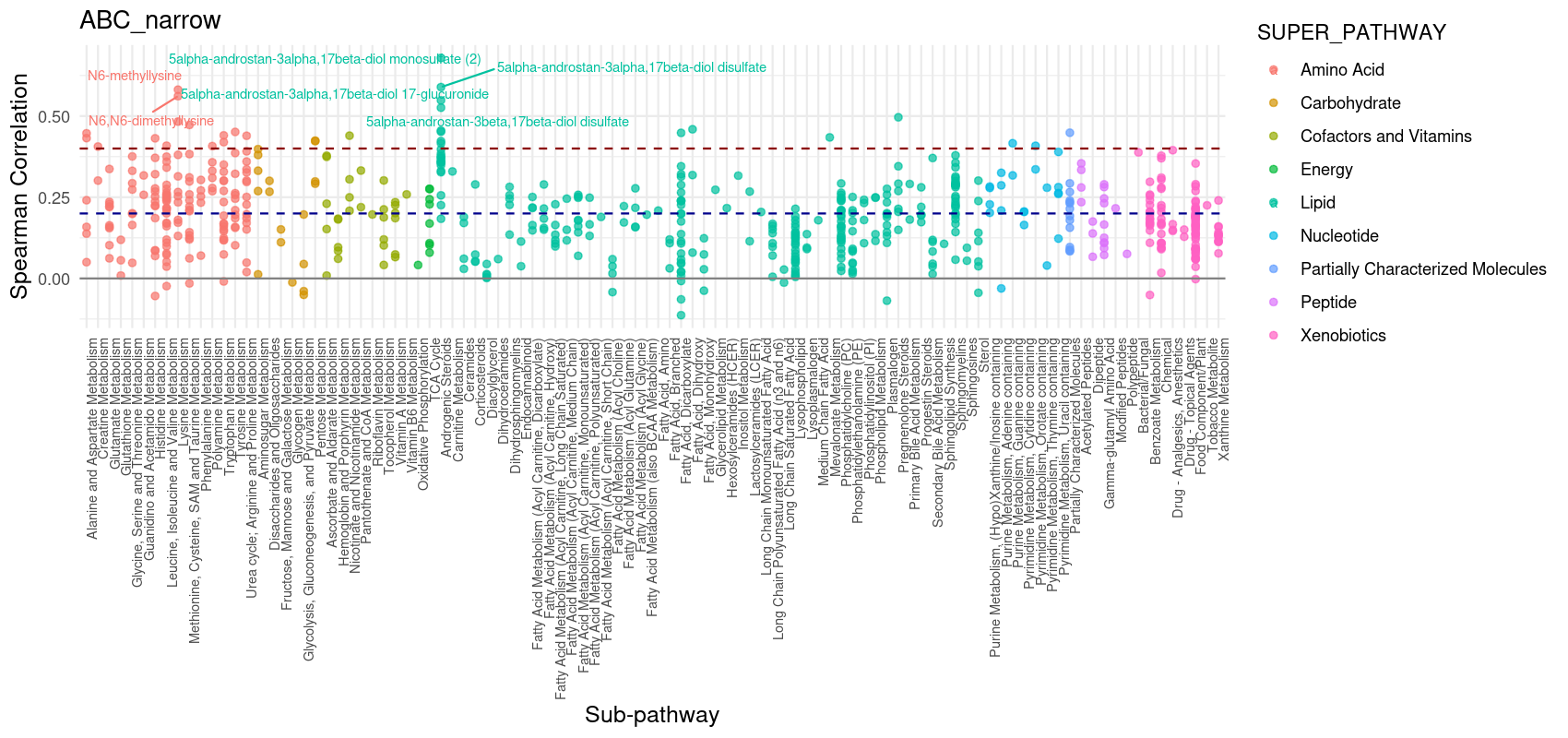

**Supplementary Figure 29. Model performance across metabolite sub-pathways in the ABC cohort.** Spearman correlation coefficients between predicted and observed metabolite levels are shown for the ABC cohort, grouped by biochemical sub-pathway and color-coded by super pathway. Each point represents an individual metabolite. The red and blue dashed lines indicate the mean and median correlation values, respectively. Selected metabolites with the highest correlations are labeled.

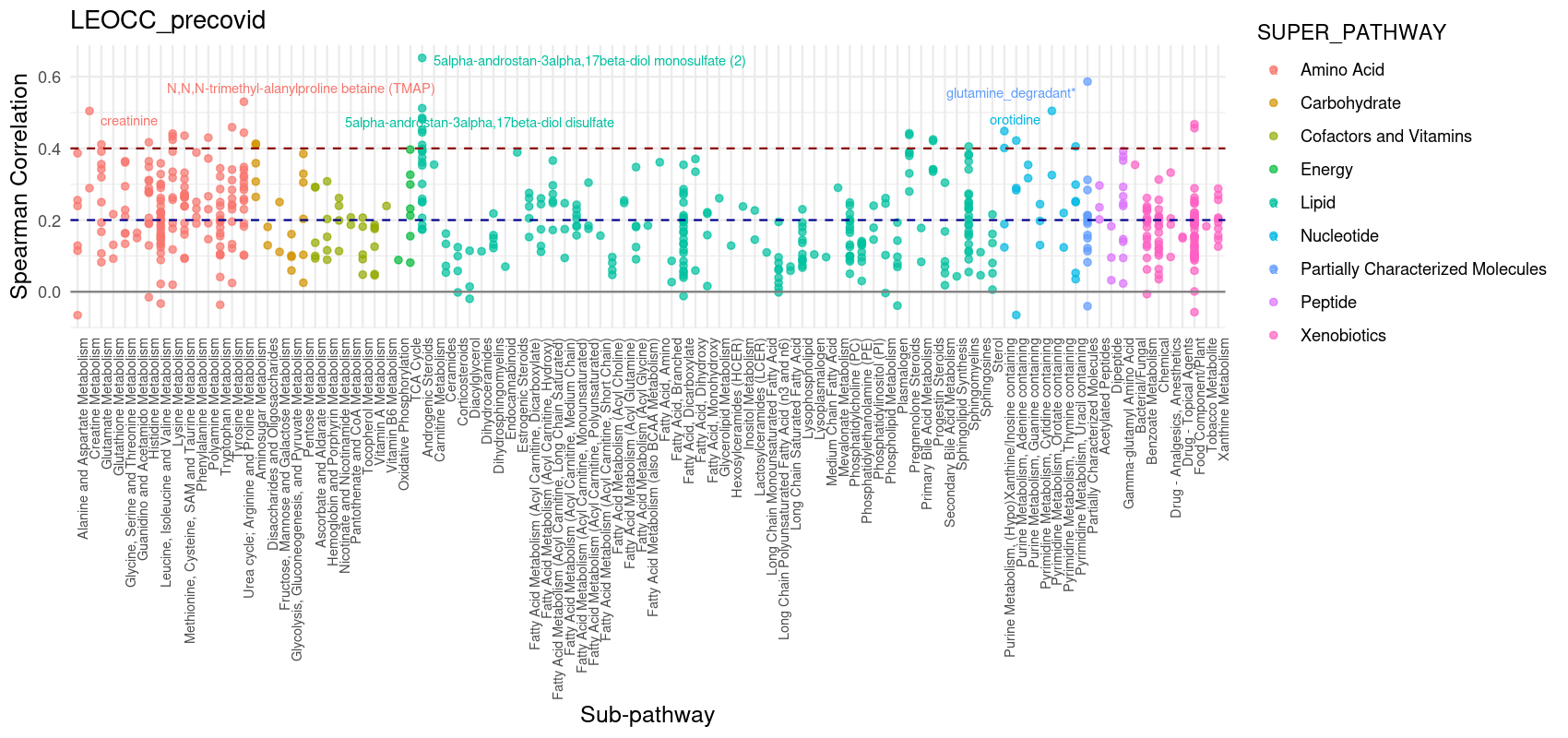

**Supplementary Figure 30. Model performance across metabolite sub-pathways in the LEOCC cohort.** Spearman correlation coefficients between predicted and observed metabolite levels are shown for the LEOCC cohort, grouped by biochemical sub-pathway and color-coded by super pathway. Each point represents an individual metabolite. The red and blue dashed lines indicate the mean and median correlation values, respectively. Labeled metabolites correspond to those with the highest correlations within their respective pathways.

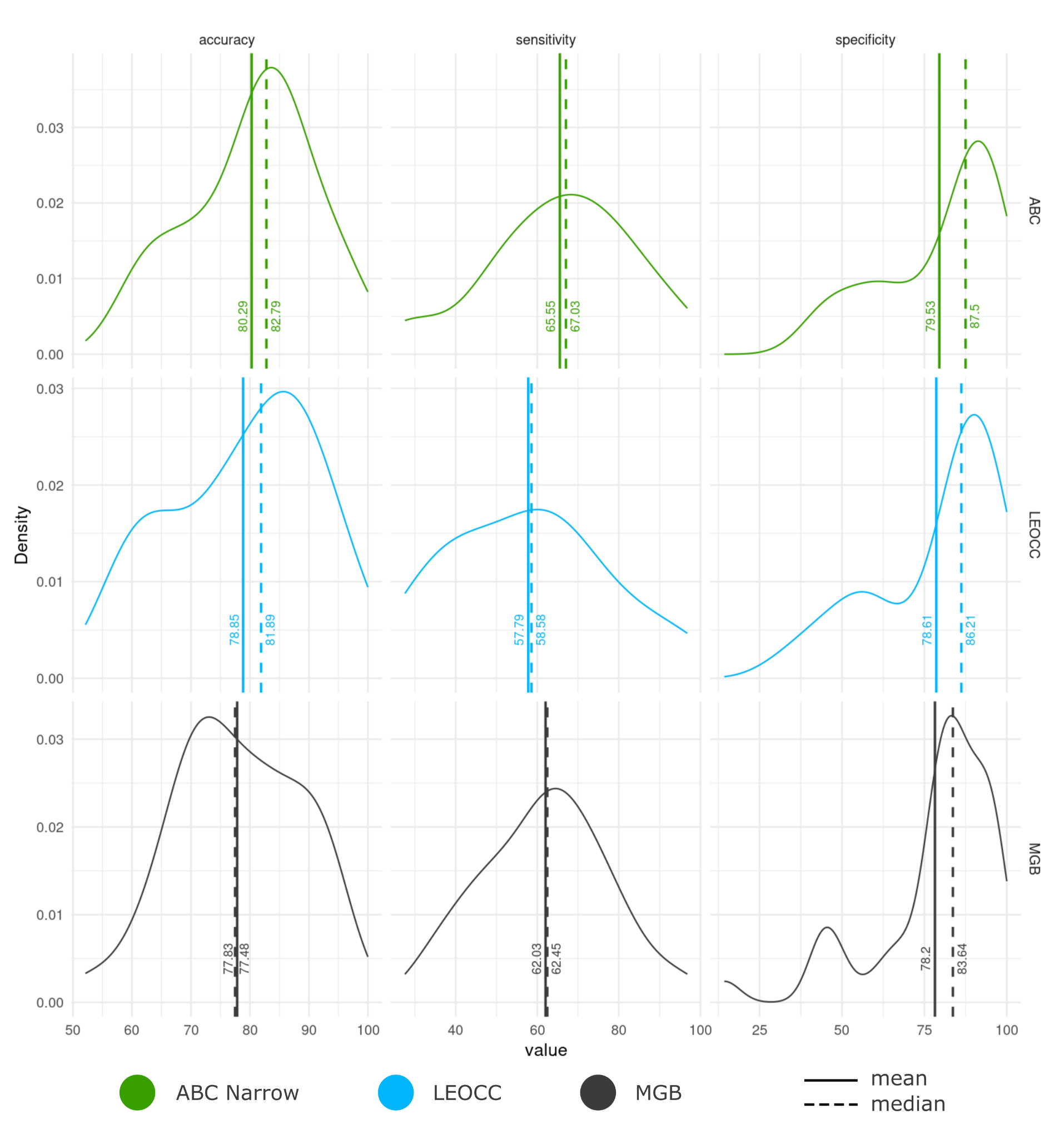

**Supplementary Figure 31. Performance of EBPs in identifying clinically abnormal values across cohorts.** Distribution of accuracy, sensitivity, and specificity across three cohorts (MGB-ABC Narrow, MGB-LEOCC, MGB-ABC). Solid and dashed vertical lines indicate the cohort-specific means and medians, respectively. EBPs generally showed high specificity and moderate-to-high accuracy across cohorts, with greater variability in sensitivity.

**A	B	C**

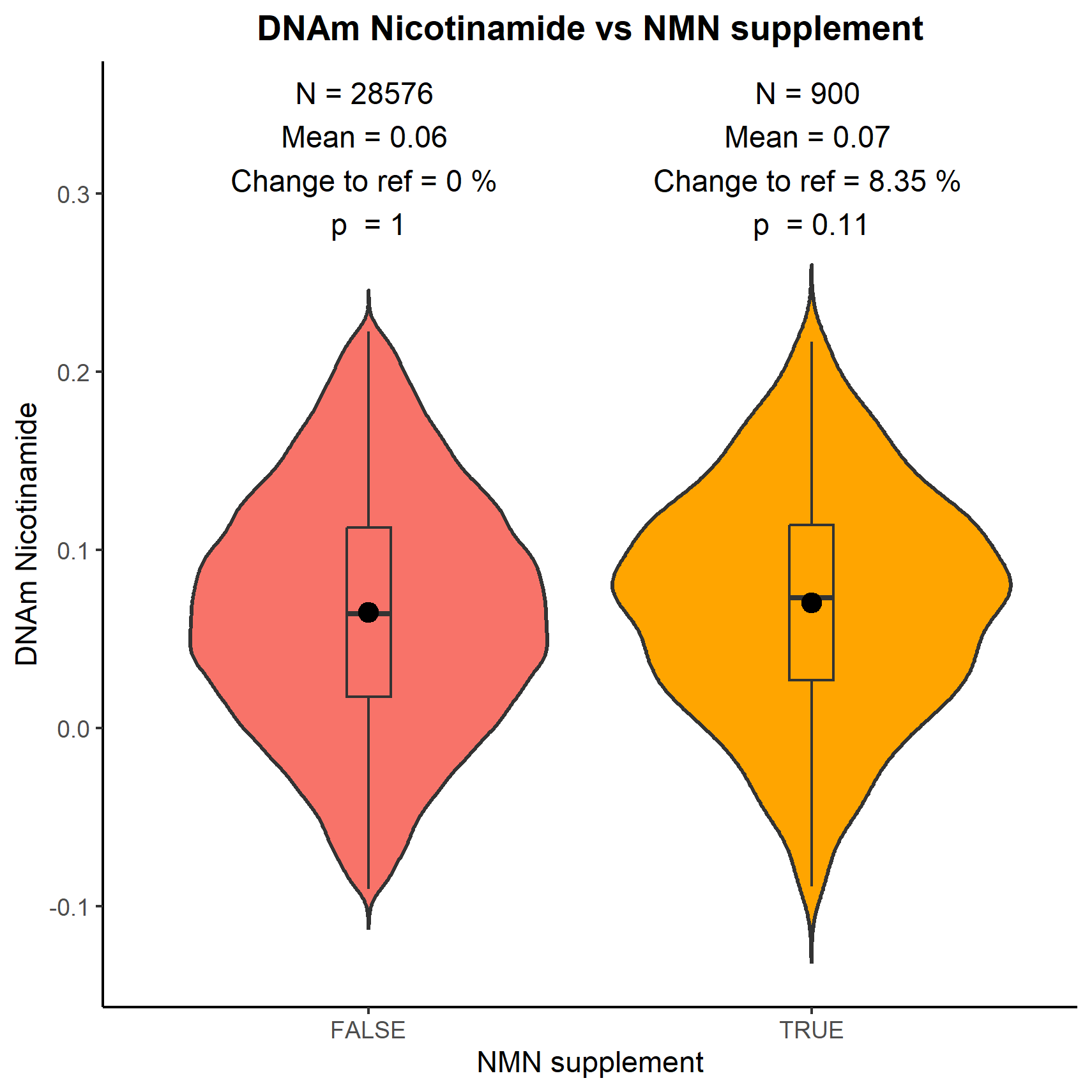

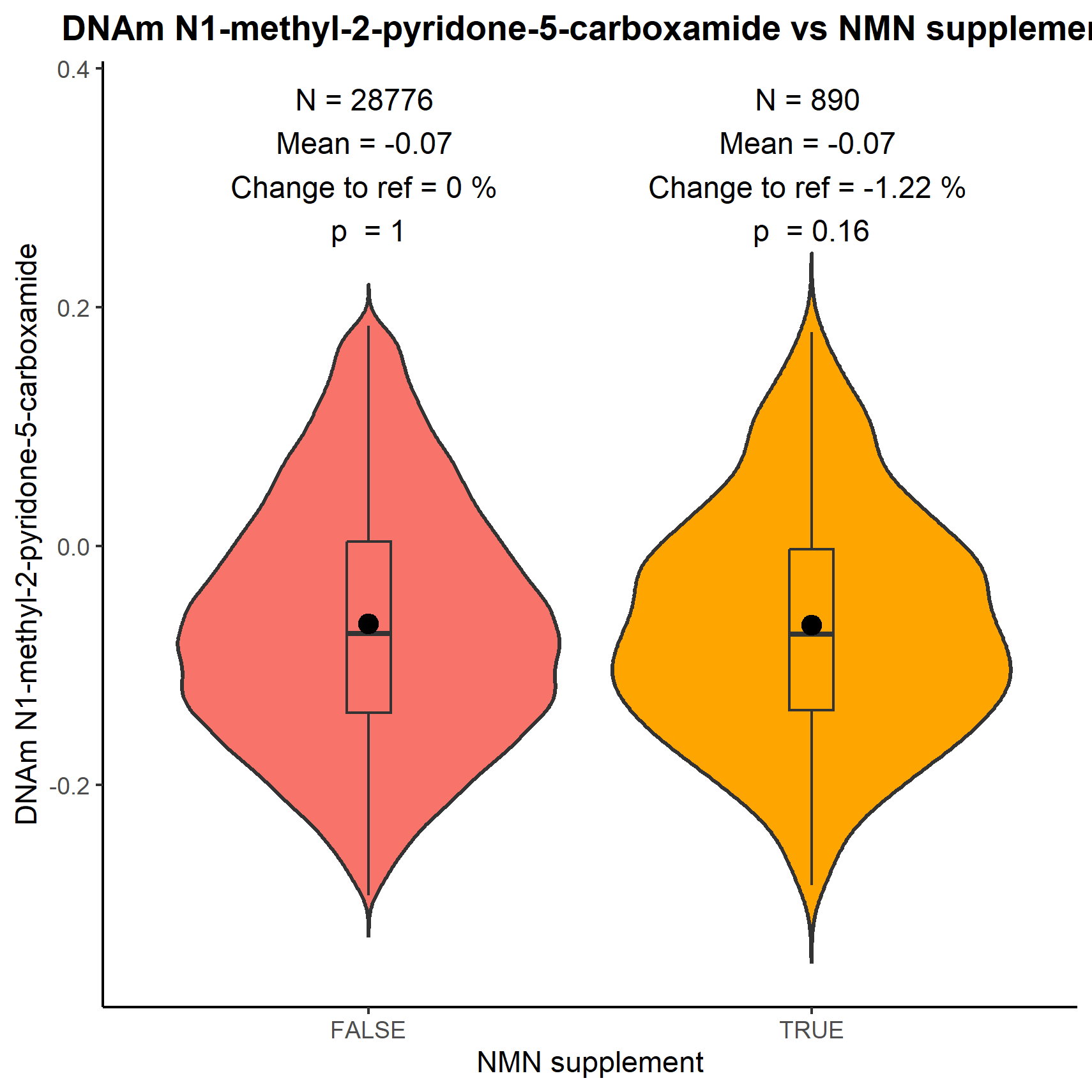

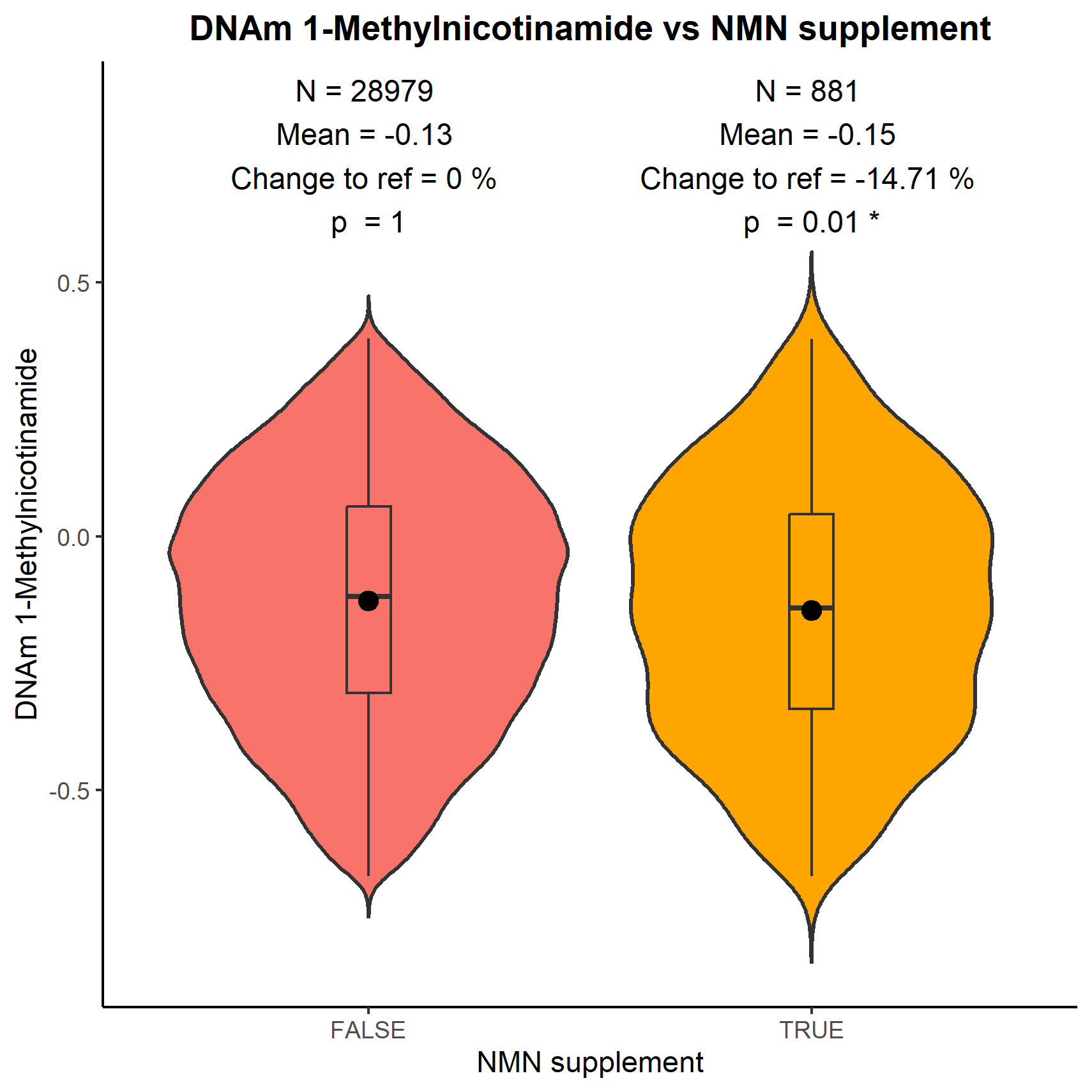

**D	E	F**

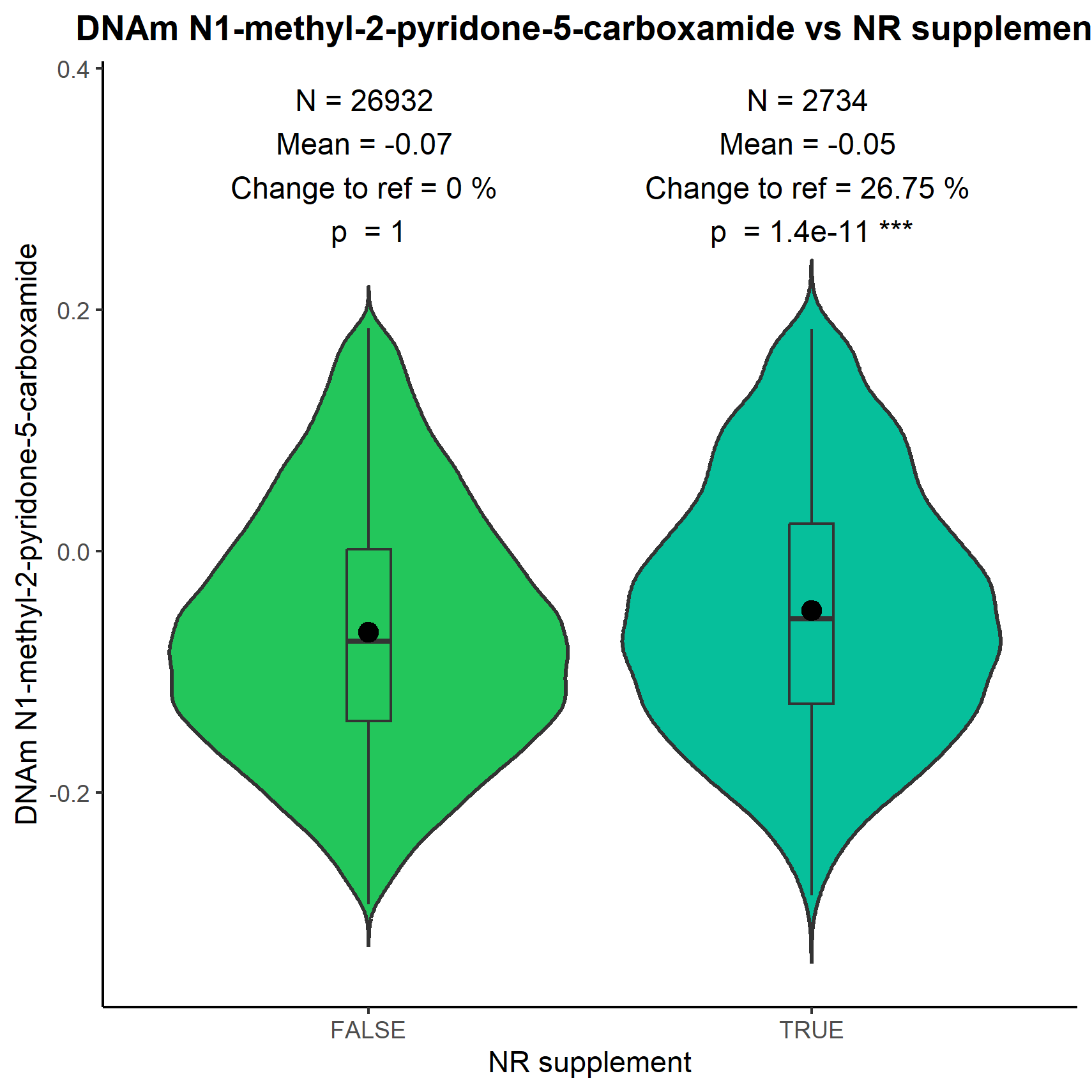

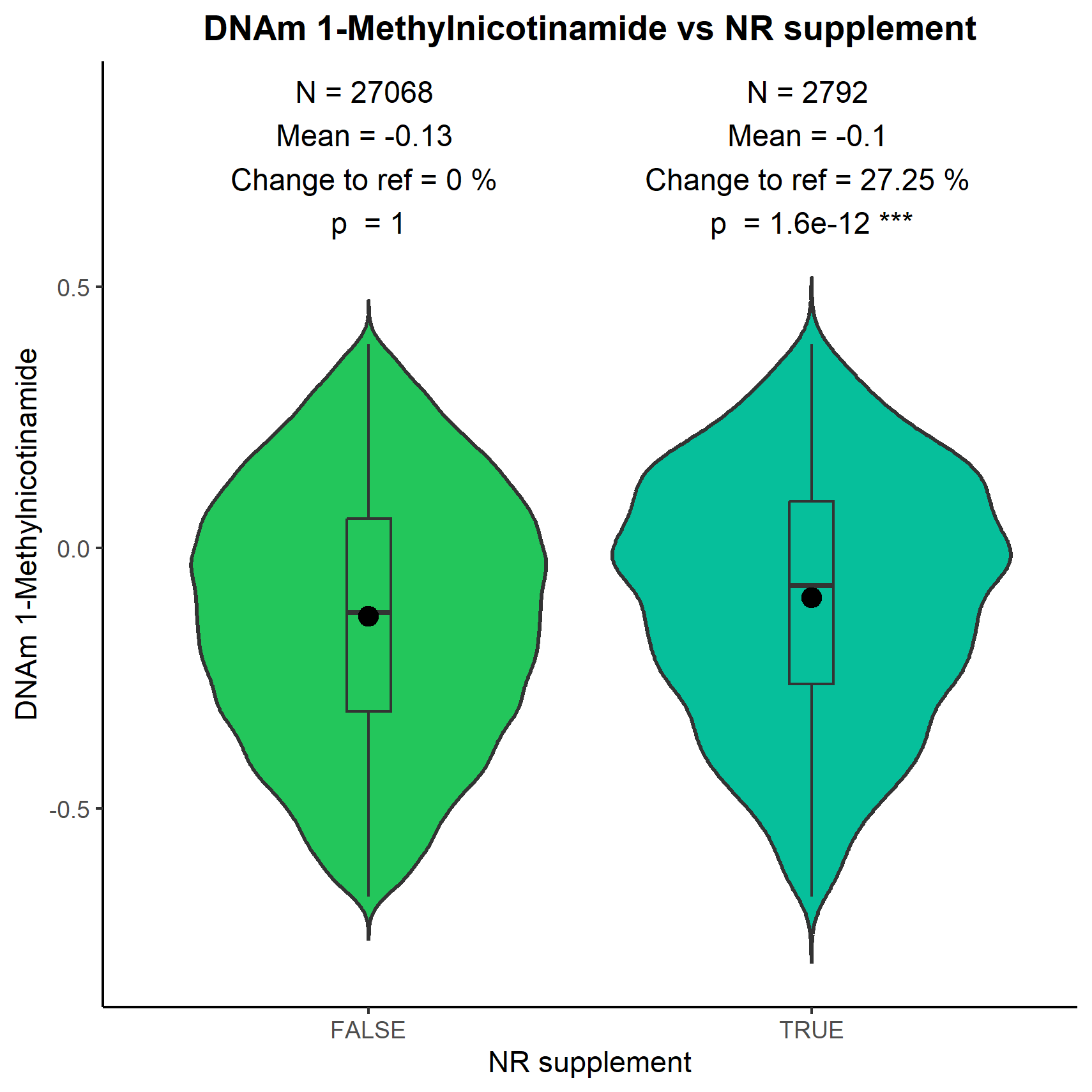

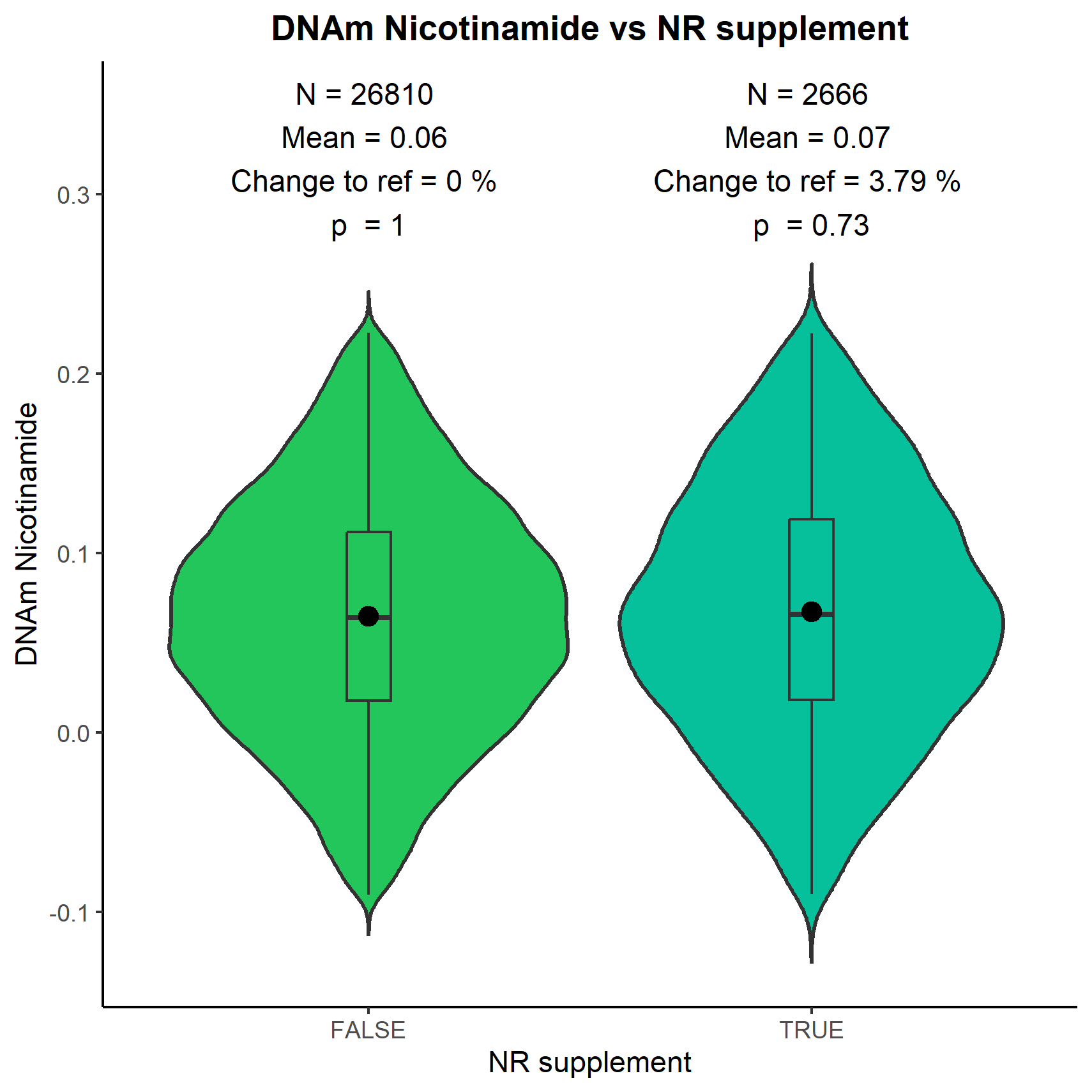

**G	H	I**
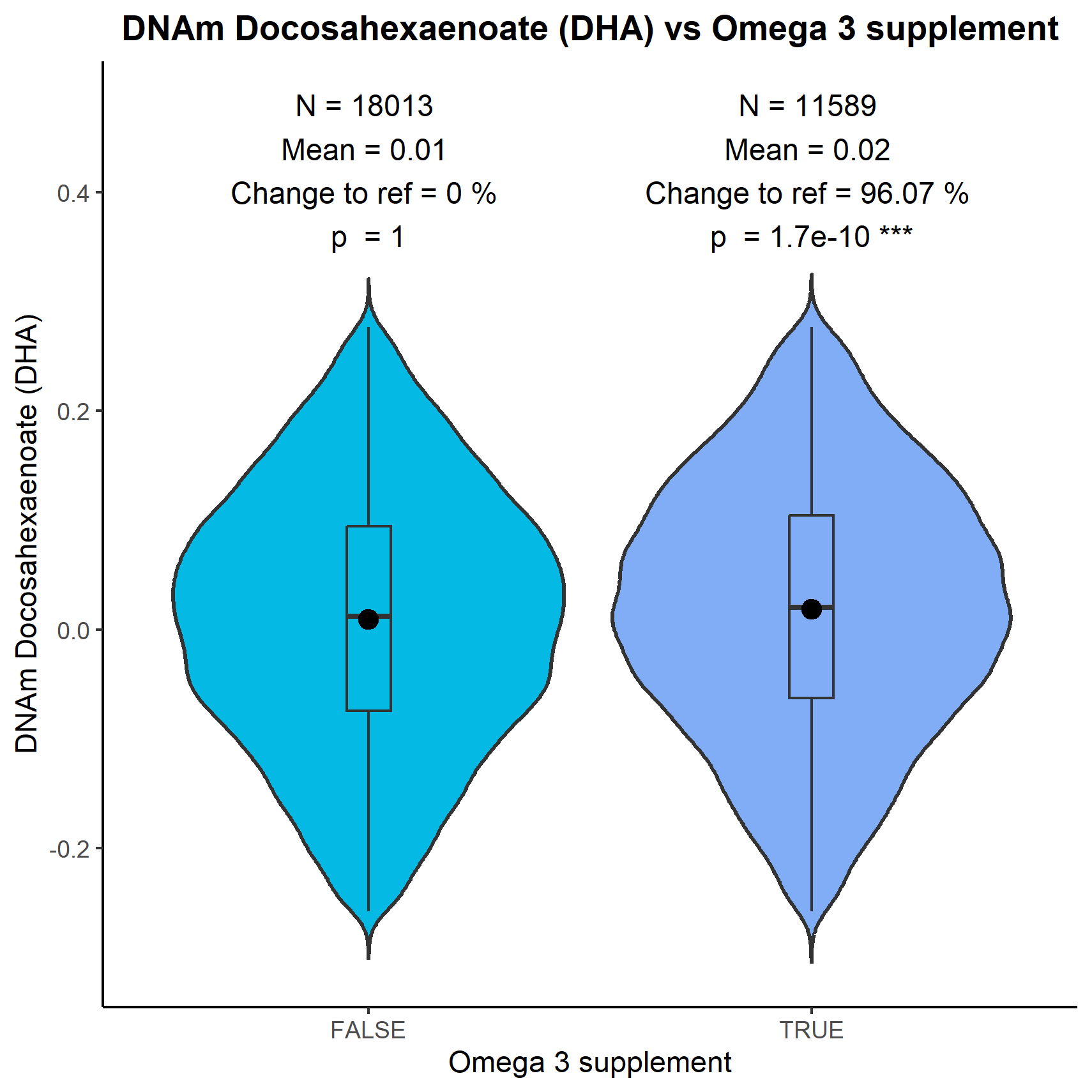

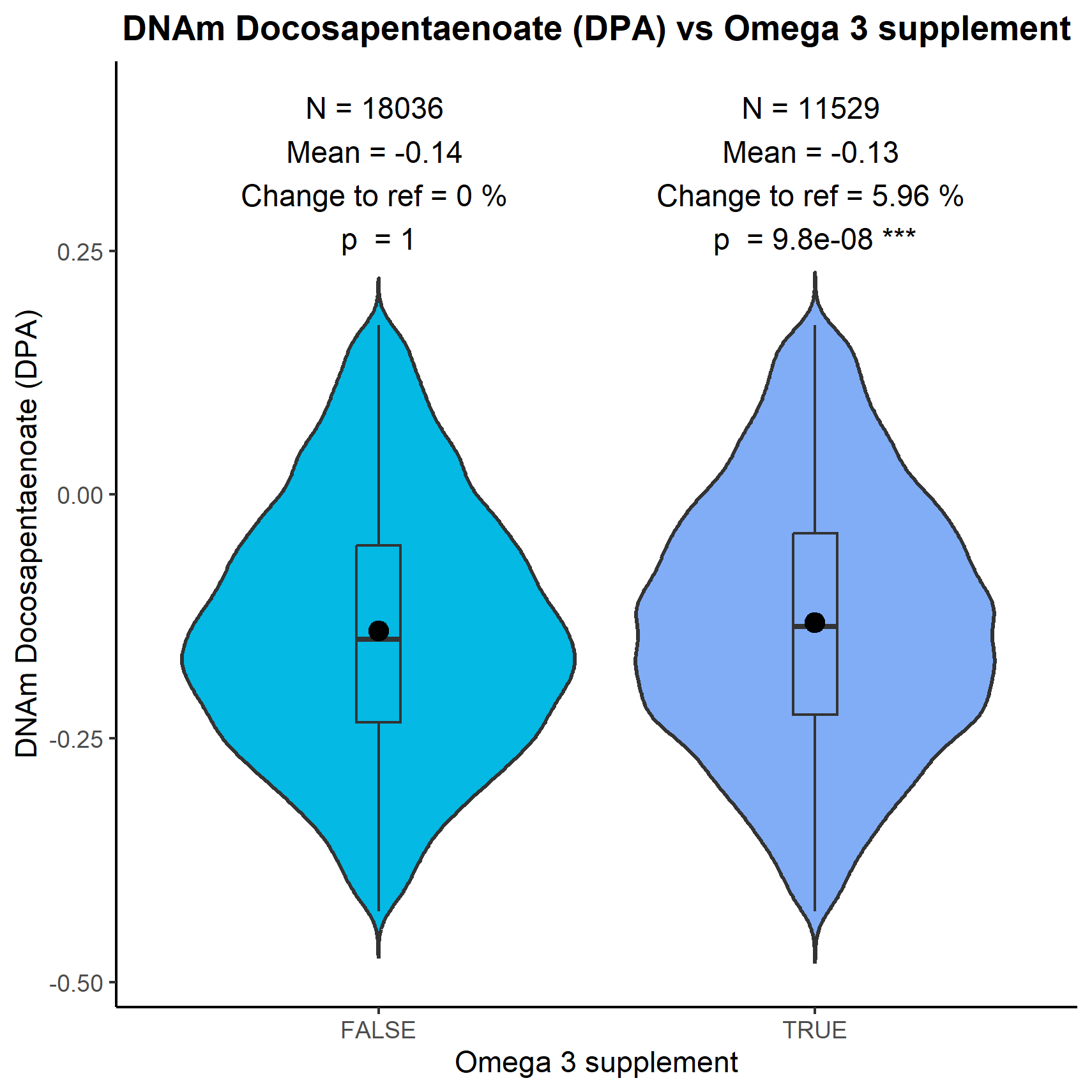

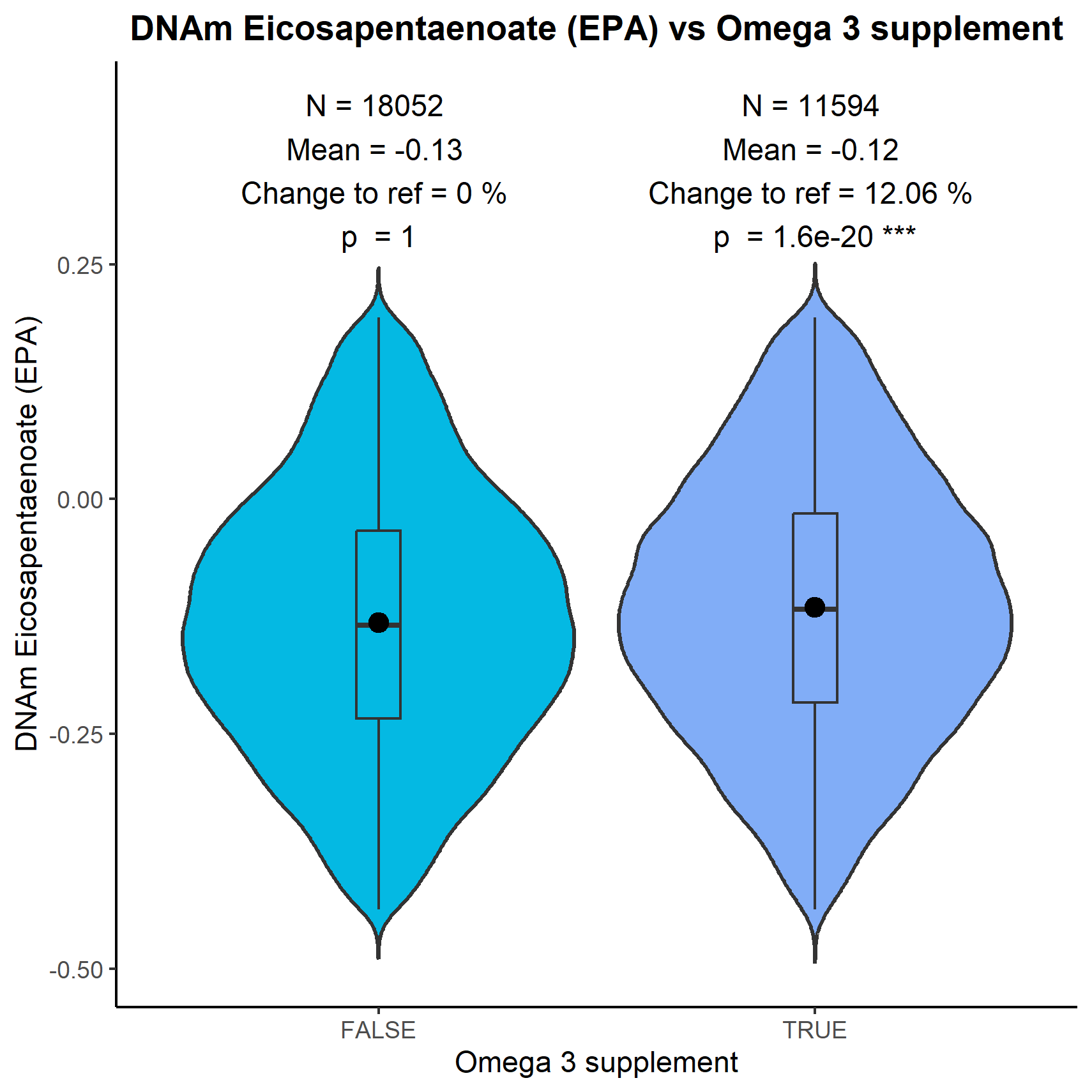

**Supplementary Figure 32.** Validation analyses evaluating the changes of the Epigenetic Biomarker Proxies (EBPs) levels in different scenarios. (**A-C**) Association of Nicotinamide, N1-methyl-2-puridone-5-carboxamide, and 1-Methylnicotinamide EBPs with the supplementation of Nicotinamide Mononucleotide (NMN). (**D-F**) Association of Nicotinamide, N1-methyl-2-puridone-5-carboxamide, and 1-Methylnicotinamide EBPs with the supplementation of Nicotinamide Riboside (NR) (**G-I**) Association of Docosahexaenoic Acid (DHA), Docosapentaenoic Acid (DPA), and Eicosapentaenoic Acid (EPA) with Omega 3 supplementation.

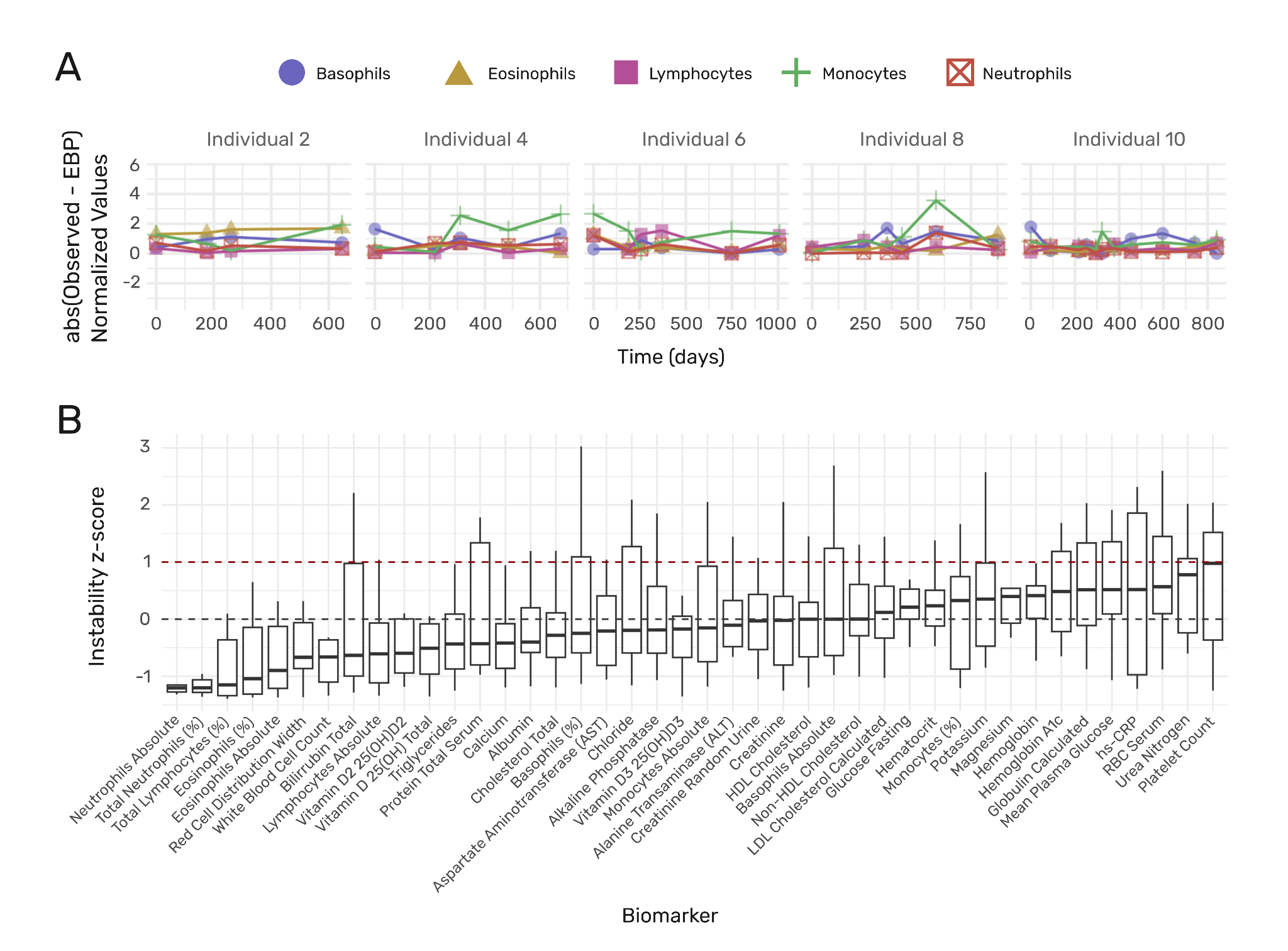

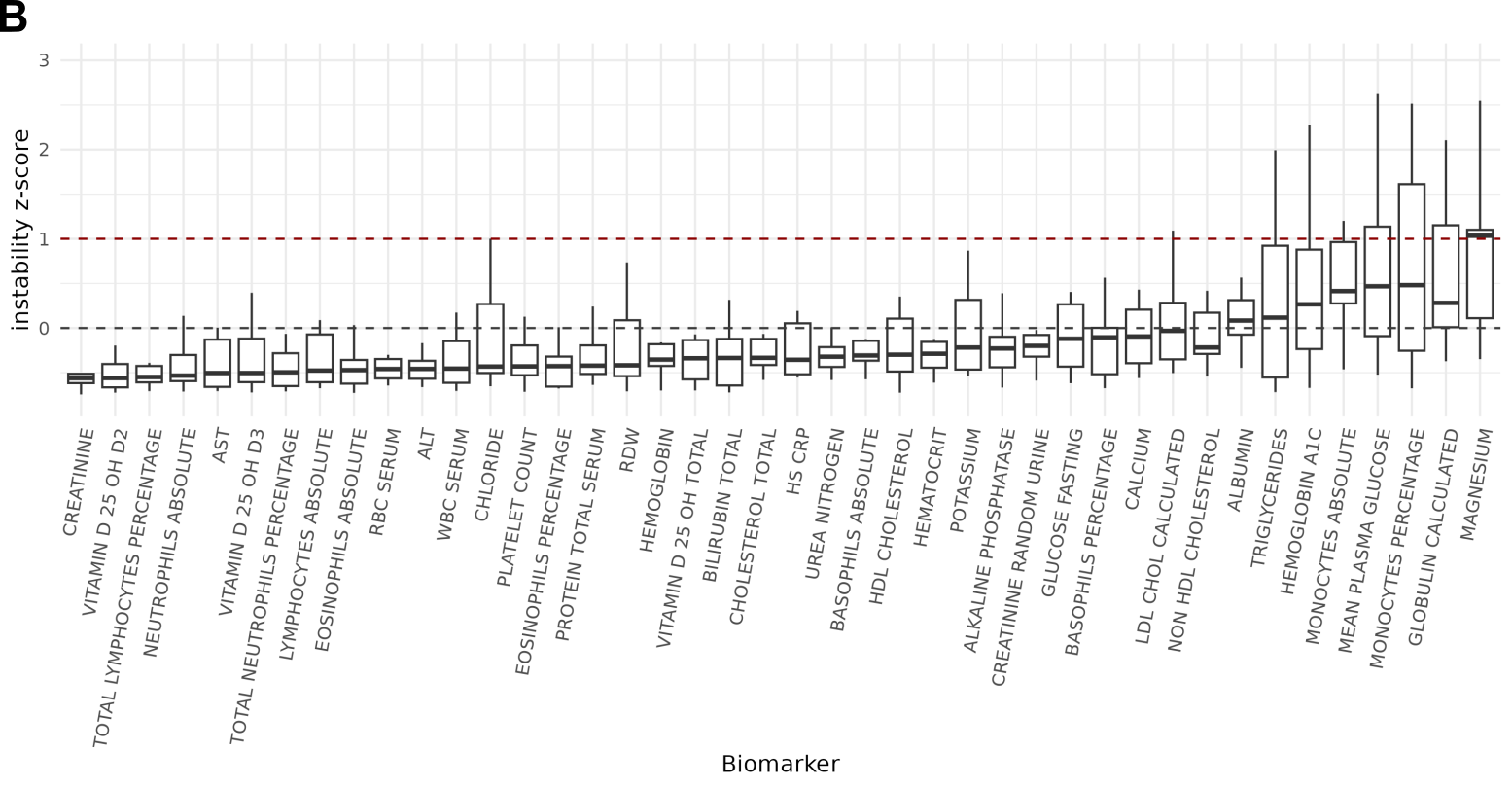

**Supplementary Figure 33. Longitudinal comparison of epigenetic biomarker predictions (EBPs) to observed clinical values. (A)** Absolute differences between observed and EBP-predicted values over time for six white blood cell types in five individuals from the TruDiagnostic cohort with ≥4 paired timepoints. **(B)** Trajectory instability z-scores across all biomarkers. Higher z-scores indicate lower longitudinal stability of the EBP for that marker.
